# Integrating multiple lines of evidence to assess the effects of maternal BMI on pregnancy and perinatal outcomes in up to 497,932 women

**DOI:** 10.1101/2022.07.22.22277930

**Authors:** Maria Carolina Borges, Gemma Clayton, Rachel M Freathy, Janine F Felix, Alba Fernández-Sanlés, Ana Gonçalves Soares, Fanny Kilpi, Qian Yang, Rosemary R C McEachan, Rebecca C Richmond, Xueping Liu, Line Skotte, Amaia Irizar, Andrew T Hattersley, Barbara Bodinier, Denise M Scholtens, Ellen A Nohr, Tom A Bond, M. Geoffrey Hayes, Jane West, Jessica Tyrrell, John Wright, Luigi Bouchard, Mario Murcia, Mariona Bustamante, Marc Chadeau-Hyam, Marjo-Ritta Jarvelin, Martine Vrijheid, Patrice Perron, Per Magnus, Romy Gaillard, Vincent W V Jaddoe, William L Lowe, Bjarke Feenstra, Marie-France Hivert, Thorkild IA Sørensen, Siri E Håberg, Sylvain Serbert, Maria Magnus, Deborah A Lawlor

**Author notes:** Joint first authors (Maria Carolina Borges, Gemma Clayton). Corresponding authors: Dr Maria Carolina Borges and Prof Deborah Lawlor, MRC Integrative Epidemiology Unit, Oakfield House, Oakfield Grove, Bristol, BS8 2BN.

## Abstract

**Importance:** Higher maternal pre-pregnancy body mass index (BMI) is associated with adverse pregnancy and perinatal outcomes. However, which of these associations are causal remains unclear.

**Objective:** To explore the relation of maternal pre-pregnancy BMI with pregnancy and perinatal outcomes by integrating evidence from three different methods (i.e. multivariable regression, Mendelian randomization, and paternal negative control analyses).

**Design:** Triangulation of multivariable regression, Mendelian randomization and paternal negative control results from up to 14 studies in the MR-PREG collaboration.

**Setting:** Europe and North America.

**Participants:** Up to 497,932 women of European ancestry.

**Exposure:** Maternal pre- or early-pregnancy BMI based on self-reported or measured weight and height.

**Main outcomes and Measures:** Miscarriage, stillbirth, hypertensive disorders of pregnancy, gestational hypertension, preeclampsia, gestational diabetes, maternal anaemia, perinatal depression, pre-labour rupture of membranes, induction of labour, caesarean section, preterm birth, small- and large-for-gestational age, low and high birthweight, low Apgar score at 1 and 5 minutes, neonatal intensive care unit admission, and no initiation of breastfeeding.

**Results:** Multivariable regression, Mendelian randomization and paternal negative control analyses supported an association of higher maternal BMI with lower risk of small-for-gestational age and higher risk of hypertensive disorders of pregnancy, gestational hypertension, preeclampsia, gestational diabetes, pre-labour membrane rupture, induction of labour, large-for-gestational age, and high birthweight. As an example, higher maternal BMI was associated with higher risk of gestational hypertension in multivariable regression (OR: 1.67; 95% CI: 1.64, 1.71 per standard unit in BMI) and Mendelian randomization (OR: 1.58; 95% CI: 1.29, 1.93), which was not seen for paternal BMI (OR: 1.02; 95% CI: 0.99, 1.05). Findings did not support a relation between maternal BMI and perinatal depression. For other outcomes, evidence was inconclusive due to inconsistencies across the applied approaches or substantial imprecision in effect estimates from Mendelian randomization.

**Conclusions and Relevance:** Our findings support a causal role for maternal pre-/early-pregnancy BMI on a range of adverse pregnancy and perinatal outcomes. Pre-conception interventions to support women maintaining a healthy BMI may reduce the burden of obstetric and neonatal complications.

**KEY POINTS:** *Question:* What is the effect of higher maternal pre-/early-pregnancy body mass index (BMI) on adverse pregnancy and perinatal outcomes?

*Findings:* We found consistent evidence that higher maternal BMI was related to higher risk of gestational hypertension, preeclampsia, gestational diabetes, pre-labour membrane rupture, induction of labour, and having a large-for-gestational-age baby, lower risk of having a small-for-gestational-age baby, and not related to perinatal depression.

*Meaning:* These findings highlight the importance of supporting women to achieve/maintain a healthy pre-conception BMI to reduce the burden of obstetric and neonatal complications.

## INTRODUCTION

Obesity is a leading preventable cause of ill-health, mortality and morbidity across the world and affects 10% and 25% of adult women in low- and high-income countries, respectively.^1^ Higher maternal pre-pregnancy body mass index (BMI) is associated with higher risk of various adverse pregnancy and perinatal outcomes, including pregnancy loss, gestational hypertension (GH), preeclampsia (PE), gestational diabetes mellitus (GDM), perinatal depression, caesarean deliveries, preterm birth (PTB), large for gestational age (LGA), and no breastfeeding initiation.^2–12^ However, most evidence in the field comes from conventional observational studies, which may be confounded by unmeasured or inaccurately measured maternal characteristics, such as socioeconomic position, age, parity, ethnicity, smoking and alcohol intake.

Understanding the impact of maternal BMI on pregnancy and perinatal health is key to inform appropriate interventions aimed at preventing adverse outcomes and to predict their future burden in different populations. A better understanding of a potential causal role of BMI can be achieved by integrating multiple lines of evidence in a triangulation framework, where more credible inference can be made for findings that agree across different analytical approaches with different strengths and limitations.^13,14^ Whilst findings from conventional observational studies using multivariable regression might be biased by residual confounding, Mendelian randomization studies are less prone to such form of confounding but may be biased by weak instruments or unbalanced horizontal pleiotropy.^15,16^ The use of negative control designs, such as using paternal BMI as a negative control exposure, can reveal bias in associations of maternal BMI with adverse pregnancy and perinatal outcomes since paternal BMI is unlikely to affect these outcomes, but may be associated with unmeasured confounders in a similar fashion to maternal BMI (**Figure 1**). ^17,18^

**Figure 1.**
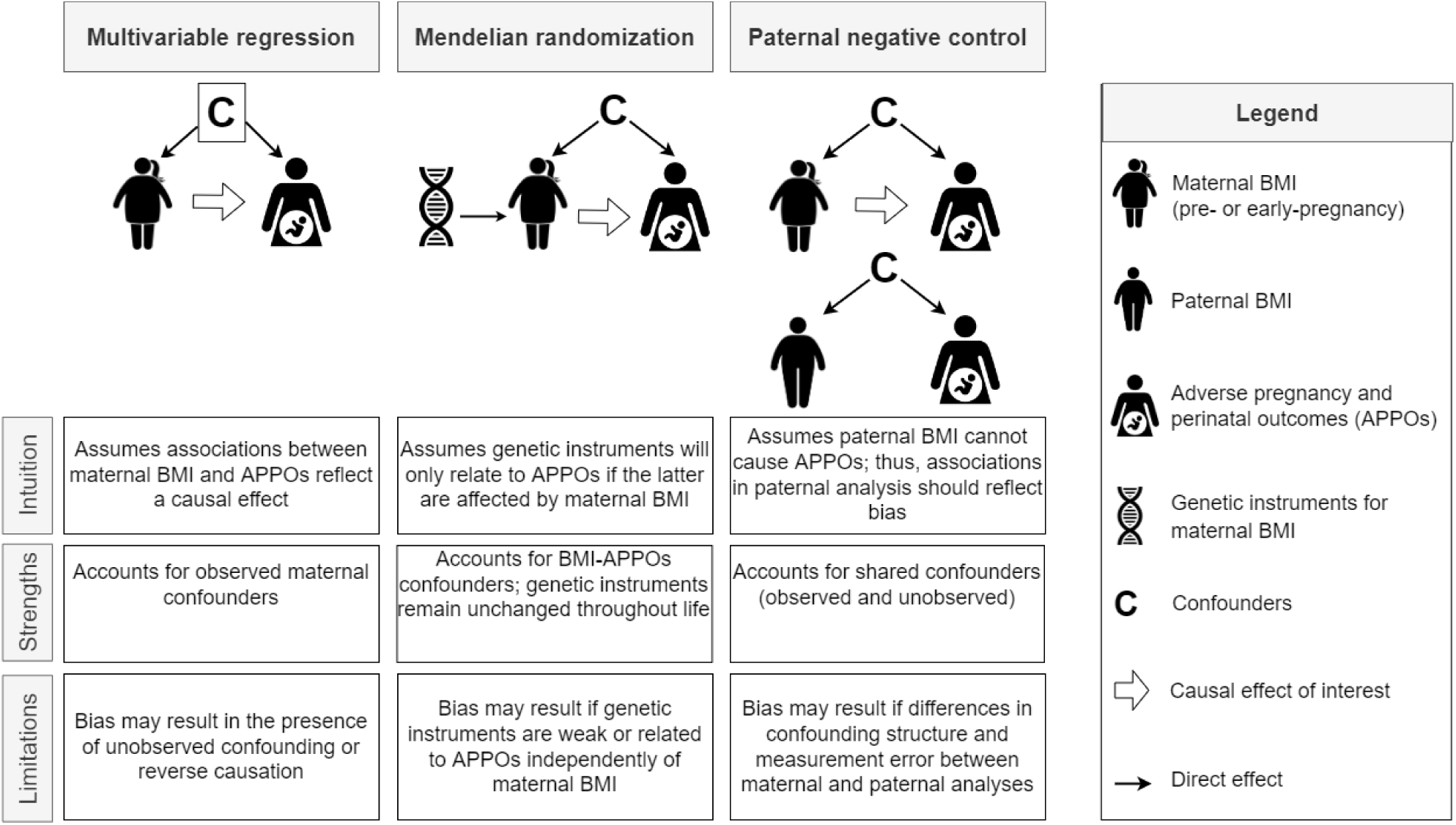
Overview of the three analytical approaches used to investigate the effect of maternal body mass index on adverse pregnancy and perinatal outcomes. A brief description of each approach is presented in the context of exploring the effect of maternal BMI on APPOs risk. Given each approach has different strengths and limitations, findings that agree across approaches are likely to be more credible. The description of each approach is simplified for illustration purposes. An extensive description of assumptions and sources of bias for each approach has been reported previously (e.g. ^17,18,50–52^). The box around the confounders in the multivariable regression reflects that assumption of the method that all confounders were accurately adjusted for in the analyses. BMI: body mass index; APPOs: adverse pregnancy and perinatal outcomes.

The aim of this study was to explore the relation of maternal (pre-/early pregnancy) BMI with a wide range of pregnancy and perinatal outcomes by integrating evidence from multivariable regression, Mendelian randomization and paternal negative control.

## METHODS

### Study Participants

Data were obtained from up to 497,932 women participating in 14 studies in Europe and North America as part of the MR-PREG collaboration (**Table 1**). We included women who had available information on at least one outcome of interest, had a singleton birth, delivered a baby without a severe known congenital anomaly and were of European ancestry since most studies included participants of European descent only or predominantly. Informed consent was obtained from all participants and study protocols were approved by the local, regional or institutional ethics committees. Details of recruitment, data collection and ethical approval of each study can be found in **Supplementary Methods**.

**Table 1.** Characteristics of the included studies.

| Cohort | Source | Country | Maximum N<sup>a</sup> | SNPs set<sup>b</sup> | BMI measurement | Maternal pre-pregnancy BMI [kg/m<sup>2</sup>]<sup>c</sup><br>Mean (SD) | Maternal age at delivery [years]<br>Mean (SD) |
| --- | --- | --- | --- | --- | --- | --- | --- |
| ALSPAC* | 53,54 | United Kingdom | 11,272 | 97 | Self-reported pre-pregnancy | 22.95 (3.82) | 28.46 (4.78) |
| BiB* | 55 | United Kingdom | 3,352 | 97 | Measure around 12 weeks of gestation | 26.63 (5.99) | 26.82 (5.96) |
| DNBC-GOYA* | 56,57 | Denmark | 2,022 | 97 | Self-reported pre-pregnancy | 23.59 (4.30) | 29.67 (4.20) |
| DNBC-PTB (controls)* | 58 | Denmark | 1,676 | 97 | Self-reported pre-pregnancy | 23.44 (3.98) | 29.78 (4.10) |
| EFSOCH* | 59 | United Kingdom | 789 | 97 | Weight self-reported pre-pregnancy, height measured during pregnancy | 24.02 (4.43) | 30.64 (5.03) |
| FinnGen | 60 | Finland | 123,579 | 97 | NA | NA | NA |
| GEN-3G | 61 | Canada | 582 | 97 | Self-reported pre-pregnancy | 25.04 (5.70) | 28.27 (4.34) |
| GenR | 62 | The Netherlands | 3,232 | 32 | Measured before 20 weeks of gestation | 25.31 (4.89) | 28.50 (5.66) |
| HAPO* | 63 | USA | 1,309 | 97 | Measured between 24 and 32 weeks of gestation | 28.46 (4.82) | 31.31 (5.27) |
| INMA | 64 | Spain | 1,039 | 32 | Self-reported pre-pregnancy weight, height measured in the first trimester | 23.37 (4.25) | 30.72 (4.02) |
| MoBa* | 65,66 | Norway | 81026 | 97 | Self-reported at 15 weeks gestation | 24.05 (4.32) | 30.13 (4.72) |
| NFBC1966 | 67 | Finland | 2,304 | 97 | Self-reported pre-pregnancy | 25.14 (4.78) | 30.22 (5.9) |
| NFBC1986 | 68 | Finland | 888 | 97 | Self-reported pre-pregnancy | 24.17 (4.61) | 25.43 (2.64)<sup>d</sup> |
| UK Biobank | 69 | United Kingdom | 211,553 | 97 | NA | NA | 29.03 (6.34)<sup>e</sup> |

<sup>a</sup> Maximum number of mothers with at least one outcome (N shown for outcome with highest sample size when more than outcome was available).
<sup>b</sup> SNPs were selected from the main GWAS meta-analyses of Locke et al. (2015)<sup>70</sup> (97 SNPs; 339,226 individuals; 95% European ancestry) and Speliotes et al. (2010)<sup>71</sup> (32 SNPs; 249,796 individuals; 100% European ancestry). To maximise the number of studies included we prioritised the 97 BMI SNPs<sup>70</sup>, however studies that had directly genotyped the 32 BMI SNPs<sup>23</sup> were also included.
<sup>c</sup> Maternal BMI is only reported where collected pre- or early in pregnancy.
<sup>d</sup> The relatively young age at delivery in the NFBC1986 cohort may be explained by the young age of the cohort at the time of the study.
<sup>e</sup> Maternal age in UK Biobank was taken from maternity record linkage on a subsample of participants and may therefore not be representative of the full sample included.
\* Studies contributing to Mendelian randomization analyses adjusted by offspring genotype.
Abbreviations: ALSPAC, Avon Longitudinal Study of Parents and Children; BiB, Born in Bradford; DNBC-GOYA, Danish National Birth Cohort-Genetics of Obesity in Young Adults Study; DNBC-PTB, Danish National Birth Cohort-Preterm Birth Study; EFSOCH, Exeter Family Study of Childhood Health; FinnGen: FinnGen (release 5); GEN-3G, Genetics of Glycaemic Regulation in Gestation and Growth; GenR: Generation R; HAPO, Hyperglycaemia and Adverse Pregnancy Outcome; INMA, Infancia y Medio Ambiente (English translation = Childhood and the Environment); MoBa: Norwegian Mother, Father and Child Cohort Study; NA: not available; NFBC1966, Northern Finland 1966 Birth Cohort; NFBC1986, Northern Finland 1986 Birth Cohort.

### Exposure measures

Maternal BMI in kg/m^2^ was calculated from measured or self-reported weight and height data (**Table 1**). Weight data was collected before pregnancy in nine studies, before 20 weeks of gestation in three studies, and between 24 and 32 weeks of gestation in one study. Two studies did not have a measure of pre- or early-pregnancy BMI and could only contribute to the Mendelian randomization analyses.

### Outcomes measures

We focused on 20 *a priori* selected binary outcomes: miscarriage, stillbirth, hypertensive disorders of pregnancies (HDP), GH, PE, GDM, maternal anaemia, perinatal depression, pre-labour membrane rupture, induction of labour, caesarean section, PTB, LGA, small-for-gestational age (SGA), low birthweight, high birthweight, low Apgar score after 1 minute, low Apgar score after 5 minutes, neonatal intensive care unit (NICU) admission, and breastfeeding initiation (see **Table 2** for definitions and total sample sizes). We included related traits among the selected outcomes to maximise the number of cohorts contributing to the analyses (e.g. studies that did not have data on gestational age could contribute with information on low birthweight but not SGA). In additional analyses, we examined four continuous traits that underlie some of these outcomes (i.e. birthweight, birth length, ponderal index at birth, and gestational age at birth). Details on outcomes definitions, distributions and sample sizes for each contributing study are available in **Supplementary Methods** and **Supplementary Tables 1A** and **1B**.

**Table 2.** **Case definition and sample size for pregnancy and perinatal outcomes across participating studies**

| Outcomes | Case definition | N | N cases | % cases |
| --- | --- | --- | --- | --- |
| *Binary outcomes* |  |  |  |  |
| Miscarriage<sup>a</sup> | Self-reported in <u>index pregnancy</u> | 91,757 | 107 | 0.12% |
|  | Self-reported in <u>previous pregnancies</u> | 459,012 | 83,237 | 18.13% |
| Stillbirth<sup>a</sup> | Self-reported in <u>index pregnancy</u> | 98,199 | 315 | 0.32% |
|  | Self-reported in <u>previous pregnancies</u> | 342,518 | 7,441 | 2.17% |
| Hypertensive disorders of pregnancy | Gestational hypertension or preeclampsia<sup>b</sup> | 225,554 | 20,920 | 9.27% |
| Gestational hypertension | Elevated blood pressure without proteinuria<sup>b</sup> | 217,439 | 13,998 | 6.44% |
| Preeclampsia | Elevated blood pressure with proteinuria<sup>b</sup> | 213,404 | 7,206 | 3.38% |
| Gestational diabetes | Hyperglycaemia first diagnosed in pregnancy<sup>c</sup> | 497,932 | 9,163 | 1.84% |
| Maternal anaemia | Hb <105 g/L in the 2<sup>nd</sup> or 3<sup>rd</sup> trimesters<sup>d</sup> | 92,002 | 2,425 | 2.64% |
| Perinatal depression | Self-reported diagnosis or assessed depression symptom scales | 90,204 | 4,780 | 5.30% |
| Pre-labour rupture of membranes* | Membrane rupture before the onset of contractions | 196,292 | 16,221 | 8.26% |
| Induction of labour* | Labour needed induction | 114,075 | 17,351 | 15.21% |
| Caesarean section* | Delivery by caesarean section | 174,486 | 22,785 | 13.06% |
| Preterm birth* | Gestational age at birth < 37weeks | 214,828 | 11,242 | 5.23% |
| Large-for-gestational age* | > 90<sup>th</sup> percentile for z-score of birthweight accounting for sex and gestational age | 118,667 | 12,386 | 10.44% |
| Small-for-gestational age* | < 10<sup>th</sup> percentile for z-score of birthweight accounting for sex and gestational age<sup>e</sup> | 118,667 | 8,958 | 7.55% |
| Low birthweight* | Birthweight < 2500g | 322,370 | 21,864 | 6.78% |
| High birthweight* | Birthweight ≥ 4000g | 322,168 | 24,936 | 7.74% |
| Low Apgar score at 1 minute* | Apgar score at 1 minute < 7 | 98,868 | 5,760 | 5.83% |
| Low Apgar score at 5 minutes* | Apgar score at 5 minutes < 7 | 101,949 | 1,187 | 1.16% |
| Neonatal intensive care unit (NICU) admission* | Admitted to NICU | 93,522 | 8,262 | 8.83% |
| Breastfeeding initiation* | Ever breastfed | 94,116 | 78,472 | 83.38% |
| *Continuous outcomes* |  |  |  |  |
| Birthweight* | NA | 326,537 | NA | NA |
| Birth length* | NA | 95,649 | NA | NA |
| Ponderal index at birth* | NA | 95,562 | NA | NA |
| Gestational age at birth* | NA | 118,723 | NA | NA |

Detailed information on each of the outcomes in each cohort is provided in **Supplementary material** and **supplementary table 1**.
<sup>a</sup> Miscarriage and stillbirths in the index pregnancy were used in multivariable regression and paternal negative control analyses, while miscarriage and stillbirth reported in previous pregnancies were used in Mendelian randomization analysis.
<sup>b</sup> Where possible, we applied the International Society for the Study of Hypertension in Pregnancy criteria (ISSHP), which defines any HDP as SBP ≥140mmHg or DBP ≥90mmHg, measured on two occasions after 20 weeks gestation, with those who are then defined as having pre-eclampsia also having proteinuria (with the raised blood pressure) of at least 30g/Dl and those defined as having gestational hypertension being those who do not meet criteria for pre-eclampsia. By contrast, in some studies (e.g. UK Biobank), information on diagnosis was extracted directly from medical records.
<sup>c</sup> Criteria used to define hyperglycaemia first diagnosed in pregnancy varies across studies. Most studies obtained information from questionnaires (i.e. self-reported diagnosis) or from medical records [ICD-10 code O24].
<sup>d</sup> Pavord et al (2019) UK guidelines on the management of iron deficiency in pregnancy. British Journal of Haematology. https://doi.org/10.1111/bjh.16221
<sup>e</sup> Different reference populations were used to calculate percentiles as detailed in Supplementary methods.
\*For these *a priori* selected outcomes, additional Mendelian randomization analyses were conducted accounting for offspring genotype.

### Covariables

The following were *a priori* considered potential confounders of the association between maternal BMI and the pregnancy and perinatal outcomes: maternal age, parity, education, smoking during pregnancy, and alcohol use during pregnancy. We also adjusted for offspring sex to improve statistical efficiency given its strong association with some outcomes (e.g. birthweight-related outcomes). Details of the distribution of these covariables in each study are provided in **Supplementary Table 2**.

### Statistical analyses

All analyses were conducted using Stata version 17 (StataCorp, College Station, TX)^19^ or R version 4.1.1 (R Foundation for Statistical Computing, Vienna, Austria).^20^ Results are presented as odds ratio (OR) for each binary outcome per standard deviation (SD) increase in maternal BMI to facilitate the comparison of results.

#### Multivariable regression analyses

In the main analyses, we used logistic regression with two sets of adjustments: (1) maternal age and offspring sex, and (2) additionally maternal education, parity, smoking during pregnancy and alcohol use during pregnancy where available. We present the fully adjusted model as the main analyses and include the minimally adjusted model in the supplementary material. Similar multivariable linear regression models were used for the additional analyses with continuously measured outcomes. Study-specific results were combined using fixed-effects metanalyses (inverse-variance weighted) for the main analyses assuming that there is one true effect size underlying all included studies, and random-effects metanalyses (DerSimonian and Laird method) for sensitivity analyses.

#### Mendelian randomization analysis

We used two-sample Mendelian randomization, in which the effect of interest is estimated by combining summary data for the association of single nucleotide polymorphisms (SNPs) with BMI and with each outcome.^21^ This approach allowed us to maximise statistical power by including all 14 studies in the analyses even when data on pre- or early-pregnancy BMI was not available (i.e. FinnGen and UK Biobank).

We selected SNPs previously reported to be strongly associated with BMI (P < 5 × 10^−8^) from two genome-wide association studies (GWAS) conducted by the Genetic Investigation of ANthropometric Traits (GIANT) consortium.^22,23^ In 12 studies, data were available for the set of 97 SNPs reported in the larger GWAS^22^, and in two (Generation R and INMA) for the set of 32 SNPs reported in the earlier GWAS^23^ (**Table 1**). Unlike the most recent BMI GWAS^24^, the cohorts included in these two GWAS^22,23^ were largely independent from the included studies for our current pregnancy/delivery outcomes meta-analyses avoiding potential biases due to sample overlap. Details of the SNPs selected and genetic association with BMI (from GIANT and from our current study) are provided in **Supplementary Table 3**.

Summary data for the SNP-BMI associations were obtained from the GIANT GWAS metanalyses of European females (**Supplementary table 3**)^22^, which included up to 171,977 women (∼0.5% of participants were also included in our study). We estimated the strength of the genetic instruments using the mean F-statistic and total R^2^ for the SNP-BMI association in the GIANT GWAS results as previously described.^25,26^ We also examined the correlation between SNP-BMI estimates in non-pregnant (data from the GIANT consortium) and pregnant women (data from participating cohorts where information on pre- or early-pregnancy BMI was available).

Summary data for the SNP-outcomes associations were obtained from each contributing study using logistic (or linear) regression assuming an additive model. For the 97 SNPs subset, we meta-analysed cohort-specific SNP-outcome associations using inverse-variance weighted fixed-effects for the main analyses and random-effects (DerSimonian and Laird method) for sensitivity analyses. The two studies for which only data on the set of 32 SNPs was available could contribute with 12 overlapping SNPs in the metanalyses.

The main two-sample MR analyses were carried out using the inverse variance weighted (IVW) method^27^. In addition, we also conducted a leave-one-out analysis at the study-level where the pooled IVW estimates were re-computed removing one study at a time to check whether pooled results were driven by a single study.

We conducted a series of sensitivity analyses to explore the plausibility of the core Mendelian randomization assumption that any effect of SNPs on the outcomes is fully mediated by maternal BMI. We explored the potential presence of invalid instruments (e.g. due to SNPs affecting the outcomes through pathways not mediated by BMI) by: (i) assessing between-SNP heterogeneity and directional pleiotropy in effect estimates using Cochran’s Q-statistic and the MR-Egger intercept test^28^, respectively; (ii) checking for the presence of outlying SNPs using leave-one-out analysis at the SNP level; (iii) using other Mendelian randomization methods that are more robust to invalid instruments than IVW (MR-Egger^28^, weighted median^29^ and weighted mode^30^). For offspring outcomes (**Table 2**), we repeated the IVW analyses using summary data for the SNP-outcomes associations adjusted for offspring genotype since maternal BMI genetic variants might influence offspring outcomes (e.g. birthweight) due the fetus inheriting these variants from the mother rather than due to a causal effect of maternal BMI influencing the intra-uterine environment.^31–33^

#### Paternal negative control analyses

We used paternal BMI as a negative control exposure to explore whether the associations of maternal BMI with pregnancy and perinatal outcomes could be explained by residual confounding due to shared familial environment influencing BMI in both partners.^18,34^ These analyses included paternal BMI data from ALSPAC (N=2,821-6,952), calculated from weight and height self-reported by the father during the first trimester, GenR (N =596-911), measured during the first trimester, and MoBa (N = 39,243-57,170), reported by the mother at 15 weeks of gestation. We used multivariable regression to estimate the association of paternal BMI with the outcomes of interest adjusting (where available) for paternal age, number of children, education, smoking and alcohol intake around the time of their partners pregnancy, as well as their partners’ BMI to account for the correlation between maternal and paternal BMI due to assortative mating or shared lifestyle^34,35^ (correlation coefficients ranging from 0.17 in ALSPAC to 0.24 in MoBa). Results were then contrasted between the adjusted maternal and paternal BMI (negative control) analyses. Similar estimates between maternal and paternal BMI analyses indicate maternal BMI is unlikely to be a cause of pregnancy and perinatal outcomes via intrauterine mechanisms assuming comparable sources of biases. Conversely, associations that are specific or stronger in the maternal compared to the paternal BMI analyses would support a causal effect of maternal BMI.

## RESULTS

### Study and participants characteristics

The characteristics of the 14 included studies are shown in **Table 1**. Mean maternal BMI ranged from 23.0 to 28.5 kg/m^2^ across studies, and mean maternal age ranged from 25 to 31 years old. The maximum sample size from each study ranged from 582 (Gen3G) to 272,249 (UK Biobank). The number of cases ranged from 107 for miscarriage in the index pregnancy (used in multivariable regression and paternal negative control analyses) to 83,237 for miscarriage in previous pregnancies (used in Mendelian randomization analyses) (**Table 2**).

### Main analyses results

Results for the main multivariable regression (fully adjusted model) and Mendelian randomization (IVW) analyses are shown in **Figure 2** (binary outcomes) and **Supplementary figure 1** (continuous outcomes).

**Figure 2.**
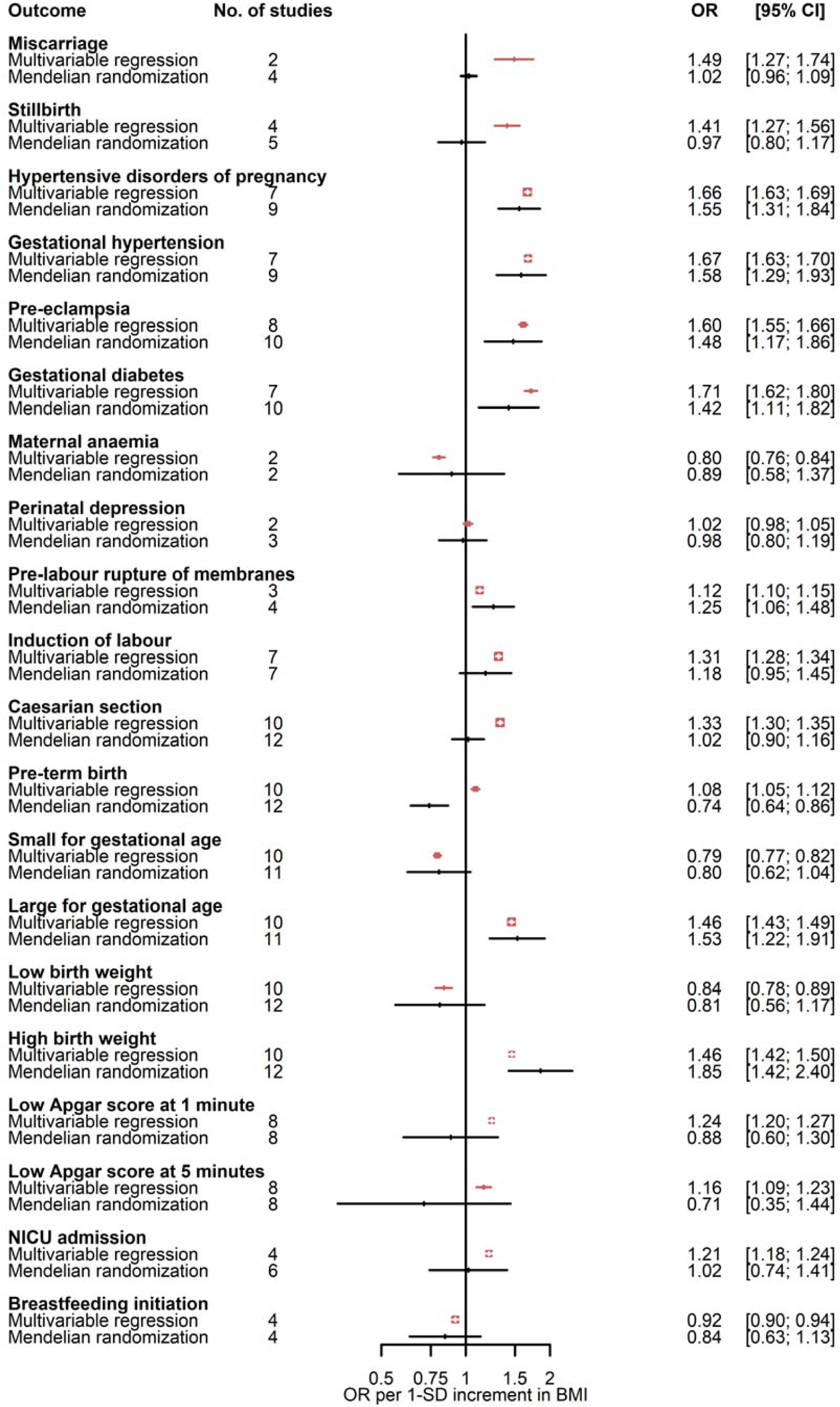
Comparison of adjusted multivariable and main Mendelian randomization estimates for associations of maternal body mass index with pregnancy and perinatal binary outcomes. BMI: body mass index; NICU: neonatal intensive care unit

In the main multivariable regression analyses, maternal BMI was associated with 19 out of the 20 binary outcomes. Higher maternal BMI was associated with higher risk of miscarriage, stillbirth, HDP, GH, PE, GDM, pre-labour membrane rupture, induction of labour, caesarean section, PTB, LGA, high birthweight, low Apgar score at 1 minute, low Apgar score at 5 minutes, and NICU admission. In addition, women with higher BMI were less likely to have maternal anaemia, have a baby SGA or with low birthweight, and initiate breastfeeding. There was little evidence of maternal BMI being associated with the risk of perinatal depression (**Figure 2**). Higher maternal BMI was associated with higher values of all continuous outcomes (i.e. birthweight, birth length, ponderal index, and gestational age) (**Supplementary figure 1**).

For the Mendelian randomization analyses, we estimated that the total R^2^ and mean F-statistic for the association of SNPs with BMI were 2.7% and 36, respectively, for the set of 97 SNPs using female-specific data from the GIANT GWAS. We observed a positive correlation (r = 0.67) between SNP-BMI estimates from females in the GIANT GWAS and SNP-BMI (pre-/early-pregnancy) estimates pooled across participating cohorts (**Supplementary figure 2**). In agreement with multivariable regression analyses, findings from Mendelian randomization indicated that higher maternal BMI is related to higher risk of HDP, GH, PE, GDM, pre-labour membrane rupture, induction of labour, LGA, high birthweight, lower risk of having baby SGA, and not associated with perinatal depression. On the other hand, in contrast with multivariable regression analyses, Mendelian randomization findings did not provide support for an association of maternal BMI with miscarriage, stillbirth and caesarean section, and indicated an inverse association of maternal BMI with PTB. As expected, given the lower statistical power, confidence intervals were wider for Mendelian randomization compared to multivariable regression analyses and included the null value for some of these outcomes (**Figure 2**). For other binary outcomes (i.e. low Apgar score at 1 and 5 minutes, NICU admission, maternal anaemia, low birthweight, and breastfeeding initiation), it was unclear whether estimates from multivariable and Mendelian randomization are in agreement given the substantial uncertainty in the latter. For continuous outcomes (i.e. birthweight, birth length, ponderal index, and gestational age), findings from Mendelian randomization indicated that higher maternal BMI was associated with higher values of continuous outcomes in agreement with multivariable regression analyses (**Supplementary figure 1**).

Paternal negative control results supported the role of maternal BMI on stillbirth, HDP, GH, PE, GDM, maternal anaemia, pre-labour membrane rupture, induction of labour, caesarean section, SGA, LGA, high birthweight, low Apgar score at 1 minute, NICU admission, and breastfeeding initiation (**Figure 3**). The association of paternal BMI with maternal perinatal depression was also close to the null, consistent with maternal multivariable and Mendelian randomization results. Associations with miscarriage, PTB, low birthweight and low Apgar score at 5 minutes were imprecise and/or more similar in direction and magnitude between paternal and maternal BMI analyses. Results for continuous outcomes were strongly attenuated for paternal BMI in relation to birthweight and length (**Supplementary figure 3**).

**Figure 3.**
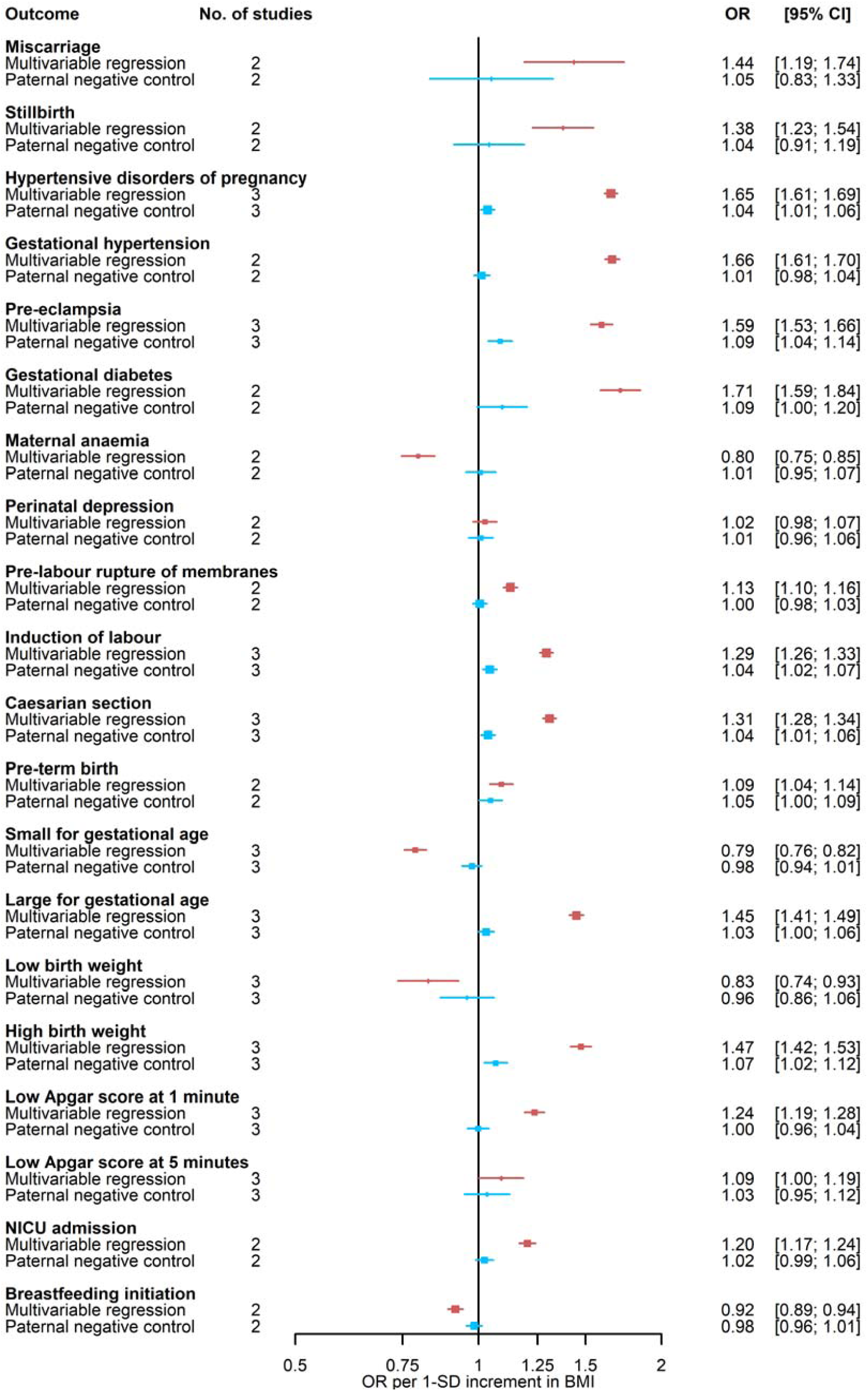
Comparison of maternal and paternal body mass index associations with adverse pregnancy and perinatal outcomes. Paternal BMI was used as a negative control exposure to explore the potential presence, direction and magnitude of bias in adjusted multivariable estimates for associations of maternal BMI with outcomes. Results are expressed as odds ratios per SD unit of maternal BMI and paternal BMI for ‘Multivariable regression’ and ‘Paternal negative control’, respectively. Multivariable regression results were adjusted for paternal BMI, maternal age, parity, education, smoking during pregnancy, alcohol use during pregnancy and offspring sex where available. Paternal negative control results were adjusted for maternal BMI, paternal age, number of children (ALSPAC only), paternal education, paternal smoking, paternal alcohol use and offspring sex. BMI: body mass index; NICU: neonatal intensive care unit

### Sensitivity analyses

Overall, findings from the main multivariable regression analyses were consistent across studies (**Supplementary figure 4**), when using random-effect metanalyses (**Supplementary figure 5**), and with minimally adjusted models (**Supplementary figure 6**). Between-study heterogeneity was substantial (i.e. Cochrane’s Q p-value < 0.05) for GDM, low Apgar score at 1 minute, gestational age and birthweight (**Supplementary table 4**).

The main Mendelian randomization analyses were not driven by any individual study as indicated by the leave-one-out analyses, with the exception of GDM, for which removing FinnGen resulted in attenuated estimates (**Supplementary figure 7**). Results were similar when using fixed- or random-effect metanalyses to pool SNP-outcome estimates across studies (**Supplementary figure 8**). There was evidence of substantial between SNP heterogeneity in the IVW analyses of maternal BMI with HDP, GH, PE, GDM, low Apgar score at 1 minute, breastfeeding initiation, and birthweight (**Supplementary table 5**). Despite that, there was no clear evidence of strongly influential outlying SNPs in the leave-one-out analyses (**Supplementary figure 9**) or directional pleiotropy as evidenced by the MR-Egger intercept test (except for PTB) (**Supplementary table 5**). Furthermore, Mendelian randomization results were generally consistent when using different Mendelian randomization methods (**Supplementary figure 10**), although estimates from MR-Egger were imprecise for some outcomes. Adjusting for offspring genotype did not substantially change effect estimates with a few exceptions, such as pre-labour rupture of membranes, LGA, and high birthweight, where adjusted results were attenuated (**Supplementary figure 11**).

Findings from the main paternal negative control analyses were consistent between studies (**Supplementary figure 12** for maternal associations additionally adjusted for partners BMI and **Supplementary figure 13** for paternal associations) and when comparing different models (**Supplementary figures 14-16**).

## DISCUSSION

By triangulating different analytical approaches, our findings are compatible with higher maternal BMI increasing the risk of HDP, GH, PE, GDM, pre-labour membrane rupture, induction of labour, LGA, and high birthweight, as well as decreasing the risk of SGA. In addition, we did not find supportive evidence for a relation of maternal pre- or early-pregnancy BMI with perinatal depression. For other outcomes, evidence is uncertain due to inconsistencies across multiple approaches (e.g. multivariable regression results for stillbirth, caesarean section, and PTB were not supported by Mendelian randomization) or substantial imprecision in effect estimates from Mendelian randomization (e.g. maternal anaemia and low Apgar score after 5 minutes).

Consistent with our results, a previous study using multivariable regression reported higher maternal BMI (across the whole distribution) was associated with increased risk of HDP, GDM and LGA, and reduced risk of SGA based on data from 265,270 mother-offspring pairs (samples partly overlapping with our study).^10^ In addition there was some evidence of a non-linear association with odds of PTB, which were higher in women who were underweight or obese.^10^ In agreement with these findings, a larger study (9,282,486 mother–infant pairs in the USA) focussed on offspring outcomes indicated that higher maternal BMI was associated with higher risk of high birthweight, LGA, as well as low Apgar score and reported a non-linear relationship with PTB risk.^36^ Other observational studies using multivariable regression have reported that maternal BMI is associated with higher risk of stillbirths^37^, induction^38^, caesarean section^38^, and not initiating breastfeeding.^39^ Previous Mendelian randomization studies have focused on a limited set of outcomes and are supportive of higher maternal BMI being related to higher mean offspring birthweight^4,40,41^ (N ∼ 9,000 to 400,000) and GDM^42^ (N = 5,485 cases and 347,856 controls).

Recent systematic reviews of randomised controlled trials (RCTs) of diet and physical activity during pregnancy (N range: 12,526-34,546) reported some evidence of reduced risk of GDM, LGA, caesarean section in those randomised to the intervention, but no effect or mixed results of the intervention on HDP, PTB, NICU admission.^43–45^ Of note, these studies aimed at managing weight gain during pregnancy rather than targeting weight reduction prior to pregnancy with a modest mean difference of −0.7 to −1.2 kg between women in the intervention compared to those randomised to standard care. In addition, evidence for many outcomes is uncertain due to the relatively small number of cases.

Although mechanisms are not fully understood, higher maternal BMI is likely to influence a range of processes that are involved in the aetiology of some of the outcomes of interest, such as insulin resistance, endothelial dysfunction, inflammation, and susceptibility to infection.^46^ In addition, maternal dysmetabolism resulting from excess adiposity has a well-recognised impact on maternal circulating nutrients, such as glucose, lipids and amino acids, some of which can cross the placenta and influence offspring outcomes, such as growth.^47–49^

### Strengths and Limitations

Key strengths of this study include exploring the potential role of maternal BMI on a wide-range of pregnancy and perinatal outcomes in large samples from multiple studies using different approaches. The credibility of findings from each approach relies on the plausibility of assumptions that are often not possible to verify, such as no unmeasured confounding in multivariable regression, similar confounding, selection and measurement error between paternal and maternal BMI analyses, and no confounding or horizontal pleiotropy in Mendelian randomization. Therefore, results in agreement across approaches strengthen the evidence on the relation of maternal BMI with the outcome. Where possible, we explored the plausibility of assumptions underlying each method. In particular, we conducted extensive sensitivity analyses to explore the plausibility of the core Mendelian randomization assumptions, and found overall these did not suggest Mendelian randomization results were driven by weak or invalid instruments. Furthermore, models adjusted by fetal genotype did not substantially change our estimates.

Key limitations of this study are as follows. First, despite the large scale of our study, statistical power varied across outcomes as some outcomes have lower prevalence and/or were not collected in all cohorts. Second, despite our efforts to capture the best and most homogeneous definition for outcomes across studies, this was not always possible as exemplified by GDM, for which the data collected was notably variable across studies (e.g. from self-report to medical records-derived information), and index miscarriage (which was used for multivariable regression and paternal negative control analyses but is poorly captured in birth cohorts during the early pregnancy period). Third, while we were interested in maternal pre-pregnancy BMI, only maternal weight reflecting early-/mid-pregnancy was available in four studies. Fourth, our analyses assumed a linear effects of BMI, which may not be the case for some outcomes like PTB, and were restricted to women of European ancestry given most studies had scarce data on women from other ancestries. While this reduces the risk of confounding by ethnicity or population structure, it may limit the generalizability to other populations of pregnant women.

### Clinical implications and conclusions

Our findings support a causal role for maternal pre-/early-pregnancy higher BMI on a range of adverse pregnancy and perinatal outcomes. Given the high prevalence of overweight and obesity our findings emphasise the need for development and testing of pre-conception interventions to support women maintaining a healthy BMI. This should be a key target to reduce the burden of obstetric and neonatal complications.

## Data Availability

In order to protect participant confidentiality, supporting data cannot be made openly available. Bona fide researchers can apply for access to study-specific executive committees. Summary association data for FinnGen is publicly available at https://www.finngen.fi/en/access_results. Researchers can apply for access to the UK Biobank data via the Access management System (AMS) (https://www.ukbiobank.ac.uk/enable-your-research/apply-for-access).

## FUNDING

The views expressed in this paper are those of the authors and do not necessarily reflect views of any funders, person or group listed in funding or acknowledgement statements.

This study was supported by the MRC Integrative Epidemiology Unit at the University of Bristol (MC_UU_00011/6), British Heart Foundation (AA/18/7/34219), the European Research Council under the European Union’s Seventh Framework Programme (FP/2007-2013) / ERC Grant Agreement (Grant number 669545), the European Union’s Horizon 2020 research and innovation programme under grant agreement No 733206 (LifeCycle), the US National Institutes of Health (R01 DK10324, U01 HG004415), the Bristol NIHR Biomedical Research Centre, and the Research Council of Norway through its Centres of Excellence funding scheme (project number 262700), and the Wellcome Trust [Grant number WT220390]. For the purpose of open access, the authors have applied a CC BY public copyright licence to any Author Accepted Manuscript version arising from this submission.

MCB has received supported from MRC Skills Development Fellowship (MR/P014054/1) and University of Bristol Vice-Chancellor’s Fellowship. DAL is a British Heart Foundation Chair (CH/F/20/90003) and NIHR Senior Investigator (NF-0616-10102). JT is supported by an Academy of Medical Sciences (AMS) Springboard Award, which is supported by the AMS, the Wellcome Trust, GCRF, the Government Department of Business, Energy and Industrial Strategy, the British Heart Foundation and Diabetes UK [SBF004\1079]. RMF was funded by a Wellcome Trust and Royal Society Sir Henry Dale Fellowship (WT104150), and is now funded by a Wellcome Trust Senior Research Fellowship (WT220390). RG received funding from the Dutch Heart Foundation (grant number 2017T013), the Dutch Diabetes Foundation (grant number 2017.81.002), and the Netherlands Organization for Health Research and Development (NWO, ZonMW, grant number 543003109). VWVJ received a Consolidator Grant from the European Research Council (ERC-2014-CoG-648916).. XL received support from the Nordic Center of Excellence in Health-Related e-Sciences. LS reports funding from a Carlsberg Foundation postdoctoral fellowship (CF15-0899). BF was supported by an Oak Foundation Fellowship and by a grant from the Novo Nordisk Foundation (12955). MCM has received funding from the European Research Council (ERC) under the European Union’s Horizon 2020 research and innovation programme (grant agreement number 947684). TAB is supported by the Medical Research Council (MRC) (UK) (MR/K501281/1), the NHMRC (Australia) (GNT1183074 and GNT1157714) and the British Heart Foundation Accelerator Award at the University of Bristol (AA/18/7/34219) and works in/is affiliated with a unit that is supported by the UK Medical Research Council (MC_UU_00011/6). LB is a senior research scholar from the Fonds de la recherche du Québec-Santé (FRQ-S) and a member of the FRQ-S-funded Centre de recherche du CHUS. MFH was supported by an American Diabetes Association (ADA) Pathways Accelerator Award (1-15-ACE-26). SEH and MCM are partly funded by the Research Council of Norway (project no. 320656, and through its Centres of Excellence funding scheme (project No 262700). JW and RMc are supported by the National Institute for Health and Care Research under its Applied Research Collaboration, Yorkshire and Humber (NIHR200166).

Study specific funding and acknowledgments are provided in Supplementary material.

We acknowledge Ville Karhunen for his support advising data analysis from NFBC1966 and NFBC1986.

