## Supplementary methods for "Integrating multiple lines of evidence to assess the effects of maternal BMI on pregnancy and perinatal outcomes in up to 497,932 women"

### INDIVIDUAL COHORT DETAILS

### ALSPAC

#### Study information

Pregnant women resident in Avon, UK with expected dates of delivery 1^st^ April 1991 to 31^st^ December 1992 were invited to take part in the study. The initial number of pregnancies enrolled is 14,541. Of these initial pregnancies, there was a total of 14,676 fetuses, resulting in 14,062 live births and 13,988 children who were alive a 1 year of age.^1,2^ Please note that the study website contains details of all the data that is available through a fully searchable data dictionary and variable search tool (<http://www.bristol.ac.uk/alspac/researchers/our-data>). Ethical approval for the study was obtained from the ALSPAC Ethics and Law Committee and the Local Research Ethics Committees (NHS Haydock REC: 10/H1010/70).

#### Study variables

Maternal body mass index (BMI) was calculated based on self-reported height and pre-pregnancy weight collected in questionnaire at 12 weeks of pregnancy. Information on miscarriages, stillbirths, and gestational diabetes were obtained from questionnaires during pregnancy and obstetric medical records. Data on preeclampsia, gestational hypertension, hypertensive disorder of pregnancy, maternal anaemia, offspring sex, birth weight, birth length, preterm birth, membrane rupture, mode of delivery, Apgar scores, and neonatal intensive care unit (NICU) admission were obtained from obstetric clinical records. Breast feeding information was collected in questionnaires 6 months and 15 months after birth. Information on parity, maternal education, maternal age, self-reported alcohol consumption, and smoking during pregnancy was collected in questionnaires during pregnancy.

#### Genotyping and imputation

ALSPAC children were genotyped using the Illumina HumanHap550 quad chip genotyping platforms by 23andme subcontracting the Wellcome Trust Sanger Institute, Cambridge, UK and the Laboratory Corporation of America, Burlington, NC, US. The resulting raw genome-wide data were subjected to standard quality control methods. Individuals were excluded on the basis of gender mismatches; minimal or excessive heterozygosity; disproportionate levels of individual missingness (>3%) and insufficient sample replication (IBD < 0.8). Population stratification was assessed by multidimensional scaling analysis and compared with Hapmap II (release 22) European descent (CEU), Han Chinese, Japanese and Yoruba reference populations; all individuals with non-European ancestry were removed. SNPs with a minor allele frequency of < 1%, a call rate of < 95% or evidence for violations of Hardy-Weinberg equilibrium (P < 5E-7) were removed. Cryptic relatedness was measured as proportion of identity by descent (IBD > 0.1). Related subjects that passed all other quality control thresholds were retained during subsequent phasing and imputation. 9,115 subjects and 500,527 SNPs passed these quality control filters.

ALSPAC mothers were genotyped using the Illumina human660W-quad array at Centre National de Génotypage (CNG) and genotypes were called with Illumina GenomeStudio. PLINK (v1.07) was used to carry out quality control measures on an initial set of 10,015 subjects and 557,124 directly genotyped SNPs. SNPs were removed if they displayed more than 5% missingness or a Hardy-Weinberg equilibrium P value of less than 1.0e-06. Additionally SNPs with a minor allele frequency of less than 1% were removed. Samples were excluded if they displayed more than 5% missingness, had indeterminate X chromosome heterozygosity or extreme autosomal heterozygosity. Samples showing evidence of population stratification were identified by multidimensional scaling of genome-wide identity by state pairwise distances using the four HapMap populations as a reference, and then excluded. Cryptic relatedness was assessed using a IBD estimate of more than 0.125 which is expected to correspond to roughly 12.5% alleles shared IBD or a relatedness at the first cousin level. Related subjects that passed all other quality control thresholds were retained during subsequent phasing and imputation. 9,048 subjects and 526,688 SNPs passed these quality control filters.

We combined 477,482 SNP genotypes in common between the sample of mothers and sample of children. We removed SNPs with genotype missingness above 1% due to poor quality (11,396 SNPs removed) and removed a further 321 subjects due to potential ID mismatches. This resulted in a dataset of 17,842 subjects containing 6,305 duos and 465,740 SNPs (112 were removed during liftover and 234 were out of HWE after combination). We estimated haplotypes using ShapeIT (v2.r644) which utilises relatedness during phasing. We obtained a phased version of the 1000 genomes reference panel (Phase 1, Version 3) from the Impute2 reference data repository (phased using ShapeIt v2.r644, haplotype release date Dec 2013). Imputation of the target data was performed using Impute V2.2.2 against the reference panel (all polymorphic SNPs excluding singletons), using all 2186 reference haplotypes (including non-Europeans).

This gave 8,237 eligible children and 8,196 eligible mothers with available genotype data after exclusion of related subjects using cryptic relatedness measures described previously.

### BiB

#### Study information

The Born in Bradford (BiB) study is a population-based prospective birth cohort. In total, 12,453 women who had 13,776 pregnancies were recruited at ~24-28 weeks gestation at a routine oral glucose tolerance test (OGTT). At the time of recruitment, this was offered to all women booked for delivery at Bradford Royal Infirmary (BRI) (with the exception of those with pre-existing diabetes (N = 70 - 0.5% of BiB pregnancies)).^3^ All women recruited had an expected delivery between March 2007 and December 2010. Full details of the study methodology were reported previously.^4^ In brief, Bradford is a city in the North of England with high levels of socioeconomic deprivation. BiB has a high proportions of White European and South Asian families, all residing in Bradford, UK, which makes it a unique cohort demographic to study. The [study website](https://borninbradford.nhs.uk/research/documents-data/) provides more information, including protocols, questionnaires and information on how researchers can access data and a full list of all available data. Parents (usually the mother) who were recruited into the study provided full informed consent, as well as detailed interview questionnaire data, measurements, and biological samples. They also consented to the linkage of their and their child’s data to routine (primary and secondary care) health and education data. Ethical approval for the study was granted by the Bradford National Health Service Research Ethics Committee (ref 06/Q1202/48).

#### Study variables

Data on weight were extracted from the first antenatal clinic records (around 12 weeks of gestation). Weight (kg, Seca 2 in1 scales, Harlow Healthcare Ltd., London, UK) and height (cm) were measured using established protocols at recruitment and used to calculate BMI. Information on miscarriages, stillbirths, and gestational diabetes, preeclampsia, gestational hypertension, hypertensive disorder of pregnancy, maternal anaemia, offspring sex, birth weight, birth length, preterm birth, membrane rupture, mode of delivery, Apgar scores, and neonatal intensive care unit (NICU) admission were obtained from various sources including questionnaires and obstetric clinical records. Information on parity, maternal education, maternal age, self-reported alcohol consumption, and smoking during pregnancy was collected in questionnaires during pregnancy.

#### Genotyping and imputing

In BiB, genotype data are available on 16,267 mothers and children. These were genotyped at Bristol Bioresource laboratories on 3 different HumanCoreExome arrays - Illumina HumanCoreExome12v1.0, HumanCoreExome12v1.1 and HumanCoreExome24v1.0 arrays and genotypes were called with Illumina GenomeStudio. Any individual or SNP missing >3% of their data was dropped and the 3 datasets combined. This resulted in 513,889 genetic variants genotyped in all 15,862 individuals. Array genotypes were harmonized, phased and imputed by the Sanger Imputation Service using the "UK10K + 1000 Genomes Phase 3 reference panel" and the "pre-phase with EAGLE2 and impute" pipelines.^5^ Other QC metrics typically used on genotype data, such as deviation from Hardy-Weinberg Equilibrium and excess homozygosity, are inappropriate here, given the ethnicity split, population structure and high levels of inbreeding and consanguinity present in this cohort.

### DNBC-GOYA

#### Study information

The GOYA (Female) sample were derived from the Danish genome-wide association study GOYA (Genomics of extremely Overweight Young Adults), nested within the Danish National Birth Cohort (DNBC).^6-8^ The DNBC is a collection of data on 92,274 pregnant women recruited between 1996 and 2002, from their first antenatal visit to their general practitioner. Women participated in four telephone interviews (16 and 30 weeks gestation and 6 and 18 months after birth). They also provided two blood samples during pregnancy. The GOYA females were drawn from the 67,863 women within the DNBC who provided information about pre-pregnancy BMI, gave birth to a live born infant and provided a blood sample during pregnancy. A case sample of the 3.6% most obese women (n=2451) was defined as those with the largest residuals from the regression of BMI on age and parity (all entered as continuous variables). The BMI for these women ranged from 32.6 to 64.4. From the remaining cohort we selected a random sample of similar size (n=2450). Information about pregnancy outcomes derived from Danish health registers. The study was approved by the regional scientific ethics committee and by the Danish Data Protection Board.

#### Study variables

Maternal BMI was calculated based on self-reported height and pre-pregnancy weight collected in questionnaire surveys during pregnancy (average: 16 weeks of gestation). Self-reported alcohol consumption, smoking information was collected in questionnaires during pregnancy and by computer assisted telephone interview 6 months after birth. We obtained data on offspring sex, parity, maternal age at birth, gestational age, length and weight at birth, and Apgar scores from the Danish Medical Birth Register^9^. Preeclampsia was defined by the ICD10 codes O14 and O15 based on data from the National Patient Register. Gestational diabetes was defined by the ICD10 code O24 based on data from the National Patient Register and by self-reported information about this condition from the pregnancy interview.

#### Genotyping and imputation

Maternal genome-wide data were obtained from the genome-wide Illumina Human 610-Quad v1.0 BeadChip (545,349 SNPs). Of the initial 4901 women, 1960 extremely overweight cases and 1,948 control women were genotyped and passed quality control. Data were imputed to HapMap release 22, as previously described.^6-8^

### DNBC-PTB

#### Study information

The Danish National Birth Cohort - Preterm Birth Study (DNBC-PTB) is a case-control study using mother-infant pairs to investigate genetic and environmental influences on spontaneous preterm birth^10^. The study is nested within the DBNC^11^; for case pairs delivery occurred before 37 weeks of gestation and for control pairs delivery occurred in gestational week 39, 40, or 41. Exclusion criteria were multiple deliveries, pregnancy complications such as placental abnormalities, preeclampsia/eclampsia, congenital abnormalities or stillbirth. Individuals were further required to be of Northern European ancestry. Ethical approval was obtained from the Regional Scientific Ethical Committee of Copenhagen and the study was also approved by the Danish Data Protection Agency.

#### Study variables

Maternal BMI was calculated based on self-reported height and pre-pregnancy weight collected in questionnaire surveys during pregnancy (average: 16 weeks of gestation). Self-reported alcohol consumption and smoking information was collected in questionnaires during pregnancy and by computer assisted telephone interview 6 months after birth. Breast feeding information was also collected by telephone interviews 6 months and 18 months after birth. We obtained data on offspring sex, parity, maternal age at birth, gestational age, length and weight at birth, membrane rupture before onset of contractions, mode of delivery and Apgar scores from the Danish Medical Birth Register^9^.

#### Genotyping and imputation

Genome-wide genotype data were obtained using the Illumina Human 660W Quad array and generated as part of the Gene Environment Association Studies (GENEVA) consortium. Data cleaning and QC steps were based on the following requirements for samples and SNPs to be included: sample missingness rate < 4%, heterozygosity within 3 standard deviations from the mean, variant missingness rate < 2%, minor allele frequency > 1%, A/T and C/G variants excluded, Hardy-Weinberg P value > 1e-06. The GWAS data were imputed with SHAPEIT and IMPUTE2 using the 1000 Genomes phase I data as reference set.

### EFSOCH

#### Study information

The EFSOCH study^12^ is a prospective study of children born between 2000 and 2004, and their parents, from a geographically defined region of Exeter, UK. All women gave informed consent and ethical approval was obtained from the local review committee.

#### Study variables

Maternal body mass index was calculated from weight (pre-pregnancy weight was self-reported) and height, measured using standard protocol at the study visit. Information on maternal smoking and occupation was collected by questionnaire. Information on maternal education and alcohol use during pregnancy were not available at the time this analysis was carried out. Pregnancy details were obtained from the Exeter Maternity Unit Database. Gestational age at delivery was calculated based on last menstrual period, or when that was unreliable or unavailable, dating by ultrasound scan was used.  Information offspring sex, and whether the birth was by Caesarean section was recorded at delivery by research midwives. Birth weight was recorded in grams to the nearest 10g, using Soehnle (Leifheit AG, Nassau, Germany) scales (model number 8310, range up to 20kg×10g). Birth length was measured in centimetres to the nearest 0.1cm, using the Harpenden infantometer (range 120cm×0.1cm). Measures were repeated three times and an average value was calculated for use in analyses.

#### Genotyping and imputation

Maternal and paternal DNA samples were extracted from parental blood samples obtained at the study visit (when the women were 28 weeks pregnant), and offspring DNA was obtained from cord blood at birth.

Genotyping of 2768 EFSOCH samples (n=969 mothers, 937 fathers and 862 children) was performed using the Illumina Infinium HumanCoreExome-24 array (n=551,839 SNPs/indels). Individual DNA samples with genotype call rate <98% were removed (n=50 individuals [1.8%]). SNPs were removed if they had call rates <95% (n=13,151 SNPs), showed evidence of deviation from Hardy-Weinberg equilibrium (P<1x10^-6^; n=455 further SNPs), or had a minor allele frequency (MAF) <1% (n=257,289 further SNPs). Genotypically-derived sex information was compared with sex information in the phenotype file, and mismatched samples were excluded (n=13 individuals [0.61%]). Kinship was estimated using King.^13^ Where evidence of labelling errors was clear, labels were updated, otherwise samples with kinship errors were excluded (n=22 individuals [0.79%]). Principal component analysis was performed to assess ancestry of the sample using flashPCA.^14^ Outliers were defined as >4.56 SD from the cluster mean (defined using 1000 Genomes European data as the reference) and excluded (n=21 individuals [0.76%]). Imputation was performed using the Michigan imputation server and samples were imputed to the Haplotype Reference Consortium HRC v1.1 reference panel. SNPs were included in the analysis if imputation quality score was >0.4 and MAF ≥1%. After imputation, and once all exclusions had been made, 7,674,771 SNPs were available for analysis in 2662 individuals (939 mothers, 911 fathers and 812 children). Analyses were adjusted for genotyping batch.

### FinnGen

#### Study information

FinnGen is the national wide network of Finnish biobanks (including Auria Biobank, Biobank Borealis of Northern Finland, Biobank of Eastern Finland, Central Finland Biobank, Finnish Red Cross Blood Service Biobank, Finish Clinical Biobank Tampere, Helsinki Biobank, Terveystalo Biobank and THL Biobank).^15^ Further information available at <https://finngen.gitbook.io/documentation/v/r5/>. These biobanks were linked to national registries, including Drug purchase and Drug Reimbursement, Digital and Population Data Services Agency, Statistics Finland, Register of primary health care visits: AVOHILMO, Care Register for Health Care: HILMO, and Finnish cancer registry. The Coordinating Ethics Committee of the Helsinki and Uusimaa Hospital District has approved the FinnGen consortium (Nr HUS/990/2017), and the ethical approval of each individual study has been described in detail elsewhere.^16^

#### Study variables

The FinnGen team released summary-level data of GWAS of 2803 clinical endpoints in 218,792 men and women at their R5 wave (available at <https://r5.finngen.fi/>). This study included 8 endpoints: miscarriage, HDP, gestational hypertension, preeclampsia, gestational diabetes, membrane rupture before onset of contractions, caesarean section, and preterm birth, which were defined based in ICD-10 codes (or the equivalent in ICD-9). For miscarriage, cases were women having records of O03 Spontaneous abortion, and controls were those without records of O00-O08 Pregnancy with abortive outcome. For HDP, cases were women having records of O10 Pre-existing hypertension complicating pregnancy, childbirth and the puerperium, O11 Pre-existing hypertensive disorder with superimposed proteinuria, O13 Gestational [pregnancy-induced] hypertension, O14 Gestational [pregnancy-induced] hypertension with significant proteinuria, O15 Eclampsia or O16 Unspecified maternal hypertension, and controls were those without any of these records. For gestational hypertension, cases were women having records of O13, and controls were those without records of O10-O16 Oedema, proteinuria and hypertensive disorders in pregnancy, childbirth and the puerperium. For preeclampsia, cases were women having records of O14, and controls were those without records of O10-16. For gestational diabetes, cases were women having records of O24.4 Diabetes mellitus arising in pregnancy, and controls were the rest. For membrane rupture before onset of contractions, cases were women having records of O42 Premature rupture of membranes, and controls were those without records of O30-O48 Maternal care related to the fetus and amniotic cavity and possible delivery problems. For caesarean section, cases were women having records of O82 Single delivery by caesarean section, controls were those without records of O80-O84 Delivery. For preterm birth, cases were women having records of O60 Preterm labour and delivery, and controls were those without records of O60-O75 Complications of labour and delivery.

#### Genotyping and imputation

FinnGen participants were genotyped with Illumina and Affymetrix chip arrays. Genotype data were imputed against SISu v3 reference panel (described at <https://finngen.gitbook.io/documentation/methods/genotype-imputation/sisu-reference-panel>), after a quality control procedure (i.e. minor allele count ≥3, a call rate ≥98%, in HWE no outliers in heterozygosity, correct sex assignment, and of Finnish ancestry).

### Gen3G

#### Study information

We initiated the Genetics of Glucose regulation in Gestation and Growth (Gen3G) prospective cohort to increase our understanding of biological, environmental and genetic determinants of glucose regulation during pregnancy and their impact on fetal development. Between January 2010 and June 2013, we invited pregnant women aged ≥ 18 years old who visited the blood sampling in pregnancy clinic in Sherbrooke for their first trimester clinical blood samples. Ethics approval: Centre Hospitalier Universitaire de Sherbrooke (CHUS) Ethics Review Board for Studies with Humans.

#### Study variables

Gen3G women classified as having pre-eclampsia (PE) if they had gestational hypertension (at least two BP measures ≥ 140/90 at least 20 minutes apart, from the 20^th^ week of gestation to 6 weeks after delivery) with proteinuria (≥ 300 mg/day or protein/creatinine ratio ≥ 0.3), as per classic definition^17^ and as having gestational hypertension (GH) if they had hypertension without proteinuria during pregnancy or up to 6 weeks after delivery. All cases were retrospectively reviewed and discussed with a nephrology fellow to ensure correct classification based on presence of hypertension and proteinuria. HDP defined as the combination of both women with preeclampsia and gestational hypertension.

Gen3G women were classified as having gestational diabetes based on the results of a routine GDM screening in the second trimester that was scheduled between 24 and 30 weeks of gestation. GDM status was determined according to IADPSG criteria, where GDM is diagnosed if the fasting glucose is above 92 mg/dl (5.1 mmol/l), the 1 hour plasma glucose is greater than 180 mg/dl (10 mmol/l) or the 2 hour plasma glucose is greater than 153 mg/dl (8.5 mmol/l). Details of delivery including induction of labour and type of labour were extracted from medical records. Details about the offspring, including whether the birth was preterm, gestational age at birth, birthweight, birth length, one and five minute APGAR scores, and whether the offspring exhibited fetal macrosomia were also obtained from medical records. Low birthweight, small for gestational age and large for gestational age were determined using the Fenton chart.^18^

#### Genotyping and imputation

Samples were genotyped using the Illumina Infinium Expanded Multi-Ethnic Genotyping (MEGA EX) genotyping array. From a total of 2,036,065 genotyped SNPs, we removed SNPs with a call rate less than 95% (n=140,897), SNPs with a Hardy-Weinberg equilibrium P less than 1x10e-8 (n=718508), SNPs where concordance between duplicates was less than 90% (n=37). At the individual level, we removed duplicates as well as a sample where duplicates were not similar (n=27), individuals with a call rate <= 98% (n=15). We also performed checks of heterozygosity and for outliers via PCA, which did not result in any further exclusions. To prepare for imputation after the sample-based QC we removed monomorphisms (n=38,920), indels (n=738) and chrXY and chr 0 (n=9,283), SNPs with MAF < 0.01 (n=269,306) and finally removed the duplicate with the smaller call rate (n=19491), and one SNP with a wrong reference allele for a final total of 838,884 SNPs. Finally, we performed imputation on the Michigan Imputation server (Minimac3) with the 1000G reference panel.

### Generation R

#### Study information

The Generation R Study is a population-based prospective cohort study from early pregnancy onwards.^19^ All pregnant women living in Rotterdam, the Netherlands with a delivery date between April 2002 and January 2006 were eligible to participate. The cohort includes 9,778 mothers and their 9,749 live-born children. The vast majority of these, 8,880 women or 91%, were enrolled during pregnancy. Of these, 8,633 had singleton live births. The study protocol was approved by the Medical Ethical Committee of the Erasmus MC, University Medical Center Rotterdam and informed consent was obtained for all participants.

*Study variables*

Maternal body mass index was calculated from weight and height measured using a standardized protocol at the baseline study visit. For our analyses, we excluded those women for whom body mass index was measured at or after 20 weeks of gestation. We obtained data on offspring sex, gestational age, length and weight at birth, induction, mode of delivery and Apgar scores from medical (midwife or hospital) records. We used questionnaires at enrolment in the study to collect information about maternal age, educational level, parity, smoking, alcohol usage.^19^ Information about gestational hypertensive disorders (gestational hypertension, preeclampsia) was available from medical records. ^20^

#### Genotyping and imputation

Custom genotyping of the 32 SNPs identified in a large-scale GWAS meta-analysis for BMI was carried out by LGC Genomics using a Taqman allelic discrimination assay (Applied Biosystems, Foster City, CA) and Abgene QPCR ROX mix (Abgene, Hamburg, Germany). Call rate was 99.3% and duplicate concordance was 99.8%. All considered SNPs found to be in HWE, with the exception of rs4836133 (ZNF608;p= 3*10^−15^). To confirm the accuracy of the genotyping results, 276 randomly selected samples were genotyped for a second time using the same method, with an error rate of <1%.

### HAPO

#### Study information

The Hyperglycemia and Adverse Pregnancy Outcomes (HAPO) Study was a multicenter, international study that collected high-quality phenotypic data related to fetal growth and maternal glucose metabolism from 25,000 pregnant women of varied geographic, ethnic, and sociodemographic backgrounds. All pregnant women at less than 32 weeks of gestation were eligible for enrollment in HAPO unless they met one of several exclusion criteria published elsewhere.^21,22^ The protocol was approved by the institutional review board at each field center. All participants gave written informed consent. An external data and safety monitoring committee provided oversight.

#### Study variables

Participants underwent a 75-g oral glucose tolerance test (OGTT) at 28 weeks’ gestation. Maternal DNA was taken from blood collected into an EDTA tube at 2 h during the OGTT, when phenotypes of interest were measured, including glucose, blood pressure, weight, and height. Glucose and C-peptide were measured in a central laboratory. Newborn anthropometric traits, including the birth length, head circumference, birth weight, percent fat mass and sum of skinfolds, as well as newborn metabolic traits, including cord glucose and C-peptide were collected within 72 hours of birth. Study phenotype collection methods have been published elsewhere.^21,22^

#### Genotyping and imputation

Genotype data that passed initial QC at the genotyping centers were released to the GENEVA Coordinating Center (CC), National Center for Biotechnology Information database of Genotypes and Phenotypes (dbGaP), and HAPO study teams, who collectively performed QC using procedures previously described by the GENEVA consortium.^23^ Poorly performing samples or SNPs were removed based on misspecified sex, chromosomal anomalies, unintended sample duplicates, sample relatedness, low call rate, high number of Mendelian errors, departures from Hardy- Weinberg equilibrium, duplicate discordance, sex differences in heterozygosity, and low minor allele frequencies. Complete QC reports are available through dbGaP, <http://www.ncbi.nlm.nih.gov/projects/gap/cgi-bin/study.cgi?study_id=phs000096.v4.p1>. Samples were phased using ShapeIT. Imputation was performed using the Michigan imputation server (https://imputationserver.sph.umich.edu/index.html) and samples were imputed to the 1000G Phase 3 v5 reference panel.

### INMA

#### Study information

Population-based birth cohorts were established as part of the INMA – INfancia y Medio Ambiente [Environment and Childhood] Project in several regions of Spain following a common protocol. This project aims to study the associations between pre- and postnatal environmental exposures and growth, health, and development from early foetal life until adolescence and has been described previously in detail.^24^ Pregnant women were enrolled during the 1st trimester of pregnancy at public primary health care centers or public hospitals. Detailed measurements were performed using ultrasound and physical examinations and biological samples were collected. Informed consent was obtained from all participants and the study was approved by the Hospital Ethics Committees in each participating region.
This particular analysis uses the INMA cohorts of Valencia (VAL), Sabadell (SAB), and Gipuzkoa (GIP). Analyses were restricted to mother-child pairs of European ancestry, with data on pre-pregnancy maternal body mass index (BMI), maternal age, child’s sex and BMI genetic variants.

#### Study variables

Maternal pre-pregnancy BMI was calculated from self-reported weight and height measured at the first trimester. Covariates included offspring’s sex, maternal age at birth, maternal education, tobacco smoking during pregnancy, alcohol intake during pregnancy and parity assessed through questionnaires at the first or third trimester. The following outcomes were investigated: offspring birthweight measured by trained personnel at birth and birthlength also measured by trained personnel during the next 12h, and derived measurements (ponderal index, low birth weight, macrosomia); gestational age was established on the basis of date of the last menstrual period (LMP) self-reported by the mothers, however, in the case of discrepancy between LMP and an early ultrasound measurement of >=7 days, the latter was used, and derived measurements (preterm birth, small for gestational age, large for gestational age); type of delivery, either vaginal or caesarean; low Apgar score at time 1 minute; gestational diabetes, assessed through impaired glucose tolerance (IGT) (controls had a negative IGT or no test); and ever breastfeeding, assessed at around age 1 year.

#### Genotyping and imputation

Maternal DNA was obtained from blood collected during pregnancy using the Chemagen protocol at the Spanish National Genotyping Centre (CEGEN) (Spain) (SAB and VAL subcohorts) and using an automated DNA extraction method by the AutoGenFlexStar machine and the Flexigen DNA Kit (Qiagen). Genotyping of 32 BMI-associated SNPs and 6 additional proxies was performed using the KASP^TM^ technology at the LGCgroup (UK). Genotypes were flipped to + strand, if required, and missing genotypes were filled with genotypes from proxy SNPs, if available. Mothers with missing data in at least 1 of the 32 SNPs (or their proxies) were excluded from the analysis.

Offspring DNA was obtained from cord blood collected at birth or child’s blood collected at the age of 4y as described for the mothers. Genome-wide genotyping was performed using the Illumina HumanOmni1-Quad Beadchip at CEGEN (SAB and VAL subcohorts) and the Illumina GSA Beadchip at the Human Genotyping Facility (HuGeF) (the Netherlands) (GIP subcohort). Genotype calling was done using the GeneTrain2.0 algorithm based on HapMap clusters implemented in the GenomeStudio software. Quality control was done using PLINK and following standard criteria. First of all, SNPs were flipped to the human genome + strand. We applied the following initial quality control thresholds: sample call rate>98% and/or LRR SD<0.3. Then, we checked sex, relatedness, heterozygosity and population stratification. Genetic variants were filtered for SNP call rate>95%, MAF>1% and HWE P-value> 1.10E-06. Imputation of genetic variants was done using IMPUTE V2 and the cosmopolitan 1000 genome panel (release March 2012) (SAB and VAL) and the Michigan imputation server using the Haplotype Reference Consortium HRC v1.1 reference panel (GIP).

### MoBa

#### Study information

The Norwegian Mother, Father and Child Cohort Study (MoBa) is a population-based pregnancy cohort study conducted by the Norwegian Institute of Public Health. Participants were recruited from all over Norway from 1999-2008. The women consented to participation in 41% of the pregnancies. The cohort includes approximately 114.500 children, 95.200 mothers and 75.200 fathers. The current study is based on version 12 of the quality-assured data files released for research in 2019. The establishment of MoBa and initial data collection was based on a license from the Norwegian Data Protection Agency and approval from The Regional Committees for Medical and Health Research Ethics. The MoBa cohort is currently regulated by the Norwegian Health Registry Act. The current study was approved by The Regional Committees for Medical and Health Research Ethics of South/East Norway (ref 2018/1256). The Medical Birth Registry (MBRN) is a national health registry containing information about all births in Norway. Blood samples were obtained from both parents during pregnancy and from mothers and children (umbilical cord) at birth.

#### Study variables

In MoBa, information on maternal (and paternal) age at delivery was available from the Medical Birth Registry of Norway (MBRN). Maternal ethnic origin was based on mothers place of birth from the MBRN data and collapsed into: white (High income, Norway or Central or Eastern Europe or Central Asia) and non-white (Africa, Asia or Latin America). Information on number of pregnancies (parity) was also taken from the MBRN and collapsed to least one pregnancy (1) or no pregnancies (0). Maternal and paternal BMI were based on self report through a questionnaire at around 15 weeks and derived from weight and height. Maternal and paternal education were also self reported at around 15 weeks and coded as low (9 year elementary education or further education 1-2 years), medium (Further education, vocational or 3+ years) and high (Higher education, university or college). Maternal smoking was defined as any smoking during pregnancy and based on the 30 week questionnaire which asked about smoking from week 0 – week 30. Paternal smoking was defined as any smoking, based on the father’s questionnaire at 15 weeks. Maternal alcohol was defined as any alcohol during pregnancy and based on the 30 week questionnaire which asked about alcohol intake from week 0 – week 30. Paternal alcohol intake was defined as alcohol at least once a week, based on the fathers questionnaire at 15 weeks. For the miscarriage and stillbirth, confounders were derived based solely on the MBRN or first questionnaire at week 15.

#### Genotyping and imputation

Genotyping of the samples was performed in seven different batches on different Illumina platforms over a period of four years at deCODE genetics, Reykjavik, Iceland (HumanCoreExome-12 v.1.1, HumanCoreExome-24 v.1.0, Global Screening Array v.1.0, HumanOmniExpress-24-v1.0, InfiniumOmniExpress-24v1.2, and GSA24-v1.0). The Genome Reference Consortium Human Build 37 (GRCh37) reference genome was used for all annotations. Genotypes were called in Illumina GenomeStudio v.2011.1 for the HARVEST substudy (11,490 triads) and v.2.0.3 for the remaining batches. Cluster positions were identified from samples with call rate ≥ 0.98 and GenCall score ≥ 0.15.

Pre-imputation QC was performed for each subpopulation on the SNP, individual, and family level. Subpopulations were defined using principal component (PC) analyses using 1000 Genomes phase 1 data to identify the European, Asian, and African core subpopulations (after removing SNPs with MAF < 1%, call rate < 95%, and HWE p-value < 0.001). The primary softwares used for the QC were PLINK 1.9 and KING 2.2.5. For the SNP-level QC, SNPs were removed if MAF < 0.5%, call rate < 95%, HWE p-value < 0.000001, discordant in duplicate pairs, associated with genotype plate and genotype batch at p-value 0.001. For individual-level QC, individuals were removed if heterozygosity outliers Fhet ± 0.2, erroneous sex assignment, known relatedness, cryptic relatedness, identity-by-decent (PI_HAT threshold of 0.15), and PC outliers both with and without 1000 Genomes. For family-level QC, families with more than 5% Mendel errors and SNPs with more than 1% of Mendel errors were removed, while other minor Mendel errors were zeroed out. Batches that were genotyped using the same array were merged (keeping only SNPs present in all batches) and the pre-imputation QC was performed on the merged batches.

Phasing and imputation was performed using the publicly available Haplotype Reference Consortium data. Phasing was performed using SHAPEIT2 with the duoHMM algorithm to incorporate the pedigree information into the haplotype estimates. IMPUTE 4 was then used to perform imputation. Dosage data was then converted to best-guess, hard call genotype data with an imputation quality score (INFO) of 0.8 and default PLINK certainty of 0.9.

Post-imputation QC was then performed following the steps outlined in the pre-imputation QC. To ensure the across batch relatedness (both known and unknown) was accounted for in all analyses the three imputation batches were merged, and post-imputation QC was performed on the overall merged dataset.

### The Northern Finland Birth Cohorts1966 and 1986 (NFBC1966 and 1986)

#### Study information

NFBC1966: The Northern Finland Birth Cohort 1966 is a prospective follow-up study of children from the two northernmost provinces of Finland.^43^ 96% of all woman in this region with expected delivery dates in 1966 were recruited though maternity health Centres (12,058 live births). All individuals still living in northern Finland or the Helsinki area (n = 8,463) were contacted and invited for clinical examination. A total of 6007 participants attended the clinical examination at the participants’ age of 31 years. DNA was extracted from blood samples given at the clinical examination (5,753 samples available)^44^.The subset with DNA is representative of the original cohort in terms the major environmental and social factors known to influence the tested trait. An informed consent for the use of the data including DNA was obtained from all subjects. The current study includes a subset of female offspring participants who gave birth during the course of the study. NFBC1966 received ethical approval from Ethics Committee of Northern Ostrobothnia Hospital District (EETTMK: 94/2011) and Oulu University, Faculty of Medicine, Oulu, Finland.

NFBC1986: The Northern Finland Birth Cohort 1986 consists of 99% of all children, who were born in the provinces of Oulu and Lapland in Northern Finland between 1 July 1985 and 30 June 1986. 9,203 live-born individuals entered the study^1^. At the age of 16, the subjects living in the original target area or in the capital area (n=9,215) were invited to participate in a follow-up study including a clinical examination. 7344 participants attend the study in year 2001/2002, of which 5654 completed the postal questionnaire, the clinical examination and provided a blood sample^2^. DNA was extracted from all 5654 blood samples. The current study includes a subset of female offspring participants who gave birth during the course of the study. NFBC1986 received ethical approval from Ethics Committee of Northern Ostrobothnia Hospital District (EETTMK: 108/ 2017) and Oulu University, Faculty of Medicine, Oulu, Finland.

#### Study variables

All study variables, were collected from the maternal birth registers of the females offspring of the NFBC1966 and NFBC1986 at the time of the study. The data is prospectively collected from all women giving birth in Finland and included in the birth registry. The measures are collected during interviews, clinical examination and diagnosed (when relevant) by trained midwife and obstetricians. The birth registers included the following variables: miscarriage, gestational age (LMP and scan confirmed, in weeks), stillbirth, birth weight (in g), birth length (in cm), mode of delivery, APGAR score, and sex. Questionnaires include: parity, maternal smoking, level of maternal education, pre-pregnancy BMI of the mother was self-reported by the mother. The following diagnoses were included when relevant: gestational hypertension, hypertensive disorders, pre-eclampsia, gestational diabetes, preterm birth, NICU admission, induction of labour, macrosomia.

#### Genotyping and imputation

| Study acronym | Population group | Country of origin | Genotyping array(s) |
| --- | --- | --- | --- |
| NFBC1966 | Finnish | Finland | Illumina HumanCNV370DUO Analysis BeadChip |
| NFBC1986 | Finnish | Finland | Human Omni Express Exome 8v1.2 |

| Study acronym | Sample QC | SNV QC | | Imputation | | |
| --- | --- | --- | --- | --- | --- | --- |
|  | Call rate | Call rate | HWE p-value | Reference panel | Software | QC |
| NFBC1966 | >95% | >95% | >1×10^-4^ | HRC | IMPUTE2 | info > 0.4 |
| NFBC1986 | >95% | >95% | >1×10^-4^ | HRC | IMPUTE2 | info > 0.4 |

### UK Biobank

#### Study information

Between 2006-2010 all people in the UK National Health Service (NHS) registry aged between 40-69 years and living within approximately 25-mile radius from one of the 22 study centres were invited to participate in UK Biobank (UKB) (PMID: 25826379; PMID: 28531320). A total of 503,325 adults (5.5% of the ~9.2 million invited) were recruited into the study. A wide range of information was assessed at baseline via a self-completed touch-screen questionnaire, a brief computer-assisted interview, physical and functional measures, and collection of blood, urine, and saliva. Hospital inpatient data (Hospital Episode Statistics (HES)) are available for the full cohort and date back to 1997 for England, 1998 for Wales and 1981 for Scotland and contain coded data on admissions, operations and procedures (https://biobank.ctsu.ox.ac.uk/crystal/crystal/docs/HospitalEpisodeStatistics.pdf). HES data also contain maternity-related admissions for England and Wales. UKB participants who had a valid email address (N=339,229) were invited to fill in a detailed online questionnaire assessing their mental health after January 2015, and 158,835 participants fully completed it by October 2017 (https://biobank.ctsu.ox.ac.uk/showcase/ukb/docs/mental_health_online.pdf).

Ethical approval for UKB was obtained from the North West Multi-centre Research Ethics Committee (MREC), and our study was performed under UKB application number 23938.

#### Study variables

Self-reported and HES data were used to derive information on miscarriage, stillbirth, hypertensive disorder of pregnancy, gestational hypertension, preeclampsia, and gestational diabetes. Perinatal depression was based on the self-reported online mental health questionnaire applied to a subsample of participants. Birth-related outcomes (caesarean section, induction of labour, preterm birth, SGA, LGA, LBW, HBW, and neonatal intensive care unit admission) were derived from maternity records; for women with more than one pregnancy record, a random record was selected. To note, not every participant have a HES/maternity record, as not all have been admitted to hospital within the period covered.

#### Genotyping and imputing

A full description of the genetic QC performed by the MRC-IEU on UK Biobank data has already been published.^25^ The full data release contains the cohort of successfully genotyped samples (n=488,377). Most participants (n=438,398) were genotyped using the UK Biobank axiom array, and 49,979 individuals were genotyped using the UK BiLEVE array. Pre-imputation QC, phasing and imputation are described elsewhere.^26^ In brief, prior to phasing, multiallelic SNPs or those with MAF ≤1% were removed. Phasing of genotype data was performed using a modified version of the SHAPEIT2 algorithm.^27^ Genotype imputation to a reference set combining the UK10K haplotype and HRC reference panels was performed using IMPUTE2 algorithms. The analyses presented here were restricted to autosomal variants using a graded filtering with varying imputation quality for different allele frequency ranges. Therefore, rarer genetic variants are required to have a higher imputation INFO score (Info>0.3 for MAF >3%; Info>0.6 for MAF 1-3%; Info>0.8 for MAF 0.5-1%; Info>0.9 for MAF 0.1-0.5%) with MAF and Info scores having been recalculated on an in-house derived ‘European’ subset.

In post-imputation quality control, individuals with sex-mismatch (derived by comparing genetic sex and reported sex) or individuals with sex chromosome aneuploidy were excluded from the analysis (n=814). The sample was restricted to individuals of ‘European’ ancestry as defined by an in-house k-means cluster analysis performed using the first 4 principal components provided by UK Biobank in the statistical software environment R. The current analysis includes the largest cluster from this analysis (n=464,708). Estimated kinship coefficients using the KING toolset identified 107,162 pairs of related individuals.^26^ An in-house algorithm was then applied to this list and preferentially removed the individuals related to the greatest number of other individuals until no related pairs remain. These individuals were excluded (n=79,448). Additionally two individuals were removed due to them relating to a very large number (>200) of individuals.

### ACKNOWLEDGEMENTS

*ALSPAC:* We are extremely grateful to all the families who took part in this study, the midwives for their help in recruiting them, and the whole ALSPAC team, which includes interviewers, computer and laboratory technicians, clerical workers, research scientists, volunteers, managers, receptionists and nurses. Please note that the study website contains details of all the data that is available through a fully searchable data dictionary and variable search tool" and reference the following webpage: http://www.bristol.ac.uk/alspac/researchers/our-data/

*BiB*: Born in Bradford is only possible because of the enthusiasm and commitment of the children and parents in BiB. We are grateful to all the participants, health professionals and researchers who have made Born in Bradford happen.

*DNBC-PTB controls:* We are very grateful to all DNBC families who took part in the study. We would also like to thank everyone involved in data collection and biological material handling.

*EFSOCH:* We are grateful to all families who participated in the Exeter Family Study of Childhood Health, and to the EFSOCH study team.

*FinnGen*: We are grateful to participants and investigators from the FinnGen study.

*Gen-3G:* We thank all families who are participating to the Gen3G cohort. The Gen3G investigators acknowledge the blood sampling in pregnancy clinic at the CHUS including the assistance of clinical research nurses for recruiting women and obtaining consent for the study in addition to the CHUS biomedical laboratory for performing some clinical assays.

*Generation R:* The Generation R Study is conducted by Erasmus MC in close collaboration with the School of Law and Faculty of Social Sciences of the Erasmus University Rotterdam, the Municipal Health Service Rotterdam area, Rotterdam, the Rotterdam Homecare Foundation, Rotterdam, and the Stichting Trombosedienst & Artsenlaboratorium Rijnmond (STAR-MDC), Rotterdam. We gratefully acknowledge the contribution of children and parents, general practitioners, hospitals, midwives, and pharmacies in Rotterdam.

*HAPO:* The authors are indebted to the participants of the HAPO Study at the following centers: Newcastle and Brisbane, Australia; Toronto, Ontario, Canada; and Belfast, U.K.

*INMA:* The authors would like to thank all the participants for their generous collaboration. A full roster of the INMA Project Investigators can be found at <http://www.proyectoinma.org/presentacion-inma/listado-investigadores/en_listado-investigadores.html>. In addition, authors would like to thank the Basque Biobank and the Spanish Genotyping Center (CEGEN).

*MoBa*: The Norwegian Mother, Father and Child Cohort Study is supported by the Norwegian Ministry of Health and Care Services and the Ministry of Education and Research. We are grateful to all the participating families in Norway who take part in this on-going cohort study. We thank the Norwegian Institute of Public Health (NIPH) for generating high-quality genomic data. This research is part of the HARVEST collaboration, supported by the Research Council of Norway (#229624). We also thank the NORMENT Centre for providing genotype data, funded by the Research Council of Norway (#223273), South East Norway Health Authorities and Stiftelsen Kristian Gerhard Jebsen. We further thank the Center for Diabetes Research, the University of Bergen for providing genotype data and performing quality control and imputation of the data funded by the ERC AdG project SELECTionPREDISPOSED, Stiftelsen Kristian Gerhard Jebsen, Trond Mohn Foundation, the Research Council of Norway, the Novo Nordisk Foundation, the University of Bergen, and the Western Norway Health Authorities”. This research has been conducted using MoBa data using application number 2552.

*NFBC1966:* We thank all cohort members and researchers who participated in the 31 and 46 yrs study. We also wish to acknowledge the work of the NFBC project center.

*NFBC1986:* We thank all cohort members and researchers who have participated in the study. We also wish to acknowledge the work of the NFBC project center.

*UK Biobank:* The authors are grateful to UK Biobank participants and investigators for access to data to undertake this study (Project #23938).

### FUNDING

*ALSPAC study funding:* The UK Medical Research Council and Wellcome (Grant ref: 217065/Z/19/Z) and the University of Bristol provide core support for ALSPAC. This publication is the work of the authors and MCB, GC and DAL will serve as guarantors for the contents of this paper. A comprehensive list of grants funding is available on the ALSPAC website (http://www.bristol.ac.uk/alspac/external/documents/grant-acknowledgements.pdf); GWAS data was generated by Sample Logistics and Genotyping Facilities at Wellcome Sanger Institute and LabCorp (Laboratory Corporation of America) using support from 23andMe.

*BiB study funding:* Born in Bradford is supported by a Wellcome programme grant (WT223601/Z/21/Z: Age of Wonder), a Wellcome  infrastructure grant (WT101597MA), a joint Medical Research Council (MRC) and UK Economic and Social Science Research Council (ESRC) programme grant (MR/N024391/1) and a British Heart Foundation Clinical Study Grant (CS/16/4/32482). Funding for DNA extraction and genotyping was from two MRC programme grants (MC_UU_00011/6 and MC_UU_12013/5).

*DNBC-GOYA study funding:* GOYA is nested within the Danish National Birth Cohort which was established with a significant grant from the Danish National Research Foundation. Additional support was obtained from the Danish Regional Committees, the Pharmacy Foundation, the Egmont Foundation, the March of Dimes Birth Defects Foundation, the Health Foundation and other minor grants. The DNBC Biobank has been supported by the Novo Nordisk Foundation and the Lundbeck Foundation. The genotyping for GOYA was funded by the Wellcome Trust (WT 084762).

*DNBC-PTB (controls) study funding:* The Danish National Birth Cohort (DNBC) is a result of major grants from the Danish National Research Foundation, the Danish Pharmacists’ Fund, the Egmont Foundation, the March of Dimes Birth Defects Foundation, the Augustinus Foundation, and the Health Fund of the Danish Health Insurance Societies. The DNBC biobank is a part of the Danish National Biobank resource, which is supported by the Novo Nordisk Foundation. The generation of GWAS genotype data for the DNBC samples was carried out within the Gene Environment Association Studies (GENEVA) consortium with funding provided through the National Institutes of Health’s Genes, Environment, and Health Initiative (U01HG004423; U01HG004446; U01HG004438).

*EFSOCH study funding:* The Exeter Family Study of Childhood Health (EFSOCH) was supported by South West NHS Research and Development, Exeter NHS Research and Development, the Darlington Trust and the Peninsula National Institute of Health Research (NIHR) Clinical Research Facility at the University of Exeter. The opinions given in this paper do not necessarily represent those of NIHR, the NHS or the Department of Health. Genotyping of the EFSOCH study samples was funded by the Wellcome Trust and Royal Society Grant 104150/Z/14/Z.

*Gen3G study funding:* Gen3G was supported by a Fonds de recherche du Québec en santé (FRQ-S) operating grant (to MFH, grant #20697); a Canadian Institute of Health Reseach (CIHR) Operating grant (to MFH grant #MOP 115071); a Diabète Québec grant (to PP) and is currently support by CIHR PJT-152989 (LB).

*Generation R study funding:* The general design of the Generation R Study is made possible by financial support from Erasmus MC, University Medical Center Rotterdam, Erasmus University Rotterdam, the Netherlands Organization for Health Research and Development and the Ministry of Health, Welfare and Sport.  This project received funding from the European Union’s Horizon 2020 research and innovation programme (733206, LifeCycle, 874739, LongITools, 824989 EUCAN-Connect).

*HAPO study funding:* This study was supported by National Institutes of Health (NIH) grants HD-34242, HD-34243, HG-004415, and CA-141688, and by the American Diabetes Association.

*INMA study funding:* This study was funded by grants from Instituto de Salud Carlos III (CB06/02/0041, G03/176, FIS PS09/00432, FIS-FEDER 03/1615, 04/1509, 04/1112, 04/1931, 05/1079, 05/1052, 06/1213, 06/0867, 07/0314, and 09/02647), Fundació La Marató de TV3 (090430), Generalitat de Catalunya-CIRIT (1999SGR 00241), Conselleria de Sanitat Generalitat Valenciana, and Fundación Roger Torné, Department of Health of the Basque Government (2005111093), Provincial Government of Gipuzkoa (DFG06/002), and annual agreements with the municipalities of the study area (Zumarraga, Urretxu , Legazpi, Azkoitia y Azpeitia y Beasain). ISGlobal We acknowledge support from the Spanish Ministry of Science and Innovation through the “Centro de Excelencia Severo Ochoa 2019-2023” Program (CEX2018-000806-S), and from the Generalitat de Catalunya through the CERCA Program.

*MoBa study funding:* MoBa is supported by the Norwegian Ministry of Health and Care services and the Ministry of Education and Research.

*NFBC1966 study funding:* NFBC1966 received financial support from University of Oulu Grant no. 65354, Oulu University Hospital Grant no. 2/97, 8/97, Ministry of Health and Social Affairs Grant no. 23/251/97, 160/97, 190/97, National Institute for Health and Welfare, Helsinki Grant no. 54121, Regional Institute of Occupational Health, Oulu, Finland Grant no. 50621, 54231 NFBC1966 received financial support from University of Oulu Grant no. 24000692, Oulu University Hospital Grant no. 24301140, ERDF European Regional Development Fund Grant no. 539/2010 A31592.

*NFBC1986 study funding:* NFBC1986 received financial support from EU QLG1-CT-2000-01643 (EUROBLCS) Grant no. E51560, NorFA Grant no. 731, 20056, 30167, USA / NIH 2000 G DF682 Grant no. 50945. This work was supported by the European Union’s Horizon 2020 research and innovation program under grant agreement No. 633595 (DynaHEALTH), grantagreement no. 733206 (LifeCycle), grant agreement no. 873749 (LongITools), grant agreement no. 848158 (EarlyCause).
