## Supplementary figures for "Integrating multiple lines of evidence to assess the effects of maternal BMI on pregnancy and perinatal outcomes in up to 497,932 women"

### Supplementary contents page

**Supplementary figure 1** - Multivariable regression and Mendelian randomization pooled association estimates for the relation between maternal BMI and continuous outcomes across studies using a fixed effect meta-analysis

**Supplementary figure 2** - Scatter plot of the pooled SNP-BMI association estimates from females in the GIANT GWAS and pregnant women from participating studies

**Supplementary figure 3** - Multivariable regression and paternal negative control pooled association estimates for the relation between maternal/paternal BMI and continuous outcomes across studies using a fixed effect meta-analysis

**Supplementary figure 4** - Multivariable regression study specific and pooled associations using a fixed effect meta-analysis for (A and B) binary outcomes and (C) continuous outcomes

**Supplementary figure 5** - Comparison of Multivariable regression pooled estimates across studies using fixed- or random-effect metanalyses for (A) binary outcomes and (B) continuous outcomes

**Supplementary figure 6** - Comparison of Multivariable regression pooled estimates across different models for (A) binary and (B) continuous outcomes

**Supplementary figure 7** - Leave-one-out analyses removing one **study** at a time and re-estimating Mendelian randomization results using IVW for binary (A and B) and continuous (C) outcomes.

**Supplementary figure 8** - Comparison of Mendelian randomization IVW results using summary data for the SNP-outcomes association pooled across studies using fixed- or random-effect metanalyses for (A) binary and (B) continuous outcomes.

**Supplementary figure 9** - Leave-one-out analyses removing one SNP at a time and re-estimating Mendelian randomization results using IVW

**Supplementary figure 10** - Comparison of Mendelian randomization results using different methods for (A) binary and (B) continuous outcomes

**Supplementary figure 11** - Comparison of Mendelian randomization IVW results using summary data for the SNP-outcomes association unadjusted or adjusted for offspring genotype for (A) binary and (B) continuous outcomes

**Supplementary figure 12** - Partner negative control - study specific and pooled – maternal - associations using a fixed effect meta-analysis for (A) binary outcomes and (B) continuous outcomes

**Supplementary figure 13** - Partner negative control - study specific and pooled – paternal - associations using a fixed effect meta-analysis for (A) binary outcomes and (B) continuous outcomes

**Supplementary figure 14** - Partner negative control – sensitivity analyses – for (A) binary outcomes and (B) continuous outcomes: ALSPAC

**Supplementary figure 15** - Partner negative control – sensitivity analyses – for (A) binary outcomes and (B) continuous outcomes: MoBa

**Supplementary figure 16** - Partner negative control – sensitivity analyses – for (A) binary outcomes and (B) continuous outcomes: GenR

**Supplementary figure 17** - Comparison of multivariable regression pooled estimates across studies using fixed- or random-effect metanalyses split by studies reporting pre and during pregnancy weight for (A) binary outcomes and (B) continuous outcomes.

### **Supplementary figure 1 -** Multivariable regression and Mendelian randomization pooled association estimates for the relation between maternal BMI and continuous outcomes across studies using a fixed effect meta-analysis

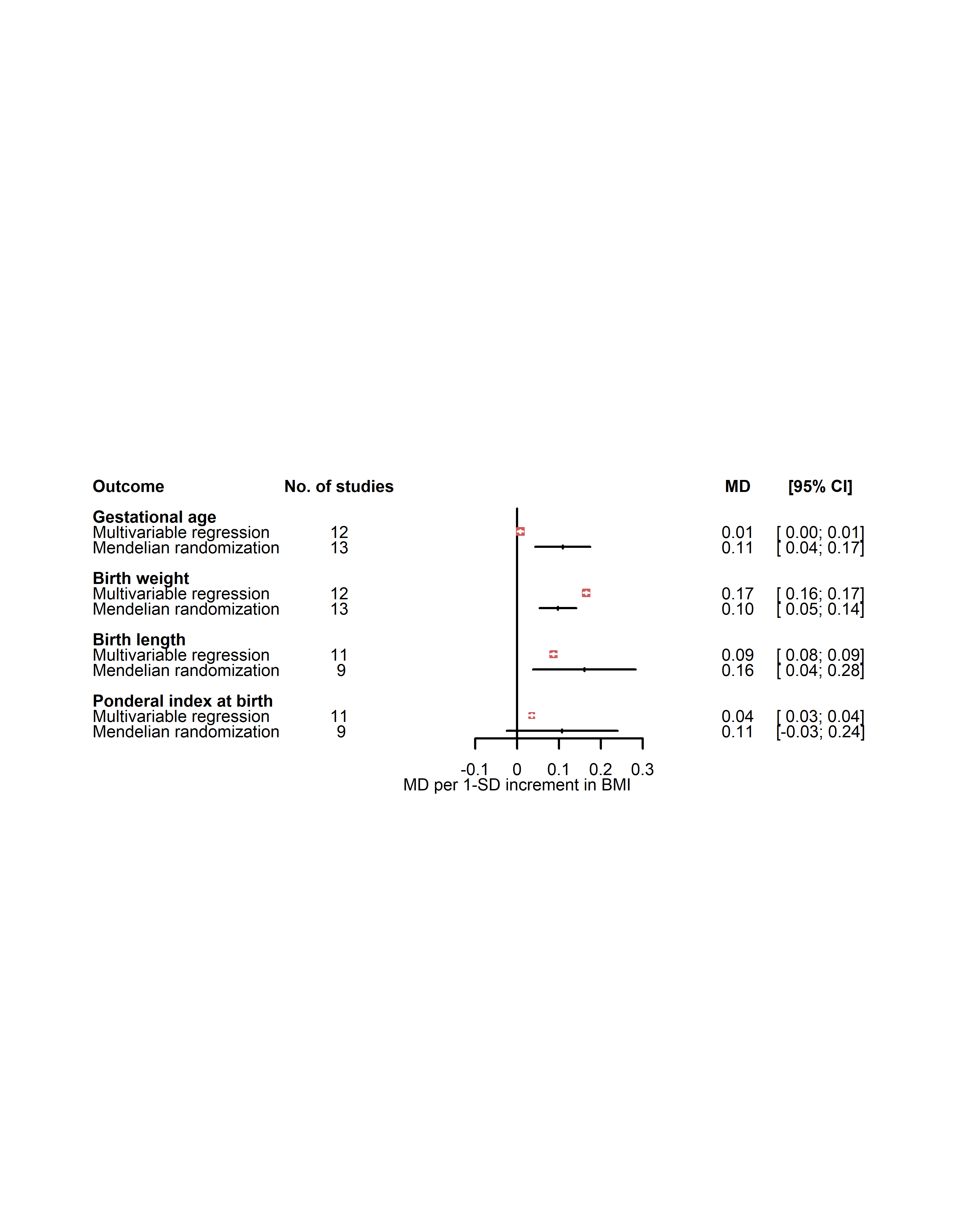

Results are expressed as mean difference in SD units of continuous outcomes per SD unit of maternal BMI. Multivariable regression results were adjusted for maternal age, parity, education, smoking during pregnancy, alcohol use during pregnancy and offspring sex where available. Study-specific estimates were pooled using a fixed-effect meta analysis. MD: mean difference; SD: standard deviation units.

### Supplementary figure 2 - Scatter plot of the pooled SNP-BMI association estimates from females in the GIANT GWAS and pregnant women from participating studies

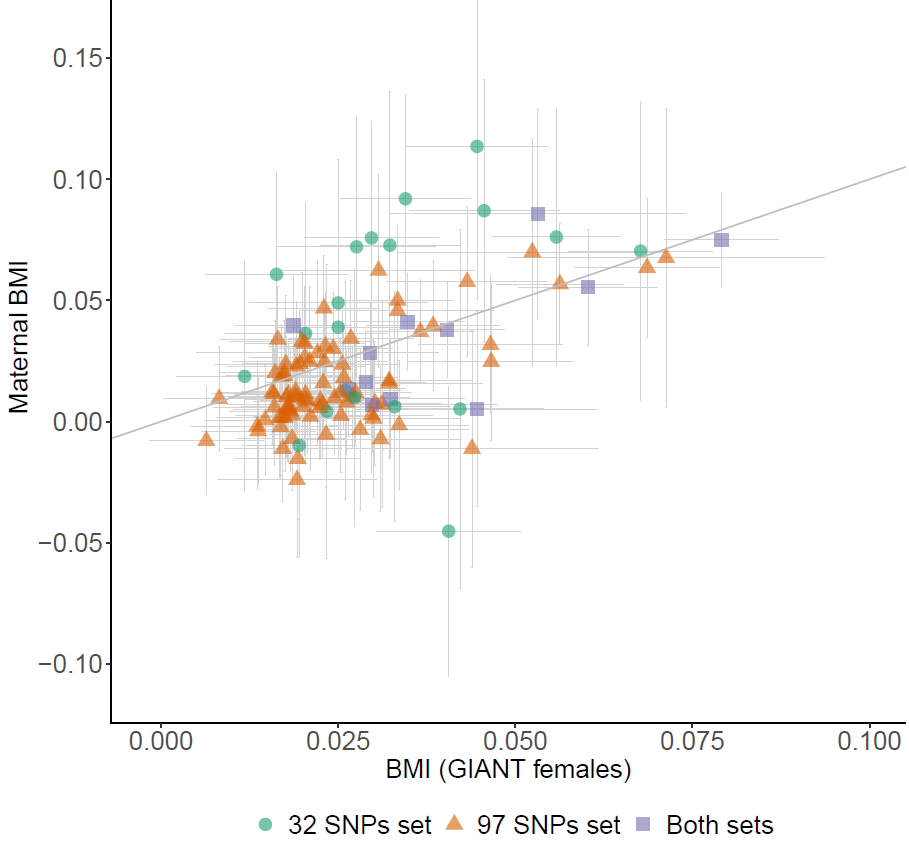

Results are expressed as change in mean BMI in SD units per additional effect allele for selected SNPs. Symbols reflect point estimates and bars reflect corresponding 95% confidence internal. BMI: body mass index; SNP: single nucleotide polymorphism; GWAS: genome-wide association study; GIANT: Genetic Investigation of ANthropometric Traits consortium.

### Supplementary figure 3 - Multivariable regression and paternal negative control pooled association estimates for the relation between maternal/paternal BMI and continuous outcomes across studies using a fixed effect meta-analysis

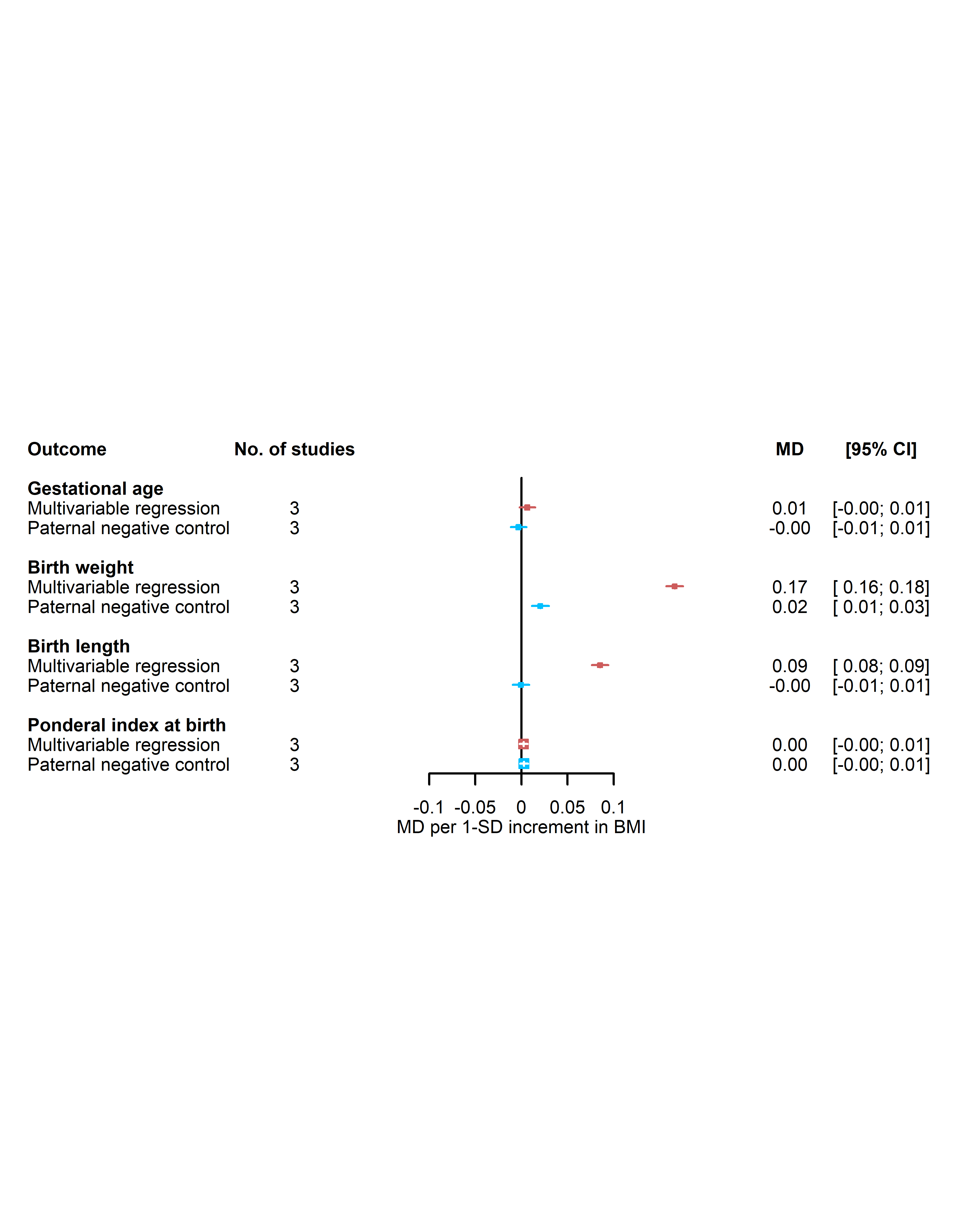

Results are expressed as mean difference in SD units of continuous outcomes per SD unit of maternal BMI and paternal BMI for ‘Multivariable regression’ and ‘Paternal negative control’, respectively. Multivariable regression results were adjusted for paternal BMI, maternal age, parity, education, smoking during pregnancy, alcohol use during pregnancy and offspring sex where available. Paternal negative control results were adjusted for maternal BMI, paternal age, number of children (ALSPAC only), paternal education, paternal smoking, paternal alcohol use and offspring sex where available. Study-specific estimates (ALSPAC, Gen-R, MoBa) were pooled using fixed-effect metanalyses. MD: mean difference; SD: standard deviation units; BMI: body mass index.

### Supplementary figure 4 - Multivariable regression study specific for (A and B) binary outcomes and (C) continuous outcomes

## A

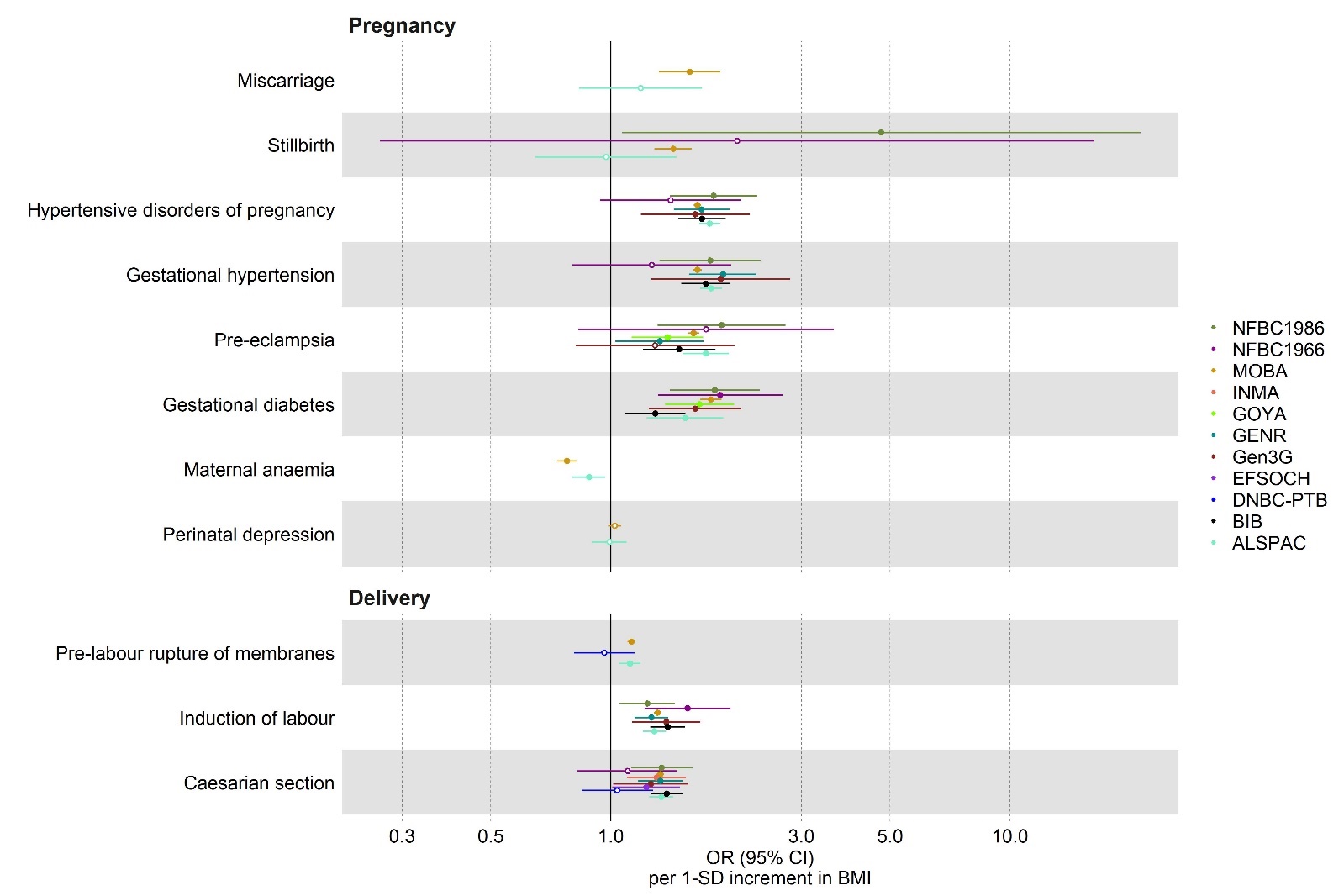

Results are expressed as OR of binary outcomes per SD unit of maternal BMI. Multivariable regression results were adjusted for maternal age, parity, education, smoking during pregnancy, alcohol use during pregnancy and offspring sex where available. OR: odds ratio; SD: standard deviation units; BMI: body mass index.

## B

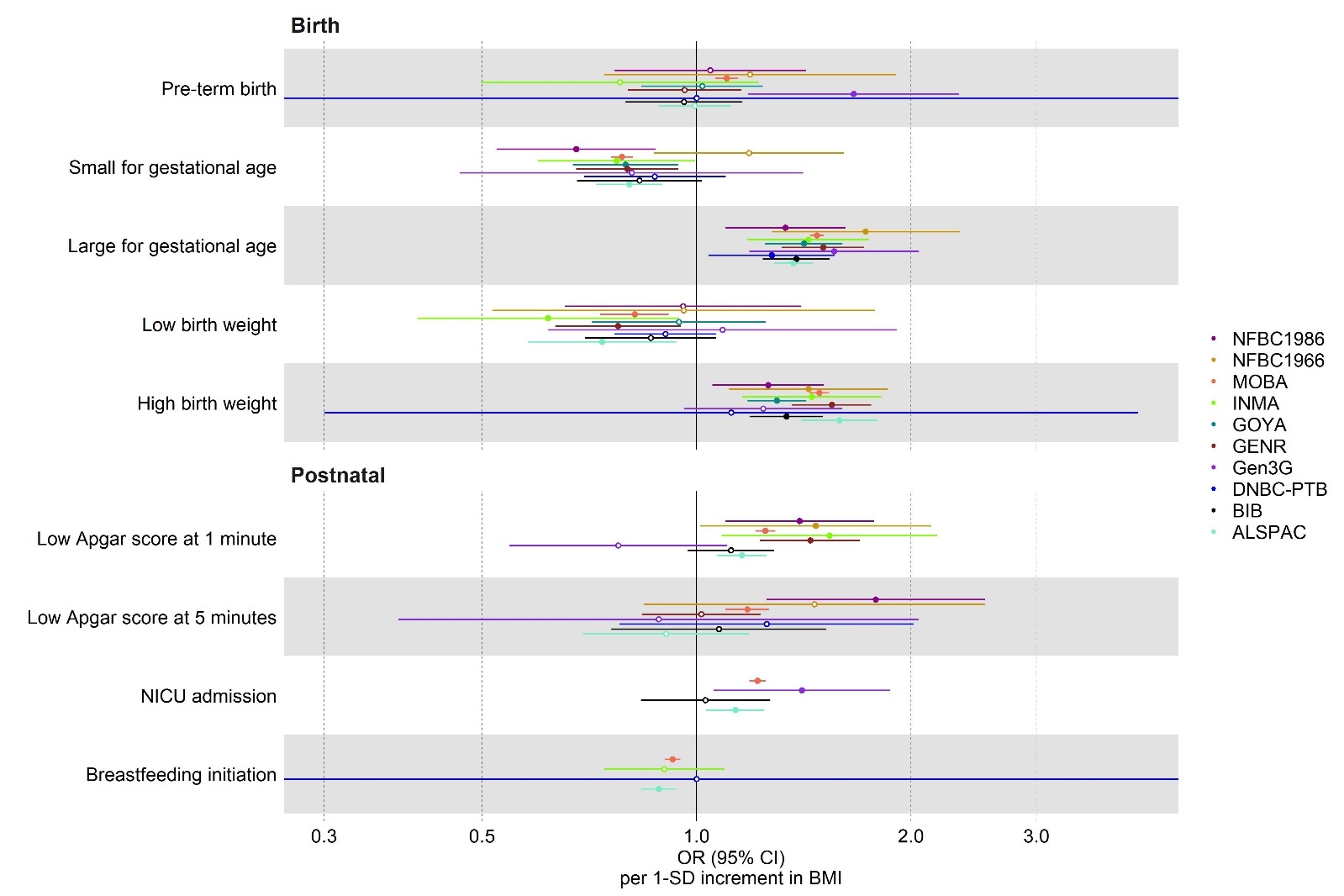

Results are expressed as OR of binary outcomes per SD unit of maternal BMI. Multivariable regression results were adjusted for maternal age, parity, education, smoking during pregnancy, alcohol use during pregnancy and offspring sex where available. OR: odds ratio; SD: standard deviation units; BMI: body mass index; NICU: neonatal intensive care unit.

## C

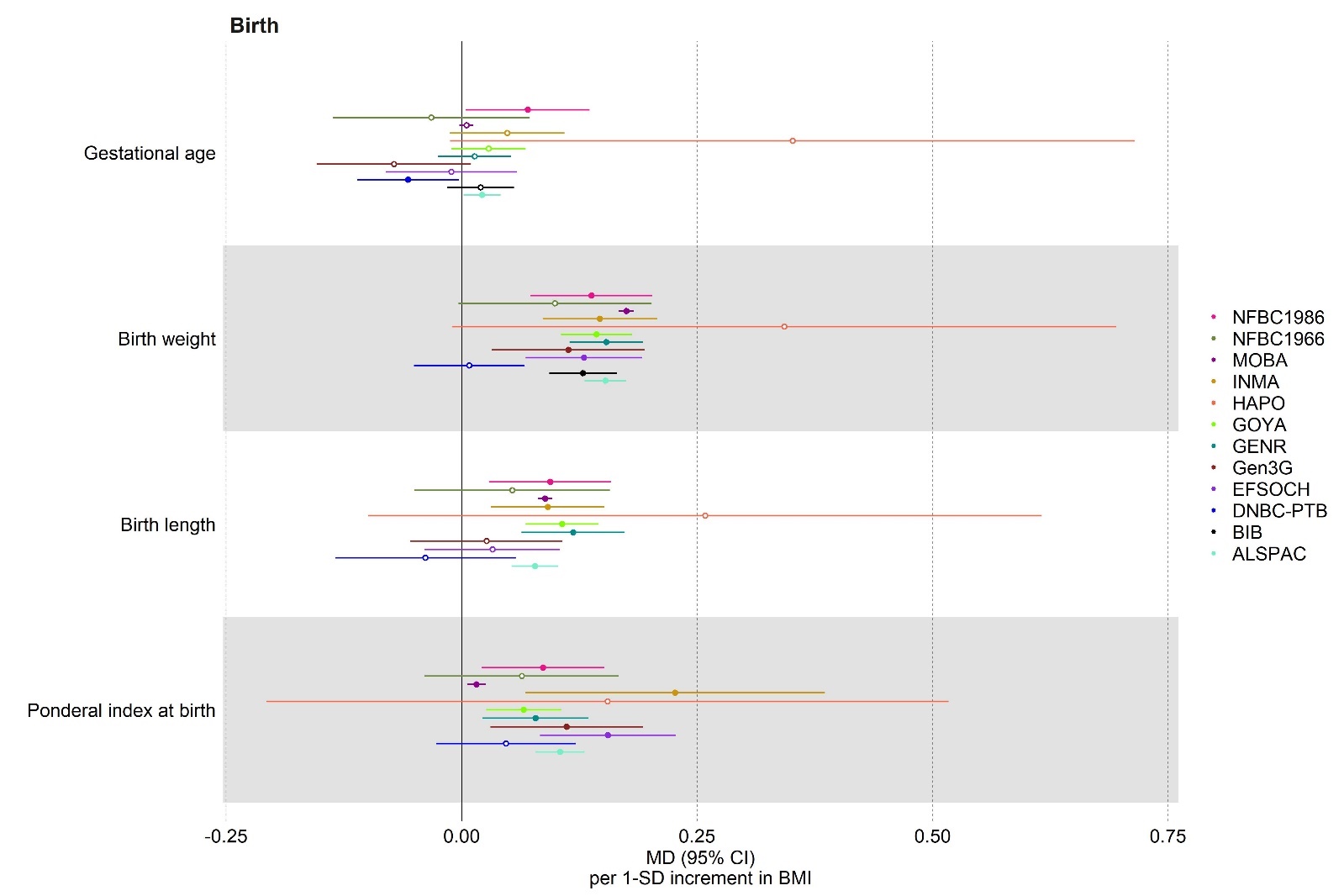

Results are expressed as mean difference in SD units of continuous outcomes per SD unit of maternal BMI. Multivariable regression results were adjusted for maternal age, parity, education, smoking during pregnancy, alcohol use during pregnancy and offspring sex where available. MD: mean difference; SD: standard deviation units; BMI: body mass index.

### Supplementary figure 5 - Comparison of Multivariable regression pooled estimates across studies using fixed- or random-effect metanalyses for (A) binary outcomes and (B) continuous outcomes.

## A

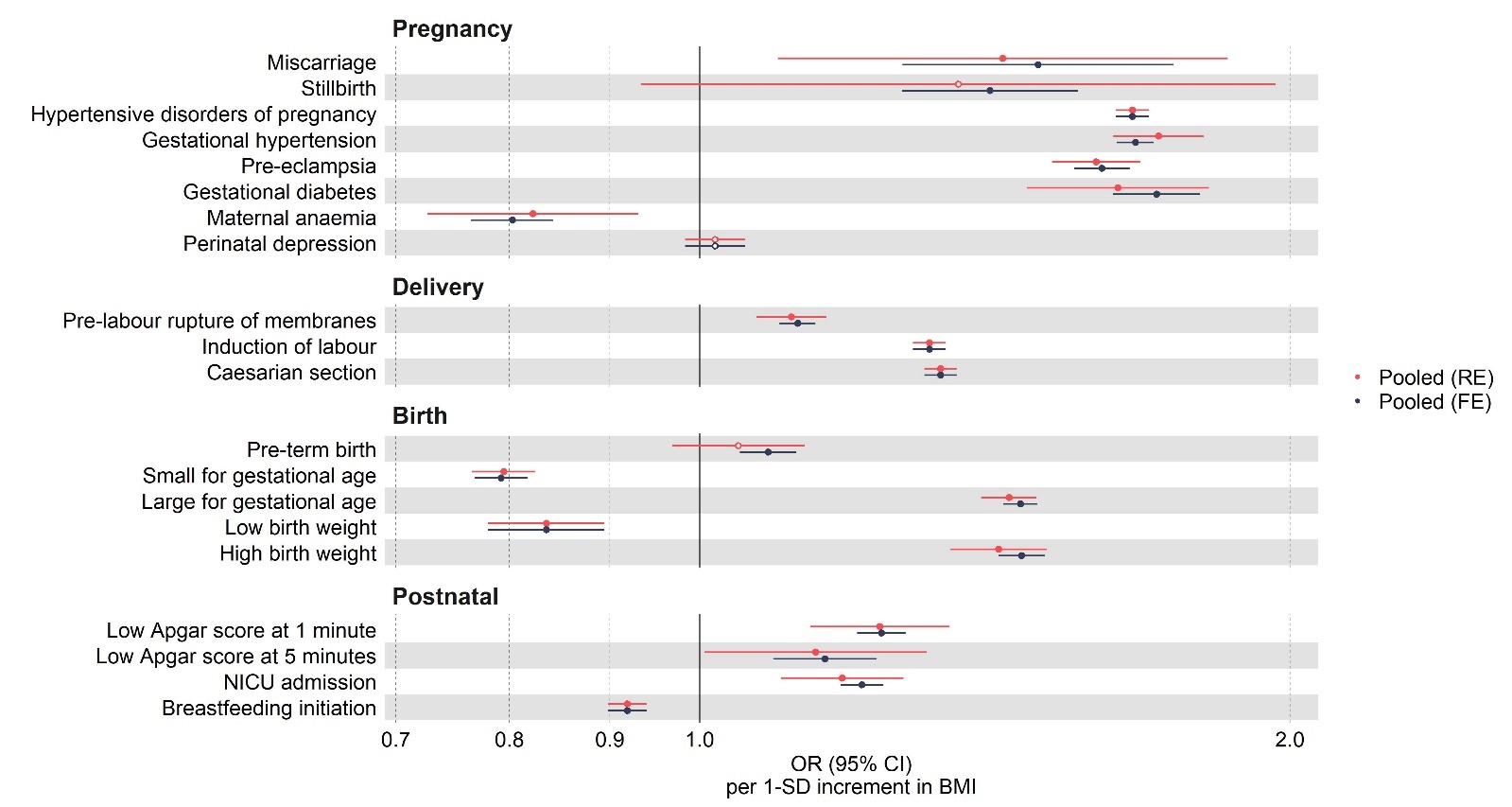

## B

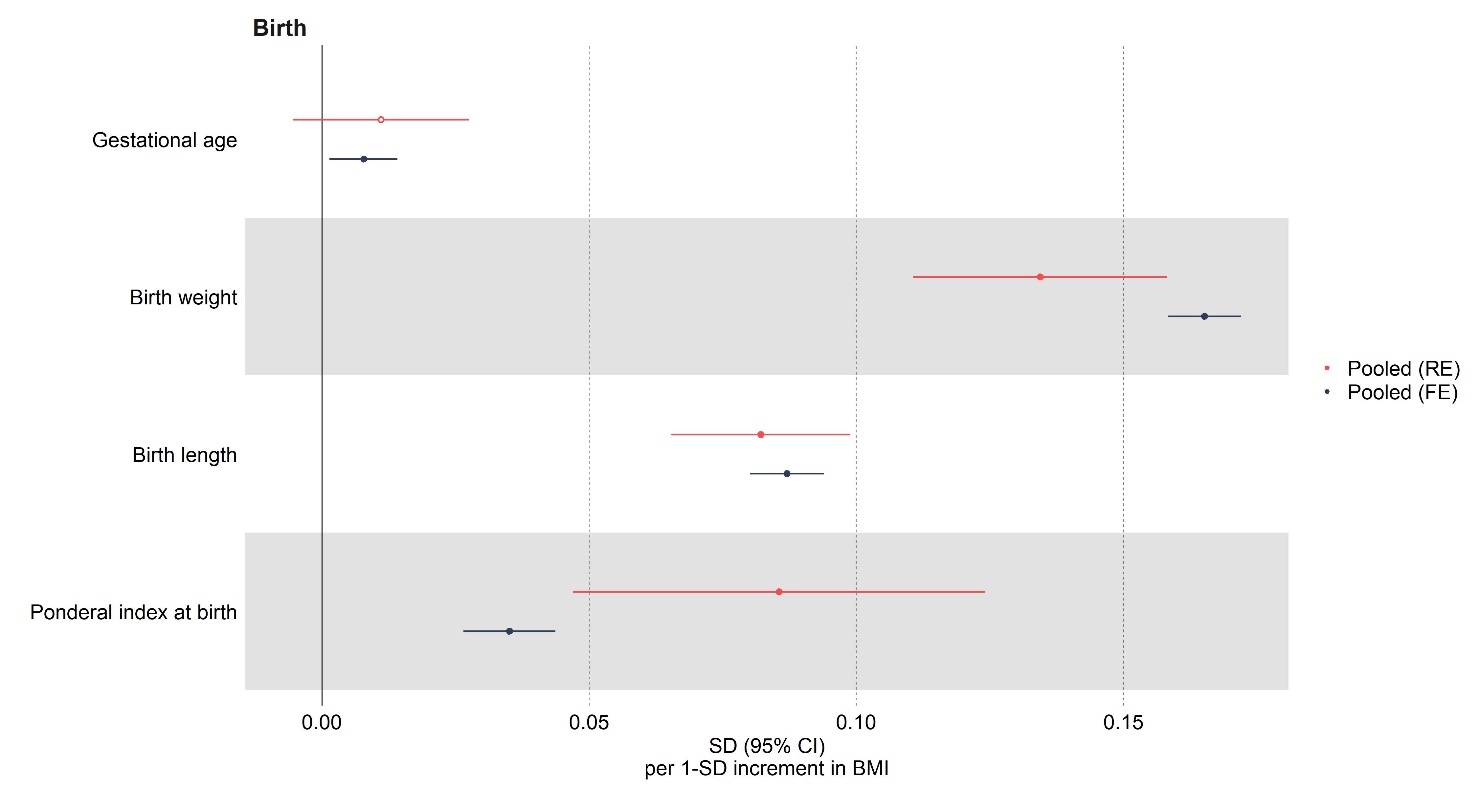

Results are expressed as OR of binary outcomes (A) or mean difference in SD units of continuous outcomes (B) per SD unit of maternal BMI. In each study, multivariable regression models were adjusted for maternal age, parity, education, smoking during pregnancy, alcohol use during pregnancy and offspring sex where available. Study-specific estimates were pooled using fixed- or random-effect metanalyses. SD: standard deviation units; BMI: body mass index; NICU: neonatal intensive care unit; FE: fixed-effect metanalysis; RE: random-effect metanalysis.

### Supplementary figure 6 - Comparison of Multivariable regression pooled estimates across different models for (A) binary and (B) continuous outcomes

## A

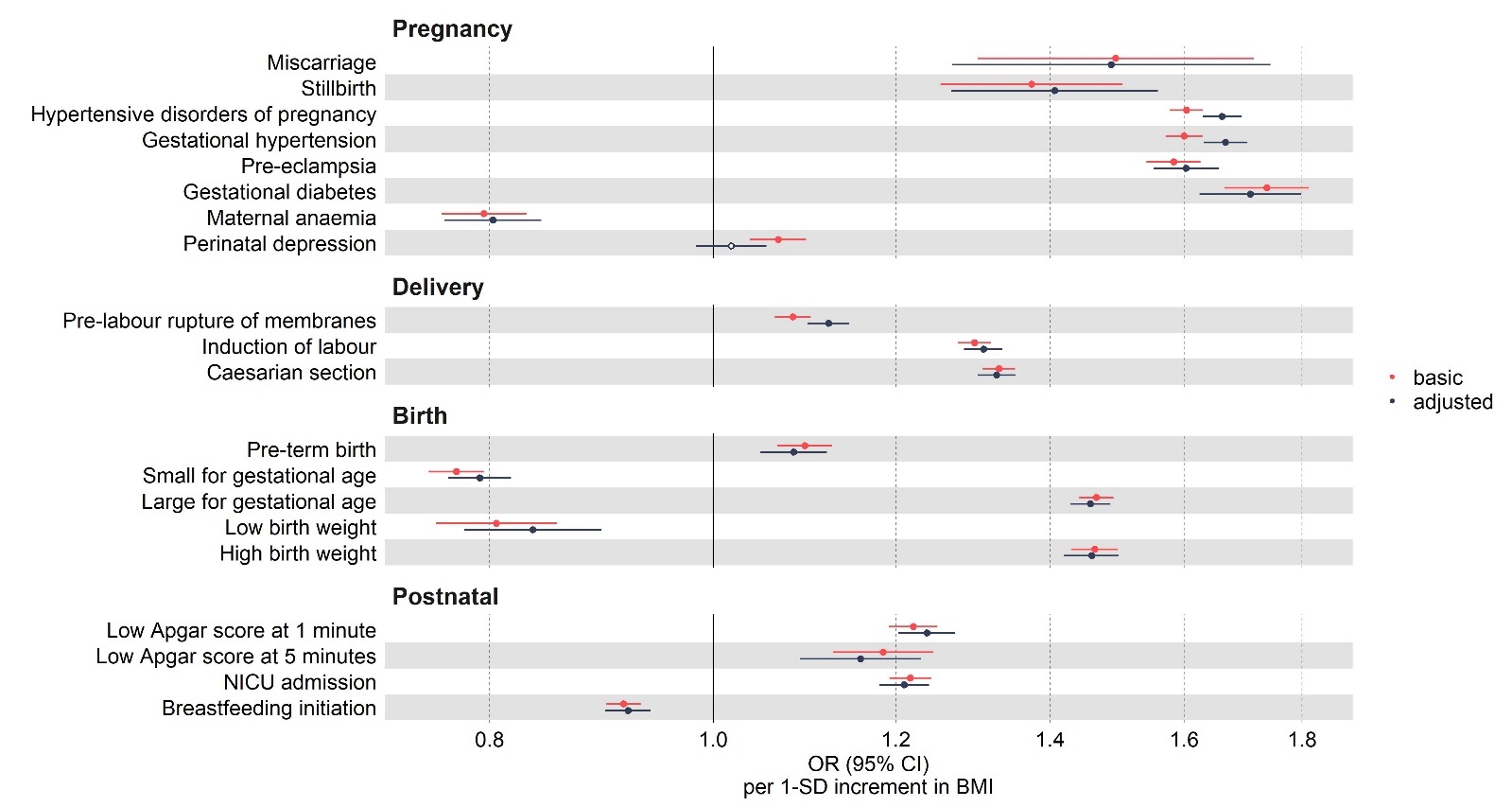

## B

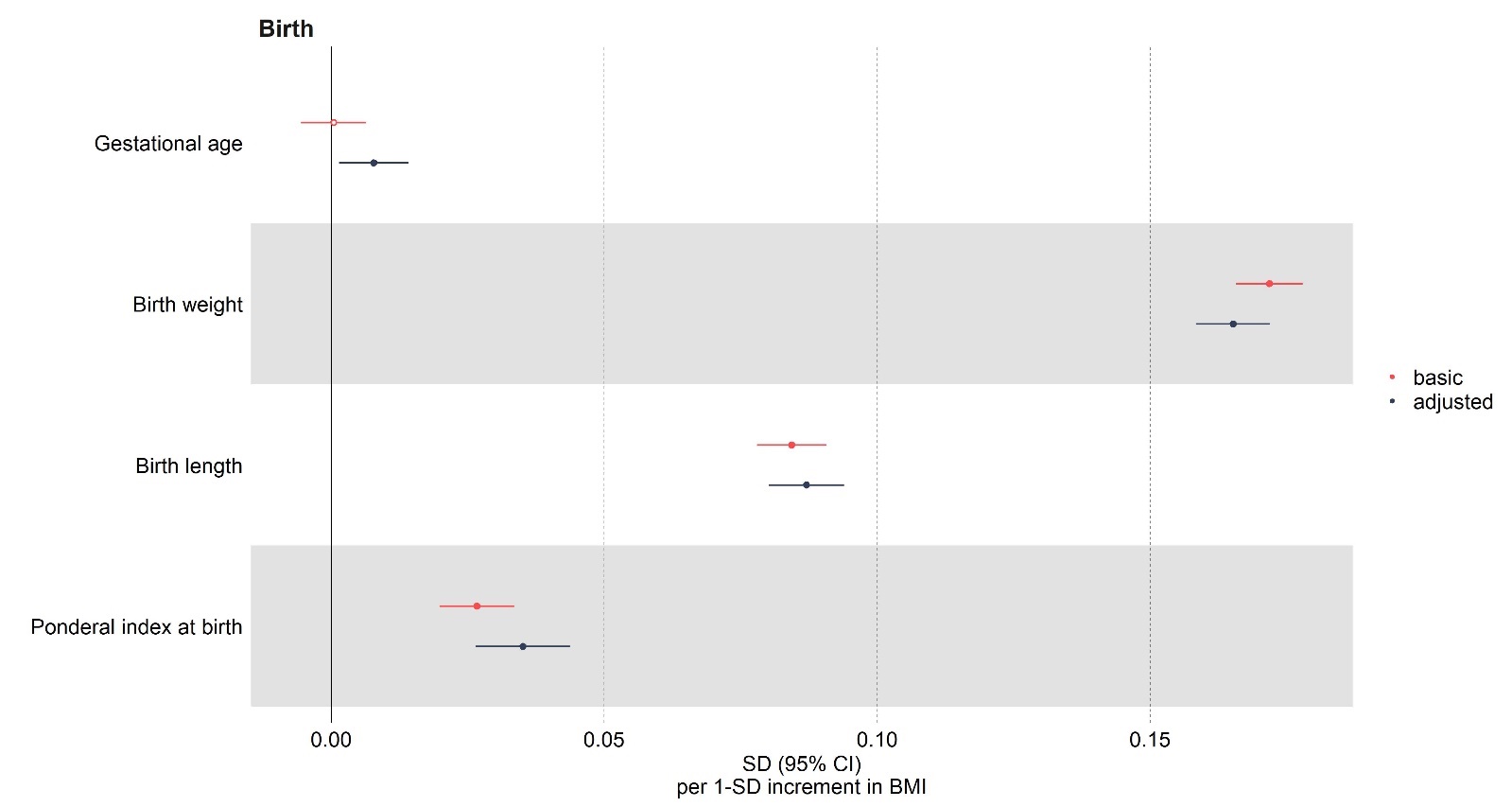

Results are expressed as OR of binary outcomes (A) or mean difference in SD units of continuous outcomes (B) per SD unit of maternal BMI. In each study, multivariable regression models were adjusted for maternal age and offspring sex (i.e. ‘basic’ model) or for maternal age, parity, education, smoking during pregnancy, alcohol use during pregnancy and offspring sex where available (‘adjusted’ model). Study-specific estimates were pooled using fixed -effect metanalyses. SD: standard deviation units; BMI: body mass index; NICU: neonatal intensive care unit; FE: fixed-effect metanalysis; RE: random-effect metanalysis.

### Supplementary figure 7 - Leave-one-out analyses removing one study at a time and re-estimating Mendelian randomization results using IVW for binary (A and B) and continuous (C) outcomes.

## A

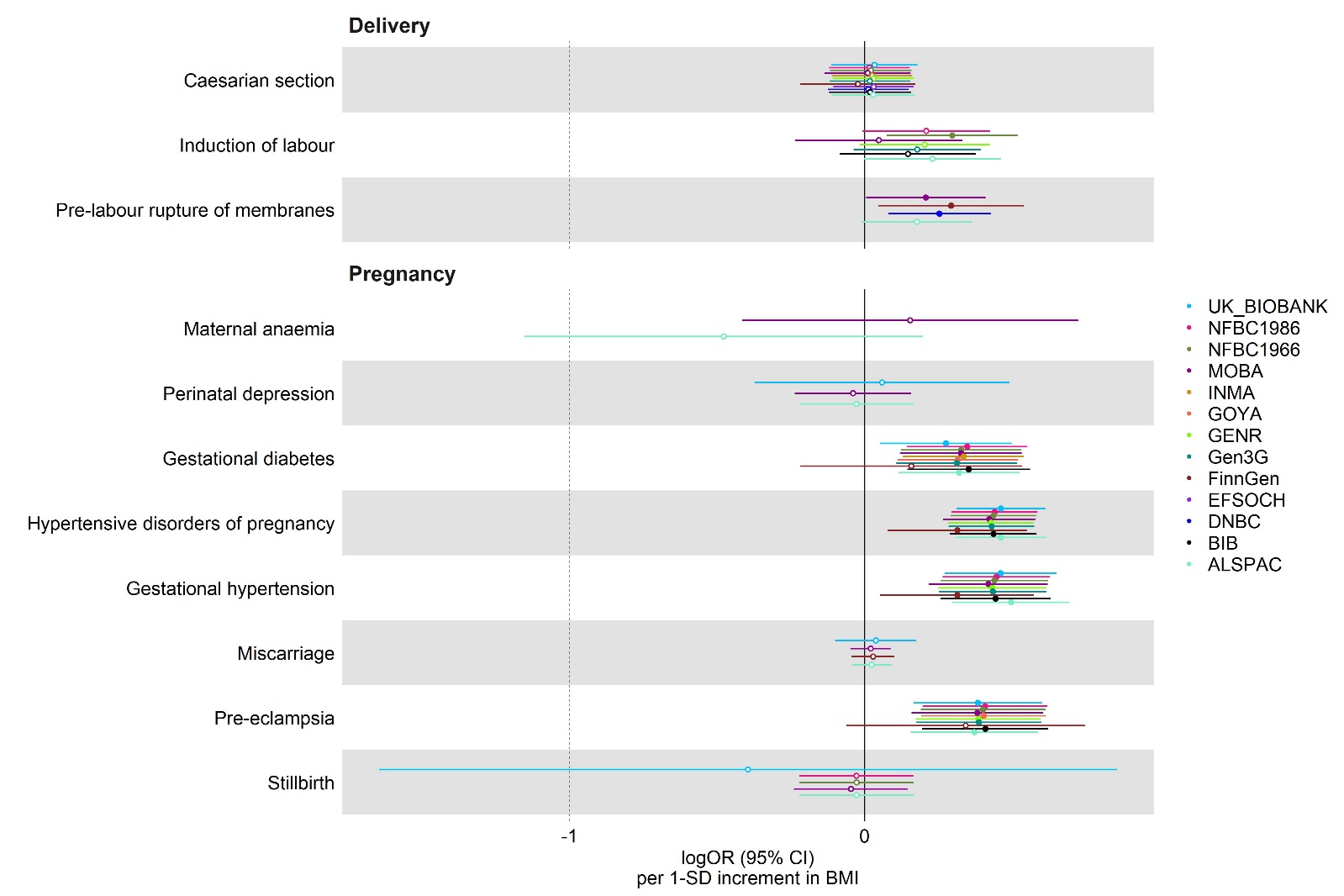

## B

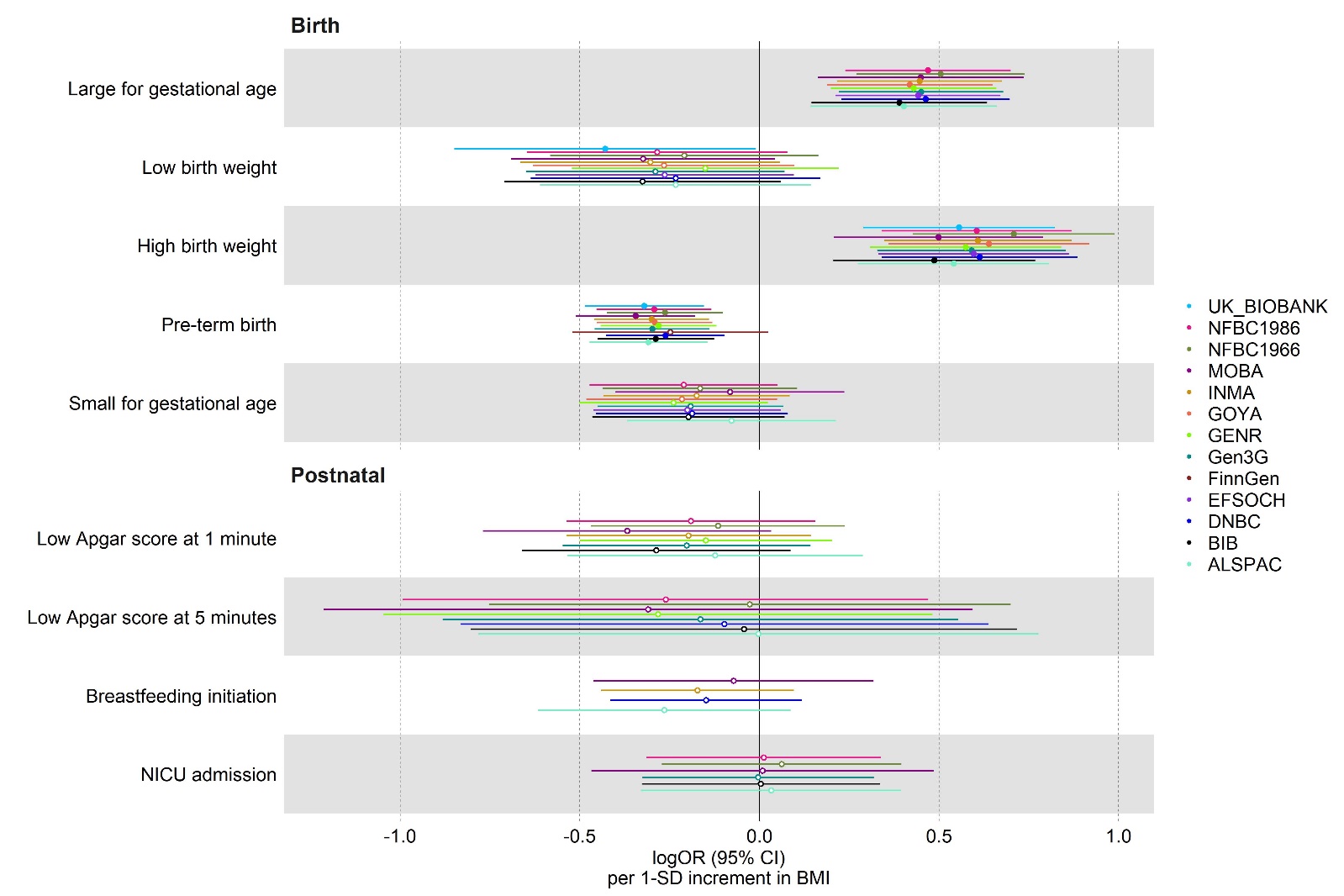

## C

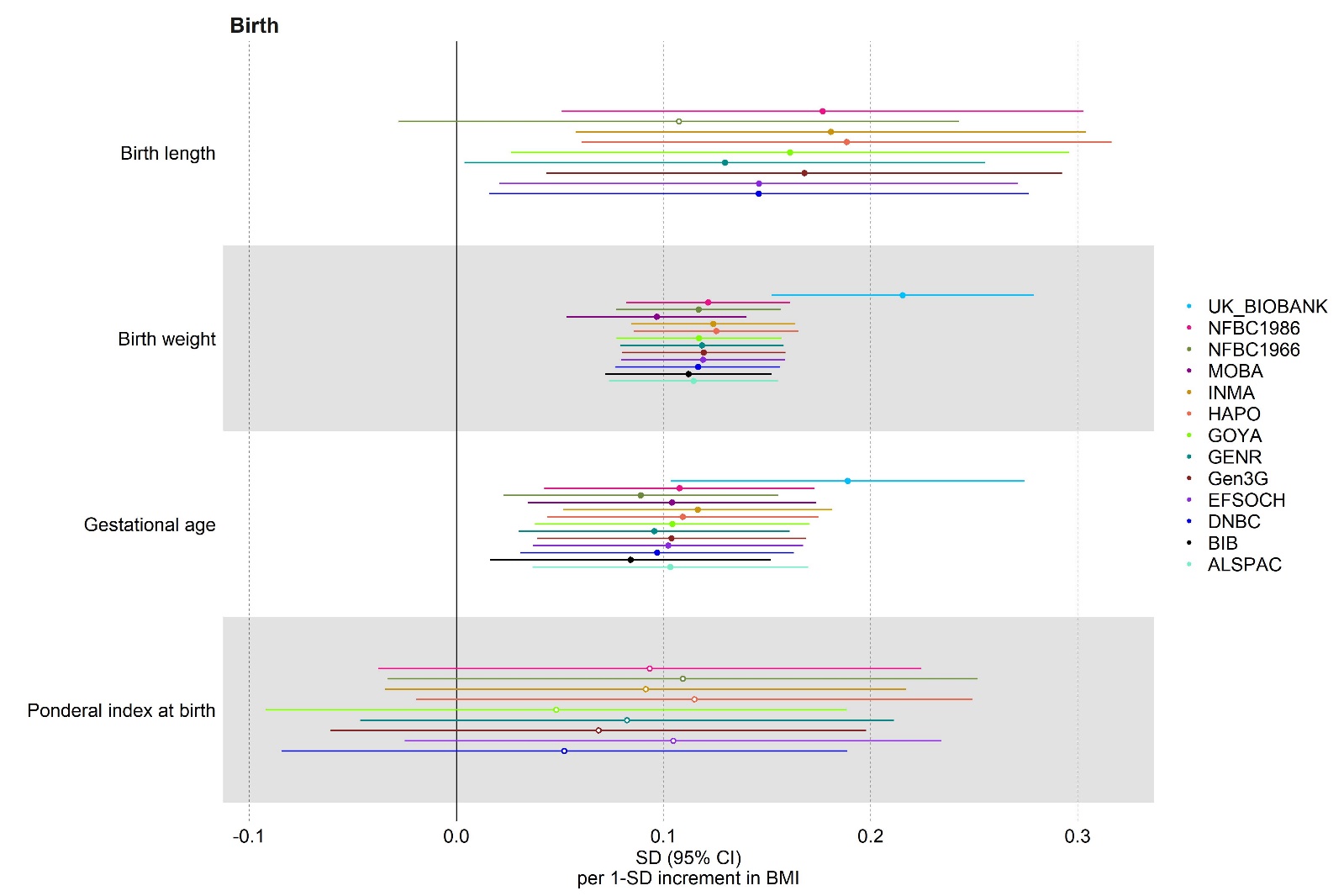

Results are expressed as log odds ratio (logOR) for binary outcomes (A and B) or mean difference in standard deviation (SD) units for continuous outcomes (C) per SD unit of maternal BMI. Study-specific results were estimated using IVW and pooled using fixed-effect metanalysis with one study removed at a time. IVW: inverse variance weighted method; BMI: body mass index; NICU: neonatal intensive care unit.

### Supplementary figure 8 - Comparison of Mendelian randomization IVW results using summary data for the SNP-outcomes association pooled across studies using fixed- or random-effect metanalyses for (A) binary and (B) continuous outcomes.

## A

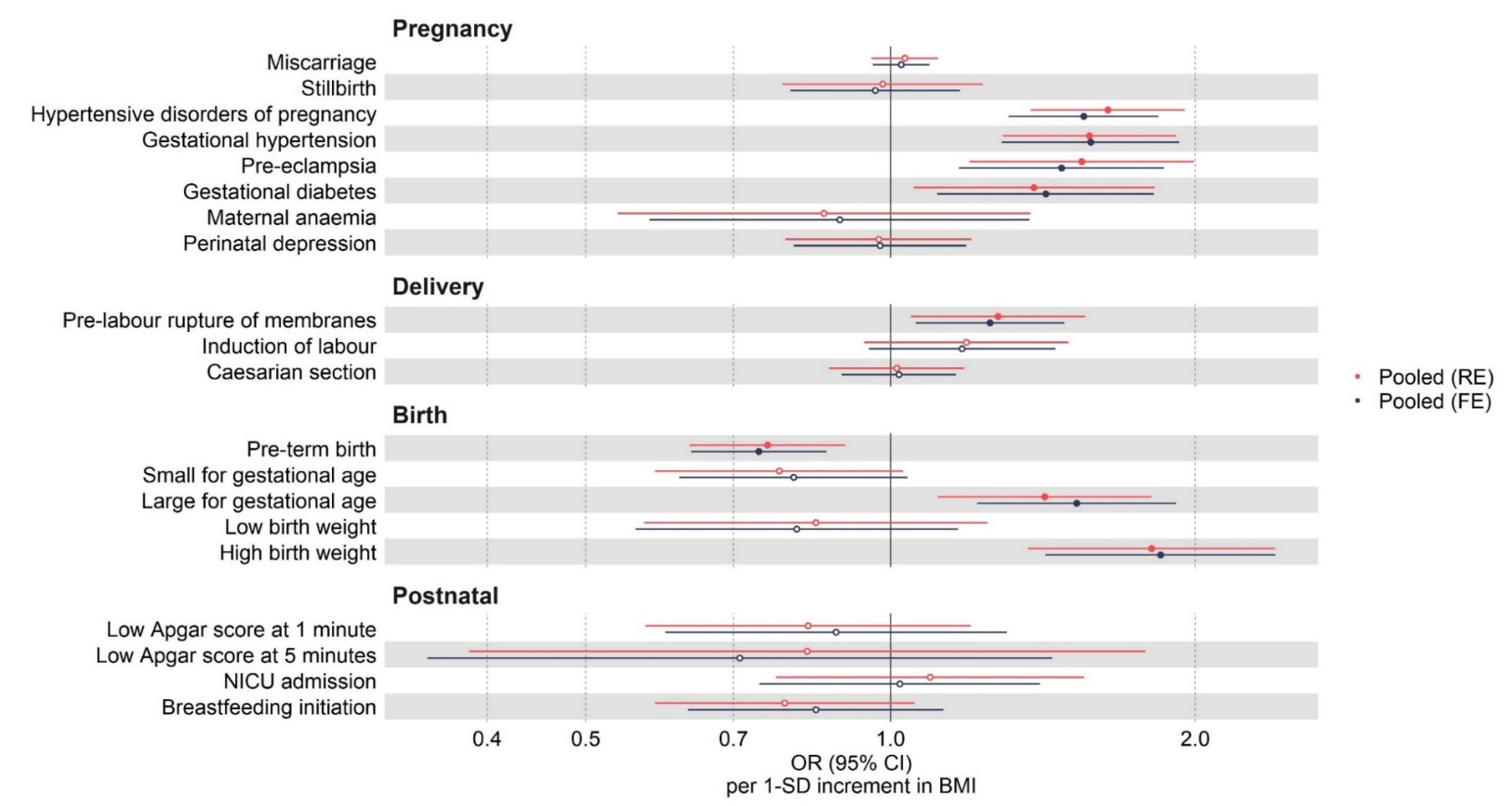
B

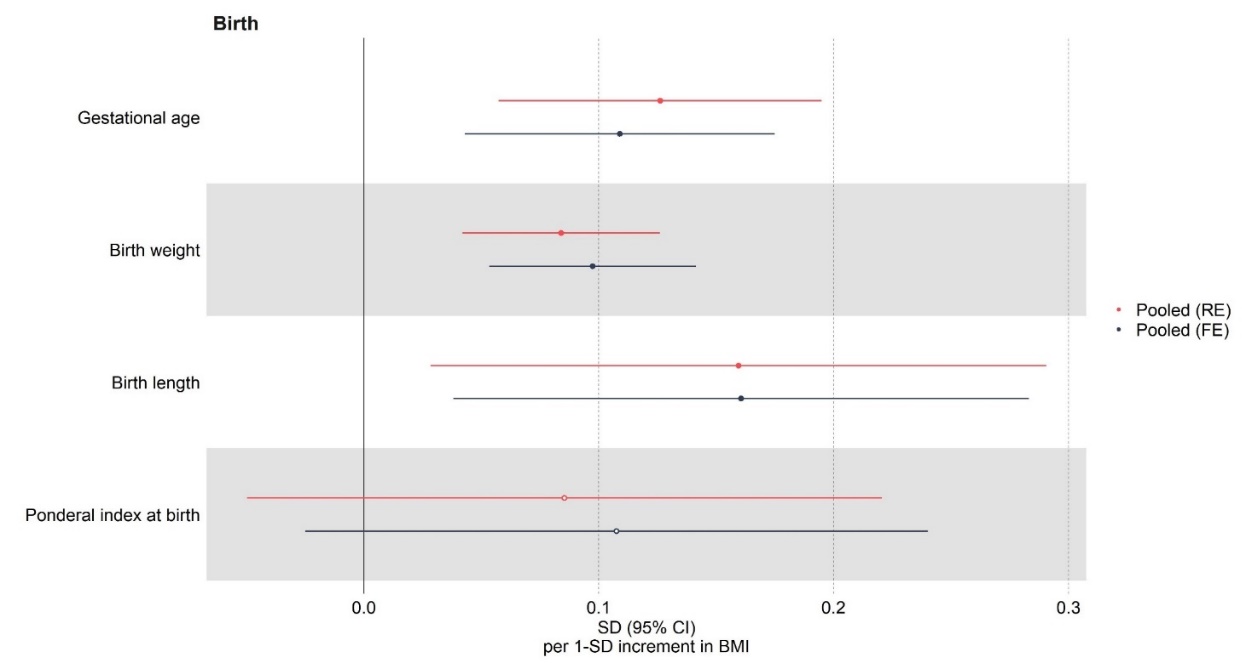

Results are expressed as odds ratio (OR) for binary outcomes (A) or mean difference in standard deviation (SD) units for continuous outcomes (B) per SD unit of maternal BMI. Study-specific results were estimated using IVW and pooled using fixed-effect metanalysis. IVW: inverse variance weighted method; BMI: body mass index; NICU: neonatal intensive care unit; FE: fixed-effect metanalyses; RE: random-effect metanalyses.

### Supplementary figure 9 - Leave-one-out analyses removing one SNP at a time and re-estimating Mendelian randomization results using IVW

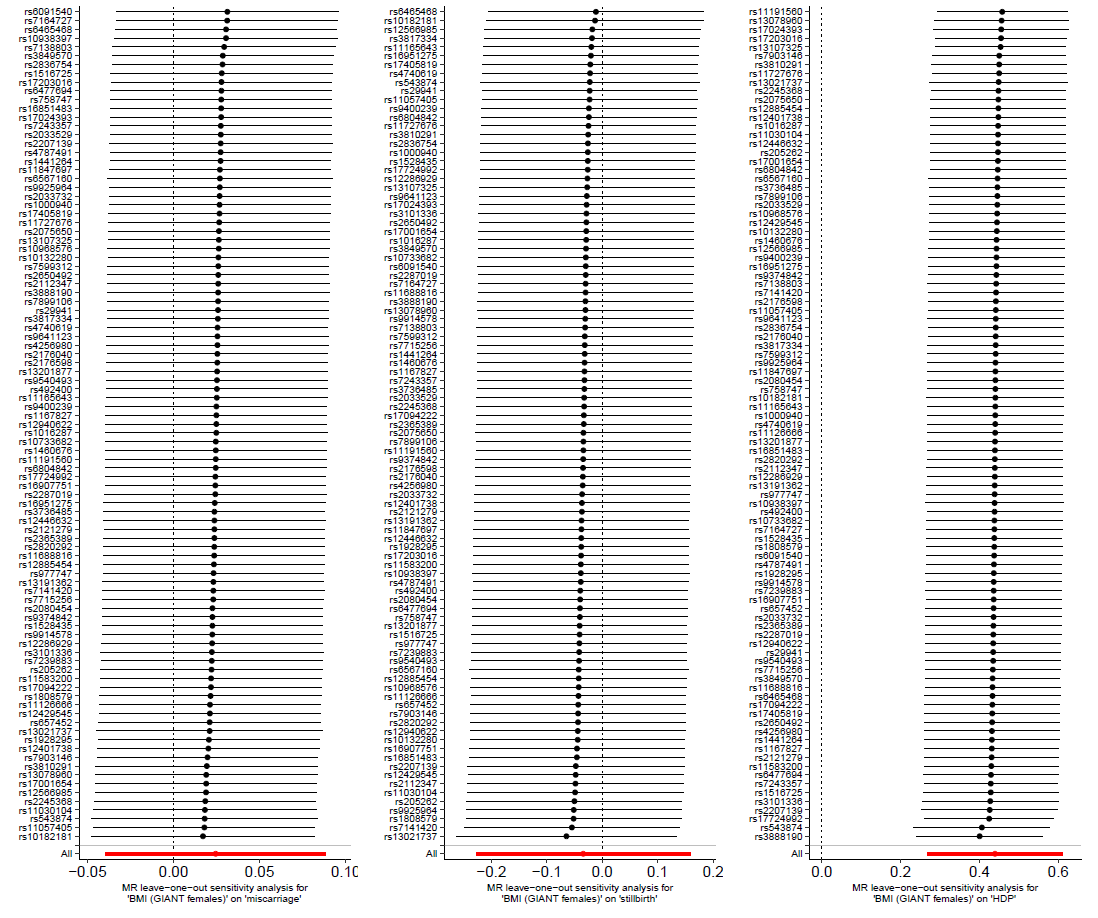

### Supplementary figure 9B

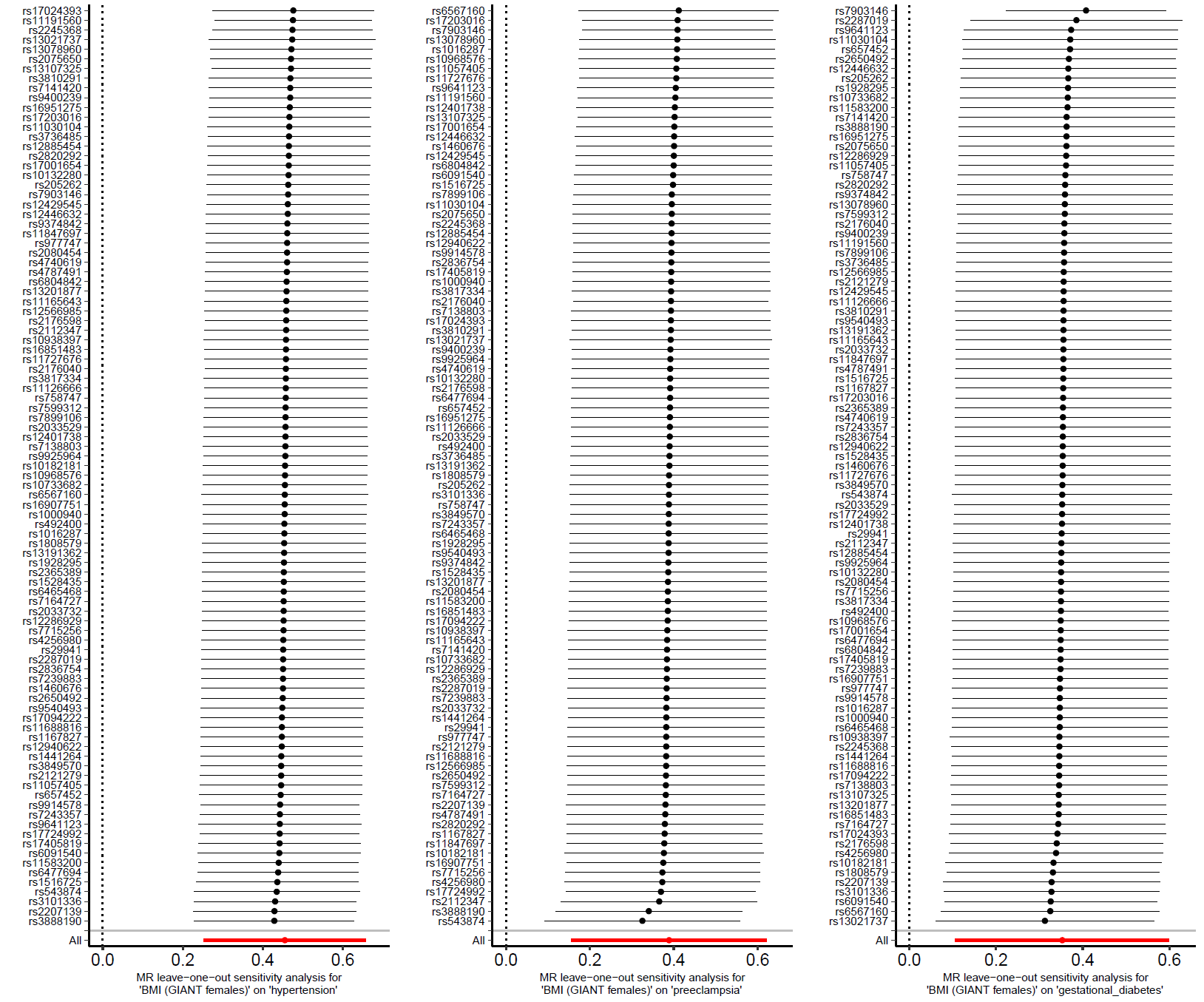

### Supplementary figure 9C

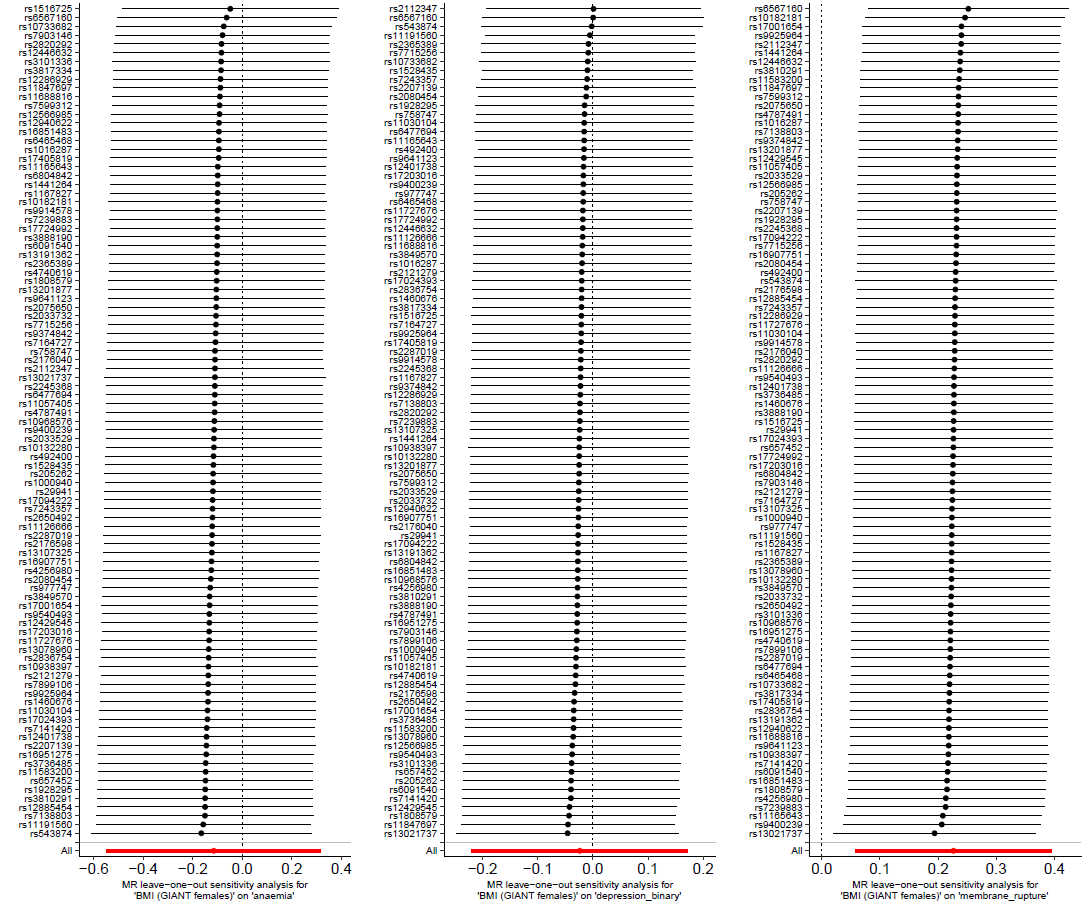

### Supplementary figure 9D

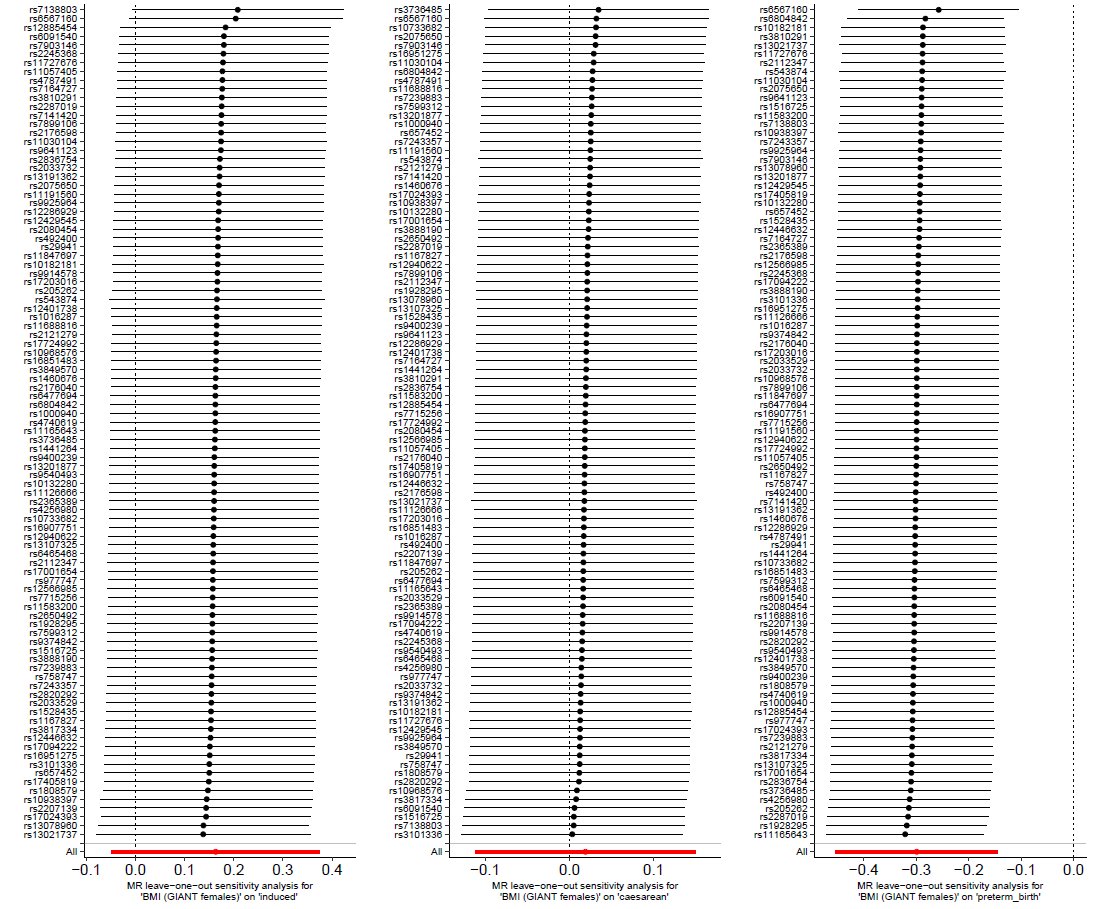

### Supplementary figure 9E

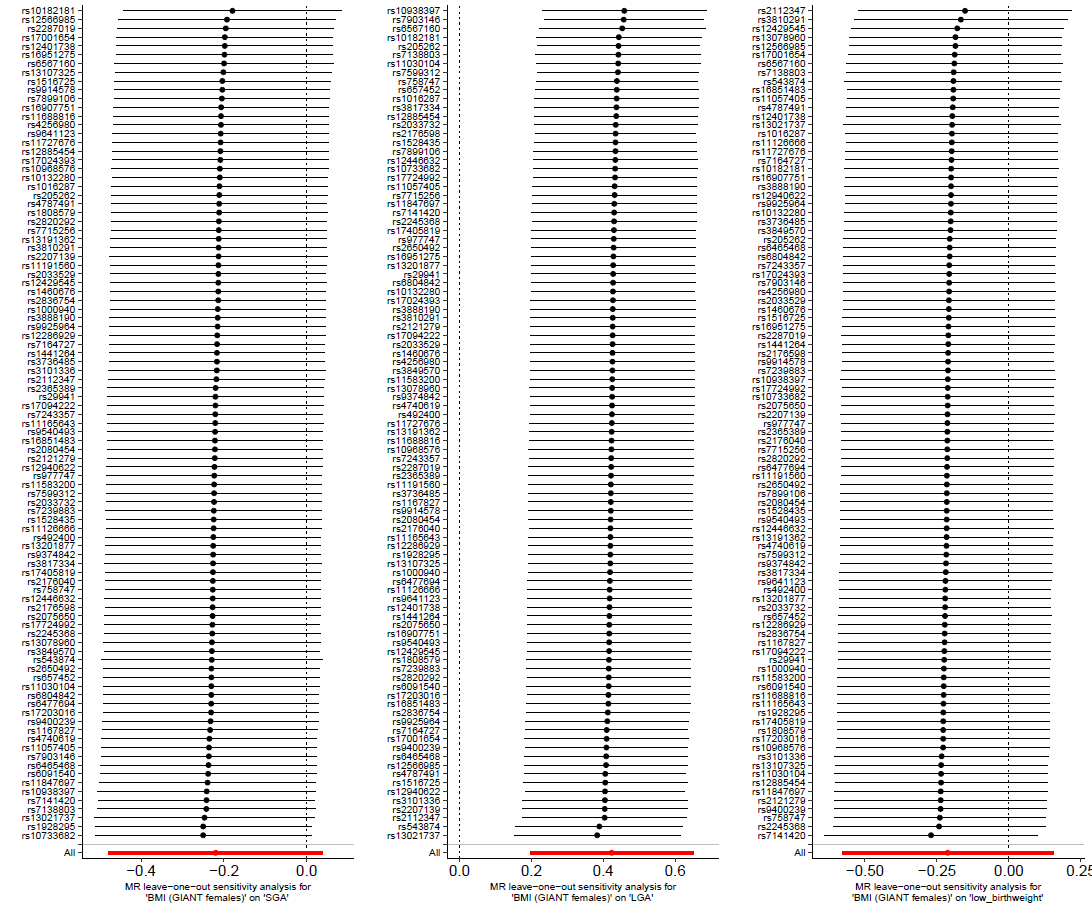

### Supplementary figure 9F

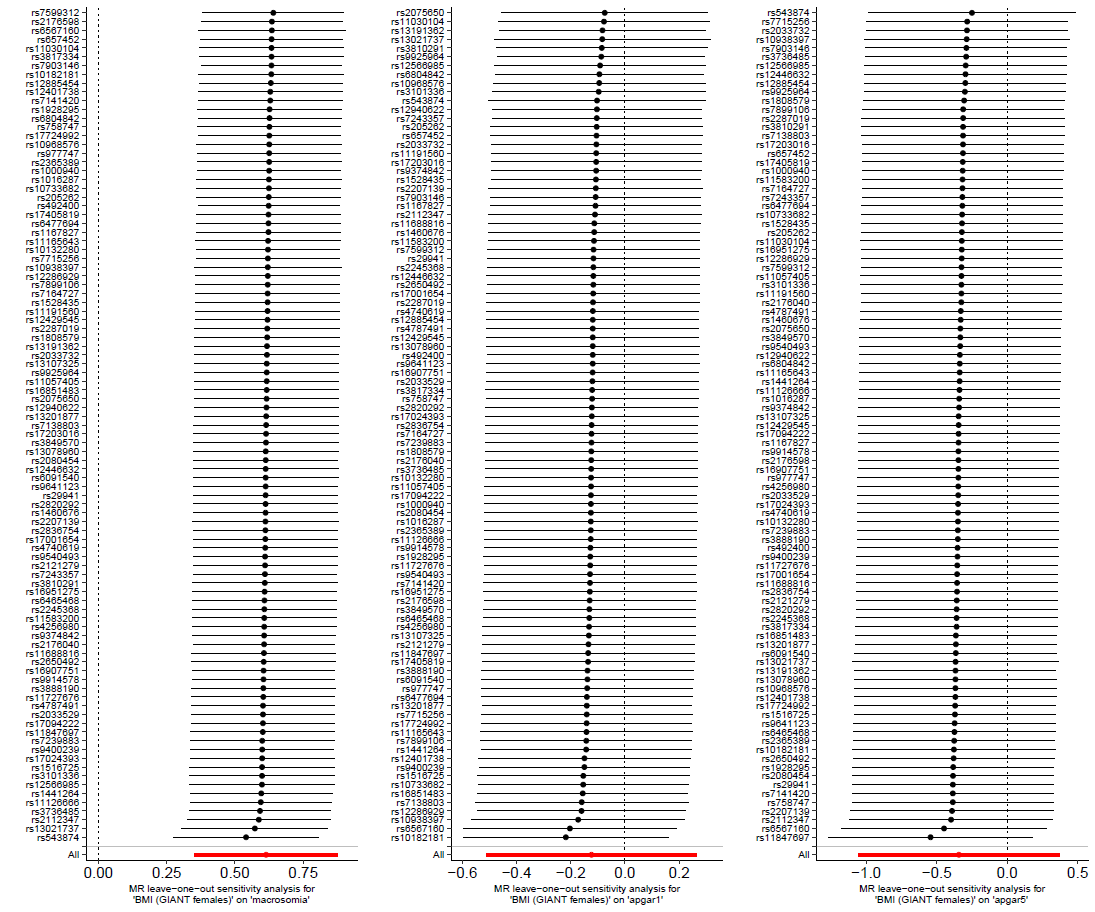

### Supplementary figure 9G

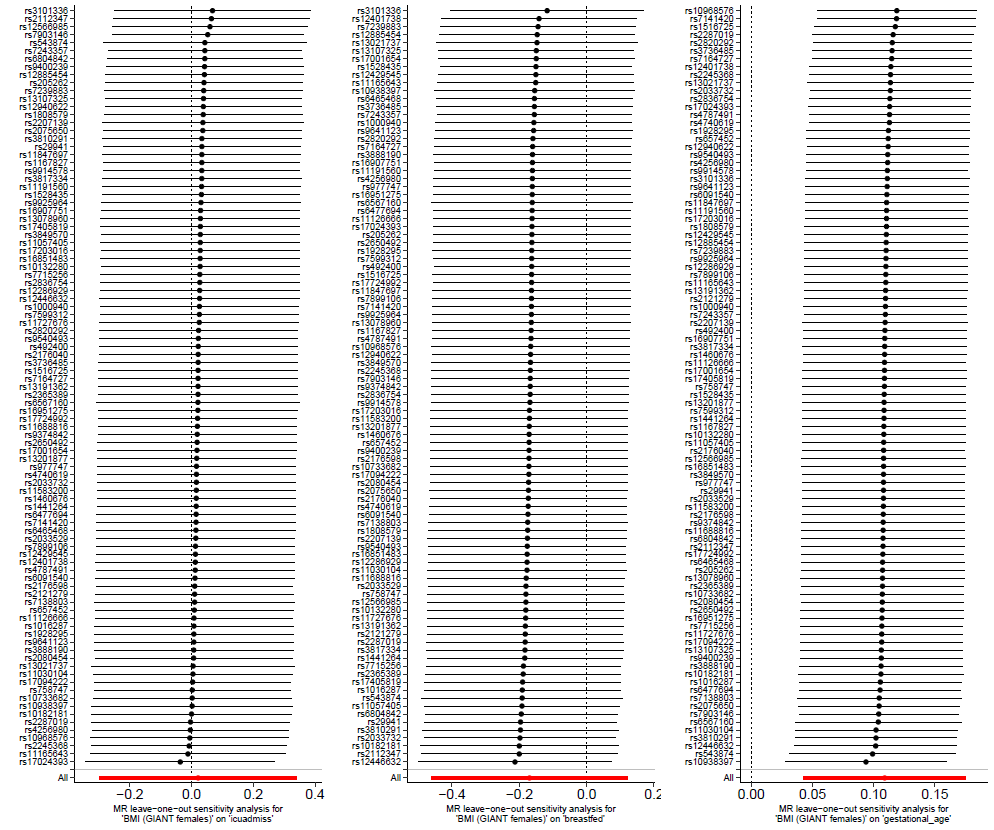

### Supplementary figure 9H

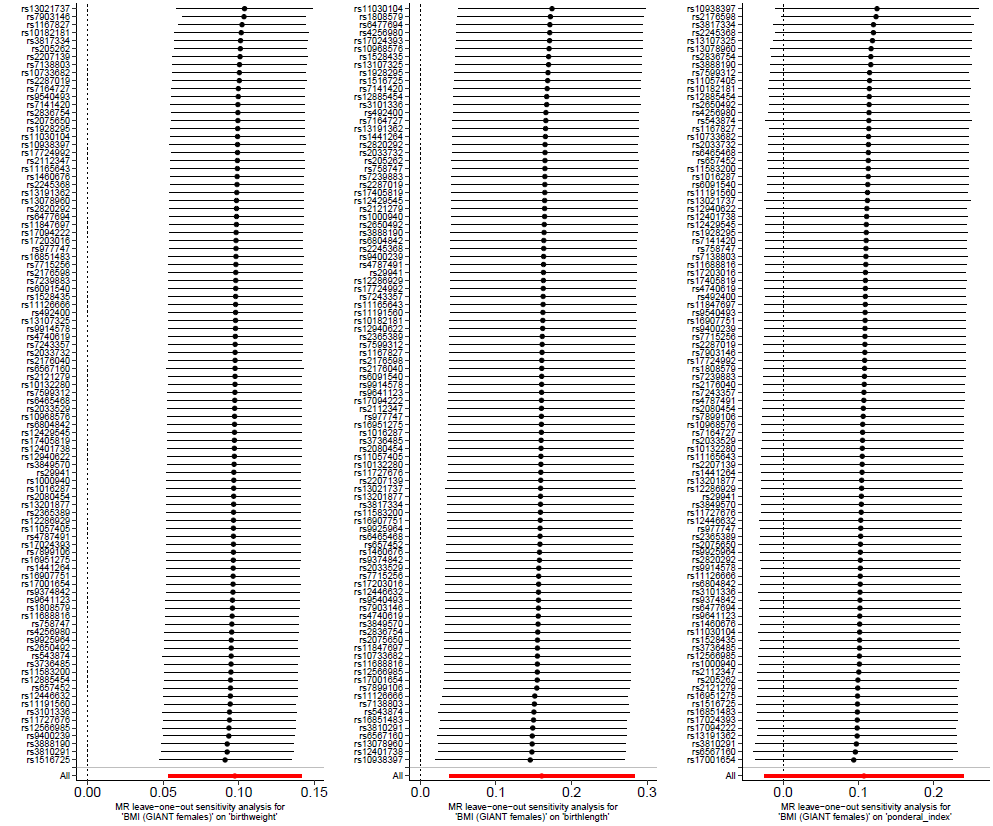

Results are expressed as log odds ratio for binary outcomes or mean difference in standard deviation (SD) unit for continuous outcomes per SD unit of maternal BMI. The figure is presented in eight panels according to the outcome as follows. **A**: miscarriage, stillbirth and hypertensive disorders of pregnancy; **B**: gestational hypertension, pre-eclampsia and gestational diabetes; **C**: maternal anaemia, perinatal depression and pre-labour rupture of membranes; **D**: induction of labour, caesarian section and pre-term birth; **E**: small for gestational age, large for gestational age and low birth weight; **F**: high birth weight, low Apgar score at 1 minute and low Apgar score at 5 minutes; **G**: NICU admission, breastfeeding initiation and gestational age; **H**: birth weight, birth length and ponderal index. BMI: body mass index; IVW: inverse variance weighted method; NICU: neonatal intensive care unit.

### Supplementary figure 10 - Comparison of Mendelian randomization results using different methods for (A) binary and (B) continuous outcomes

## A

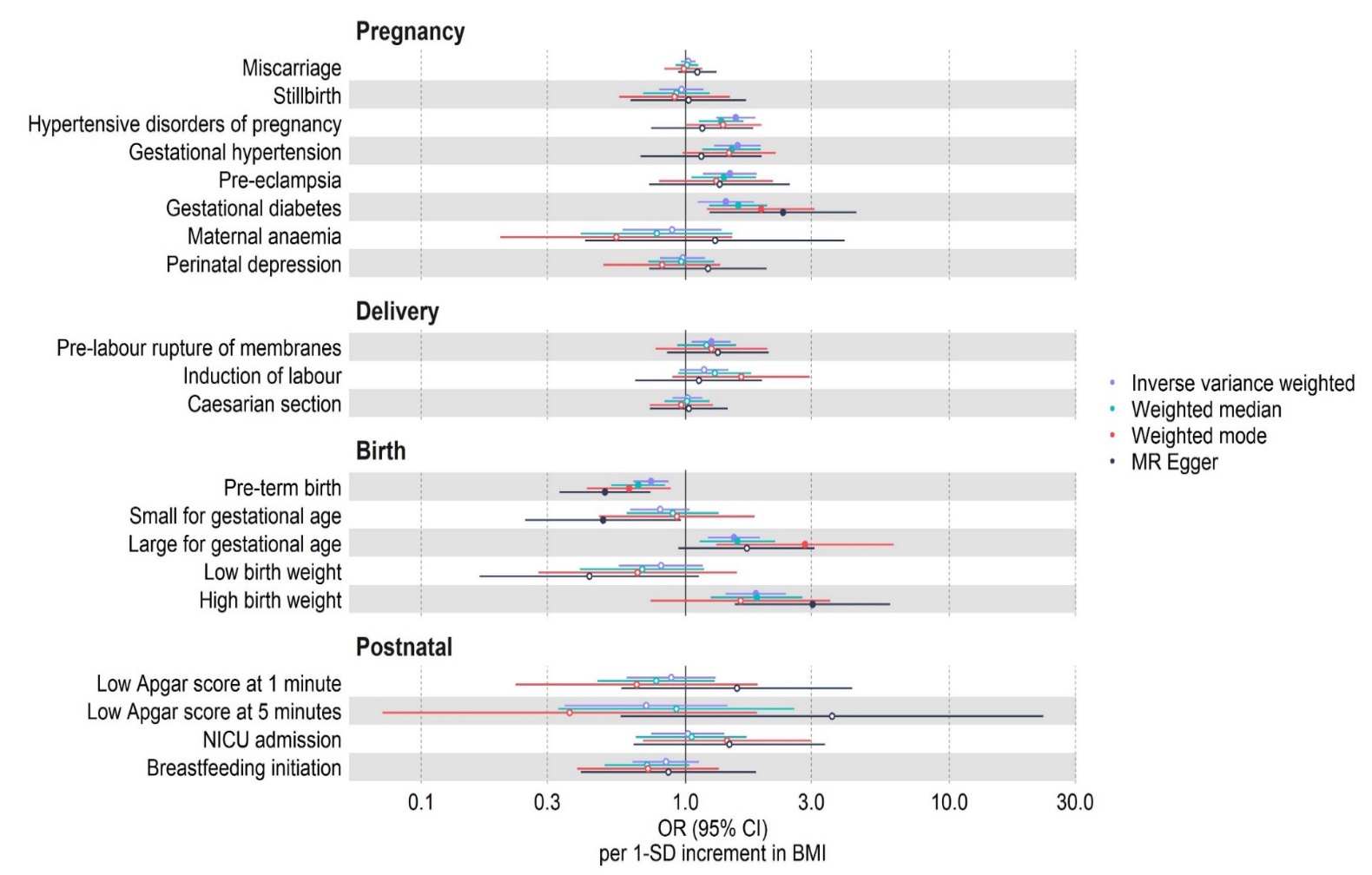

## B

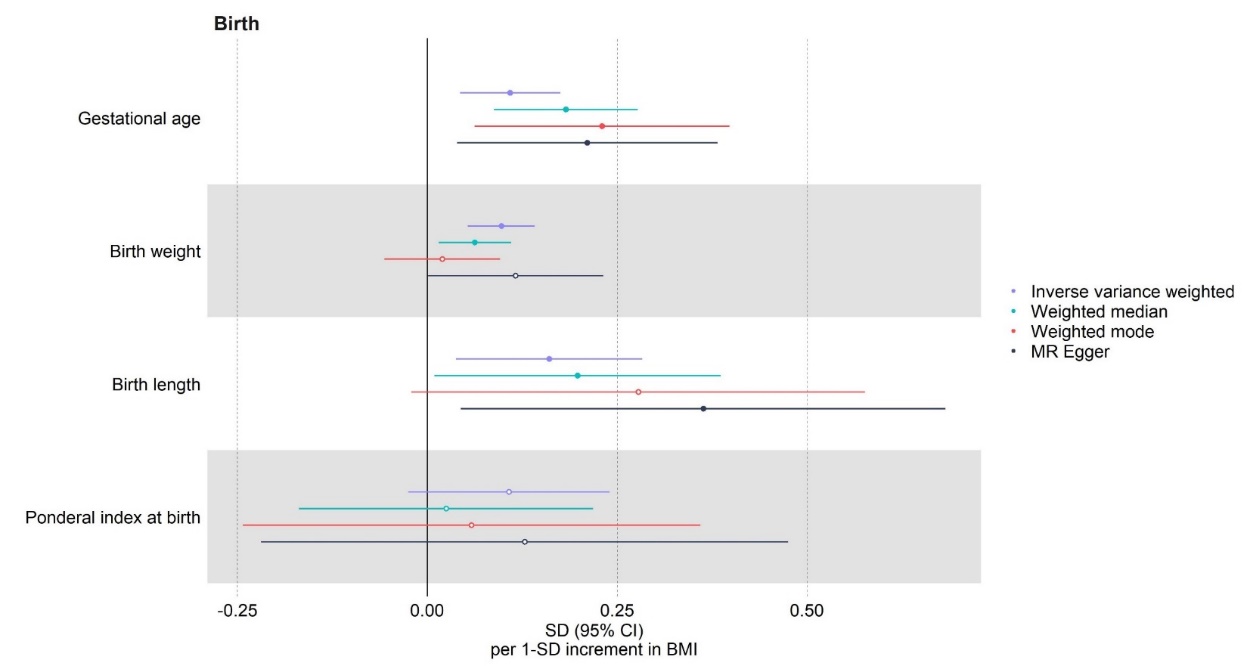

Results are expressed as log odds ratio (OR) for binary outcomes (A) or mean difference in standard deviation (SD) unit for continuous outcomes (B) per SD unit of maternal BMI. Study-specific results were estimated using different Mendelian randomization methods and combined using fixed-effect metanalysis. BMI: body mass index; NICU: neonatal intensive care unit; SD: standard deviation units.

### Supplementary figure 11 - Comparison of Mendelian randomization IVW results using summary data for the SNP-outcomes association unadjusted or adjusted for offspring genotype for (A) binary and (B) continuous outcomes

## A

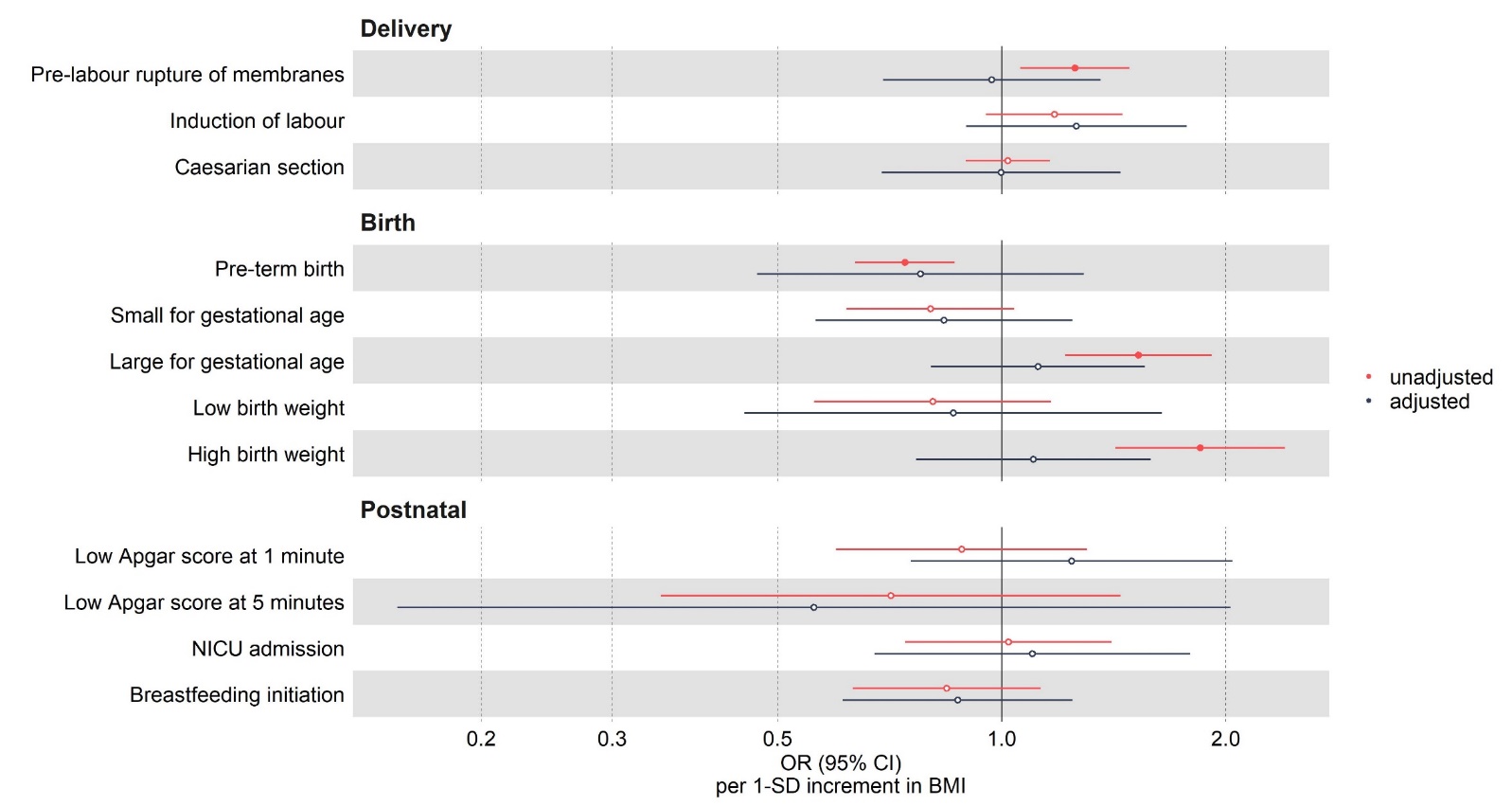

## B

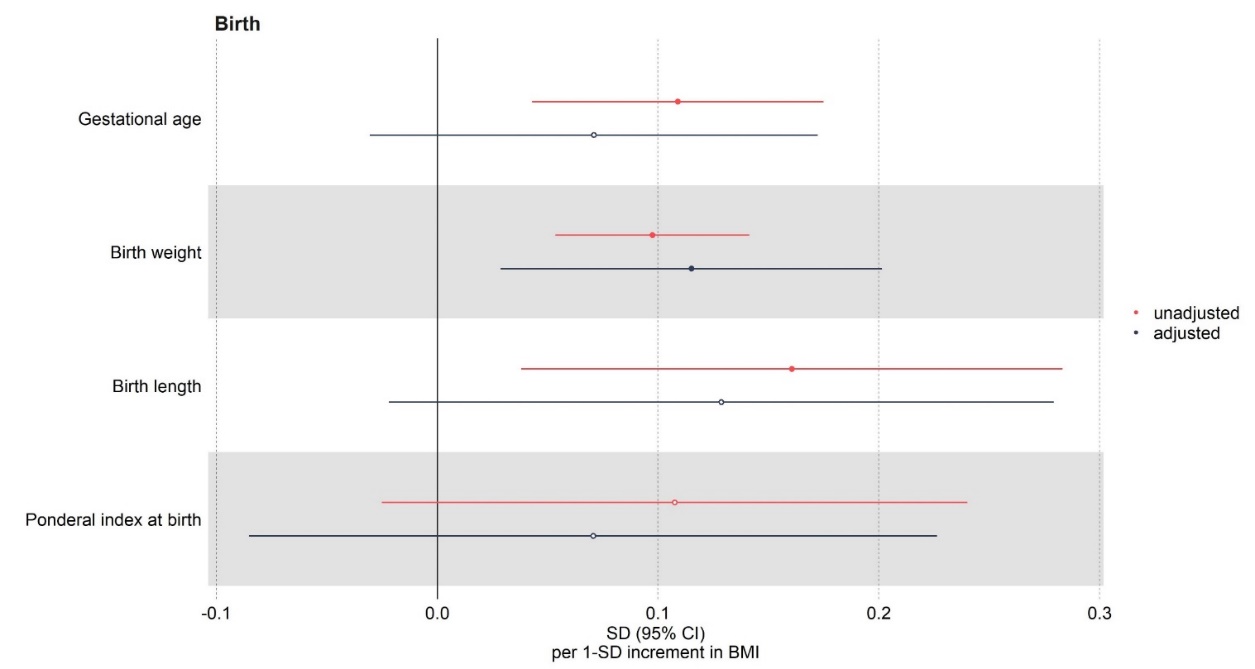

Results are expressed as log odds ratio (OR) for binary outcomes (A) or mean difference in standard deviation (SD) unit for continuous outcomes (B) per SD unit of maternal BMI. Study-specific results were estimated using IVW and combined using fixed-effect metanalysis. Only outcomes selected a priori were included in this analysis. IVW: inverse variance weighted method; BMI: body mass index; NICU: neonatal intensive care unit; SD: standard deviation units.

### Supplementary figure 12 - Partner negative control - study specific and pooled – maternal - associations using a fixed effect meta-analysis for (A) binary outcomes and (B) continuous outcomes

## A

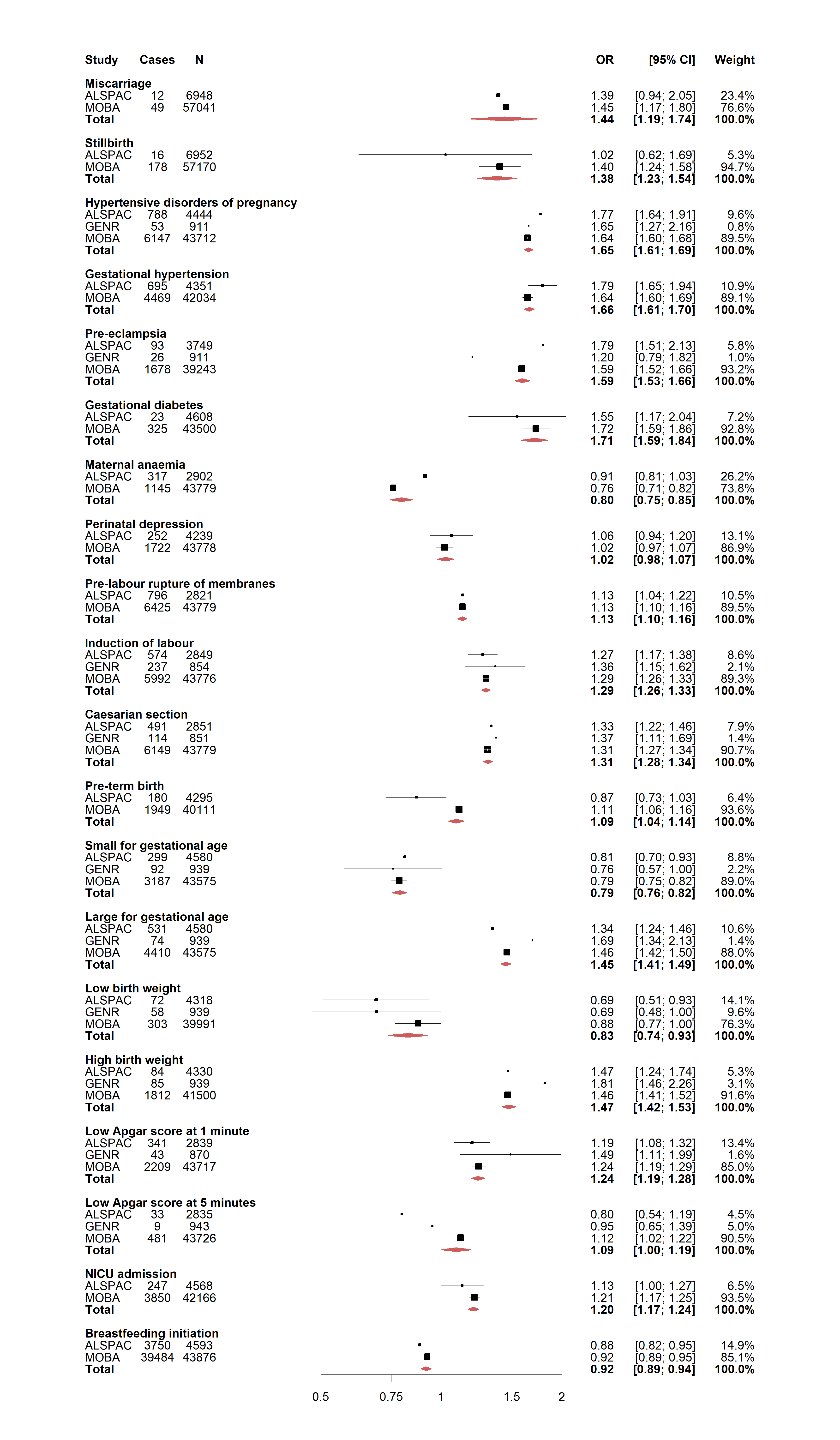

## B

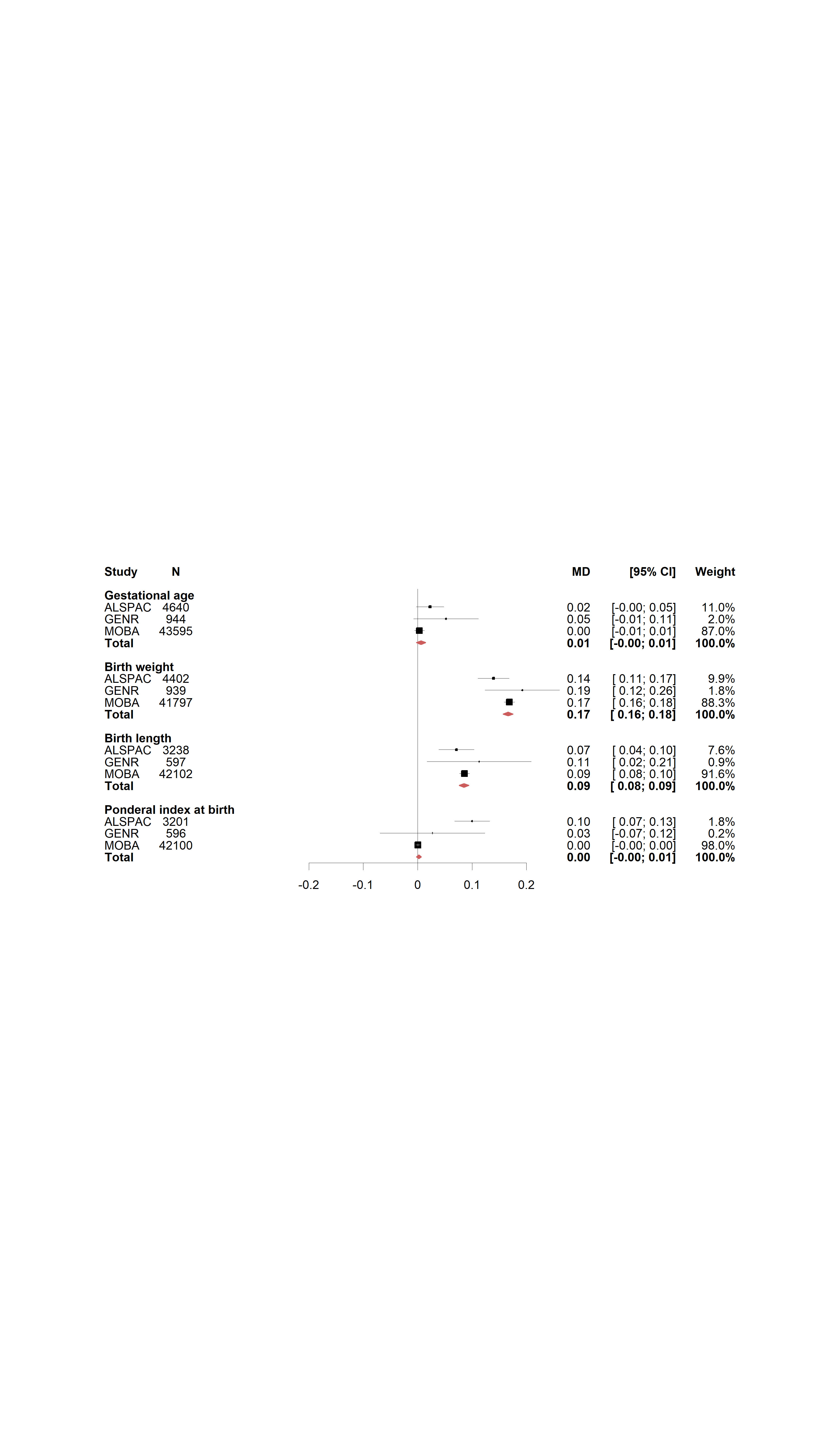

Results are expressed as odds ratio (OR) for binary outcomes (A) or mean difference in standard deviation (SD) unit for continuous outcomes (B) per SD unit of maternal BMI. Study-specific results for maternal BMI are presented for each cohort included in the partner negative control analyses and combined using fixed-effect metanalysis. Multivariable regression results were adjusted for paternal BMI, maternal age, parity, education, smoking during pregnancy, alcohol use during pregnancy and offspring sex where available. BMI: body mass index; NICU: neonatal intensive care unit; SD: standard deviation units.

### Supplementary figure 13 - Partner negative control - study specific and pooled – paternal - associations using a fixed effect meta-analysis for (A) binary outcomes and (B) continuous outcomes

## A

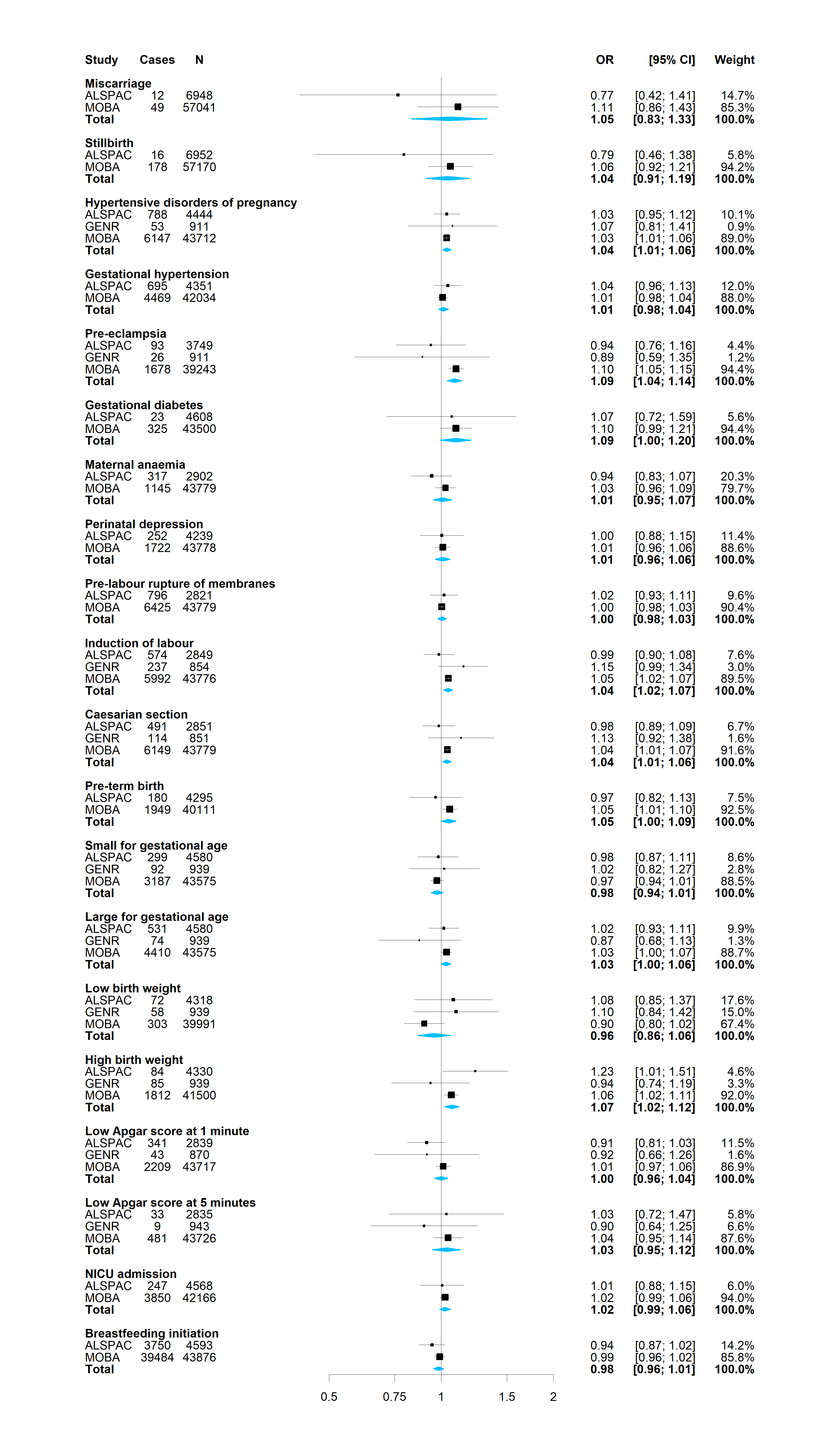

## B

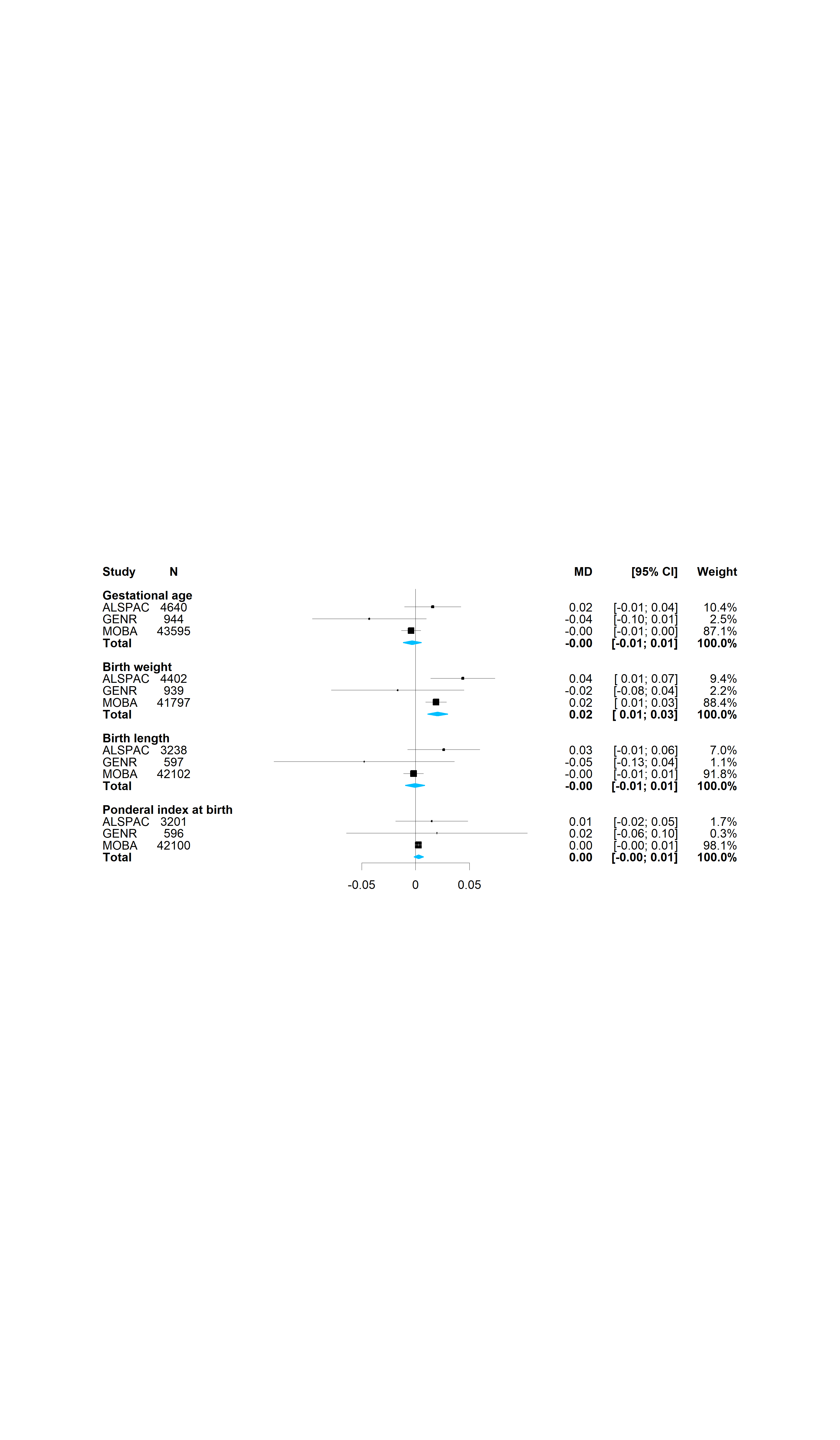

Results are expressed as odds ratio (OR) for binary outcomes (A) or mean difference in standard deviation (SD) unit for continuous outcomes (B) per SD unit of paternal BMI. Study-specific results for paternal BMI are presented for each cohort included in the partner negative control analyses and combined using fixed-effect metanalysis. Paternal negative control results were adjusted for maternal BMI, paternal age, number of children, paternal education, paternal smoking, paternal alcohol use and offspring sex where available. BMI: body mass index; NICU: neonatal intensive care unit; SD: standard deviation units.

### Supplementary figure 14 - Partner negative control – sensitivity analyses – for (A) binary outcomes and (B) continuous outcomes: ALSPAC

## A

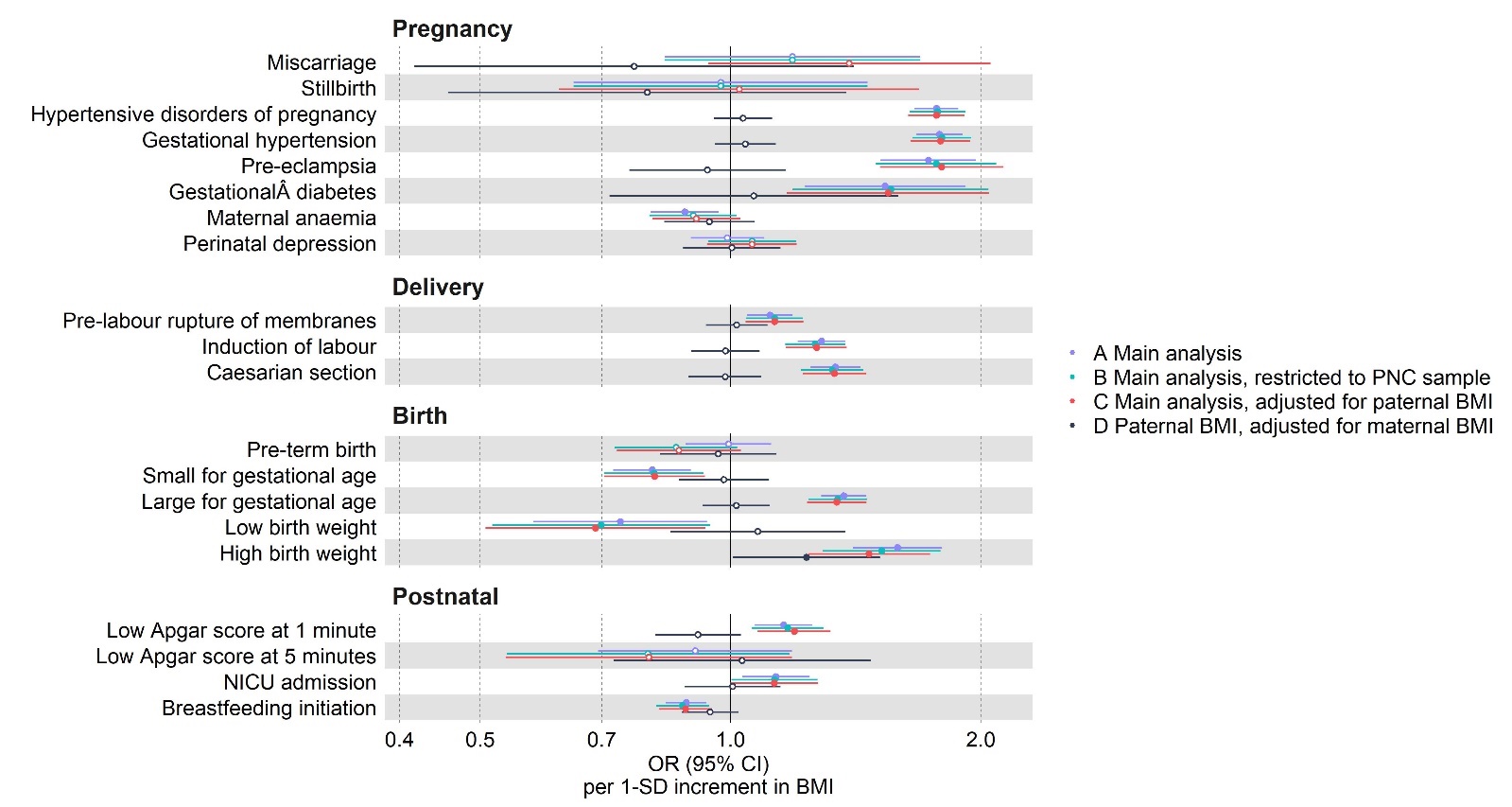

## B

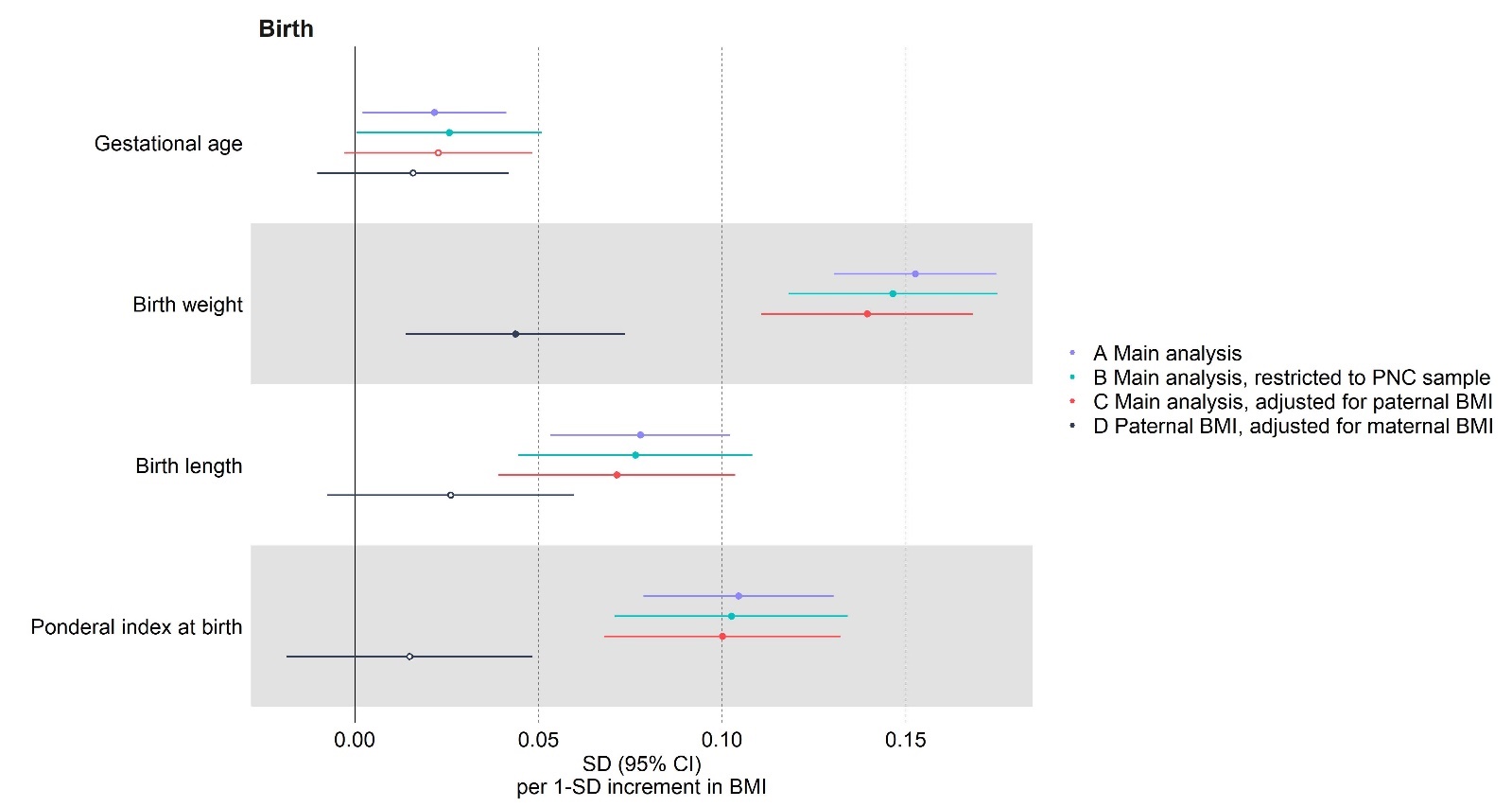

Results are expressed as odds ratio (OR) for binary outcomes (A) or mean difference in standard deviation (SD) unit for continuous outcomes (B) per SD unit of maternal BMI (for analyses A-C) and paternal BMI (for analysis D) respectively. The following study specific results for ALSPAC are presented: A Main analysis (per SD unit of maternal BMI); B Main analysis, restricted to PNC sample (per SD unit of maternal BMI); C Main analysis adjusted for paternal BMI (per SD unit of maternal BMI); D Paternal BMI adjusted for maternal BMI (per SD unit of paternal BMI). Our main comparison of interest here is between C and D, however we additionally check for sample selection or selection bias by including analyses A and B as sensitivity analyses. As analysis B restricts to only those with paternal BMI data, we would expect analyses A and B to be similar if there was little to no selection bias. as BMI: body mass index; NICU: neonatal intensive care unit; SD: standard deviation units. PNC: Paternal negative control.

### Supplementary figure 15 - Partner negative control – sensitivity analyses – for (A) binary outcomes and (B) continuous outcomes: MoBa

## A

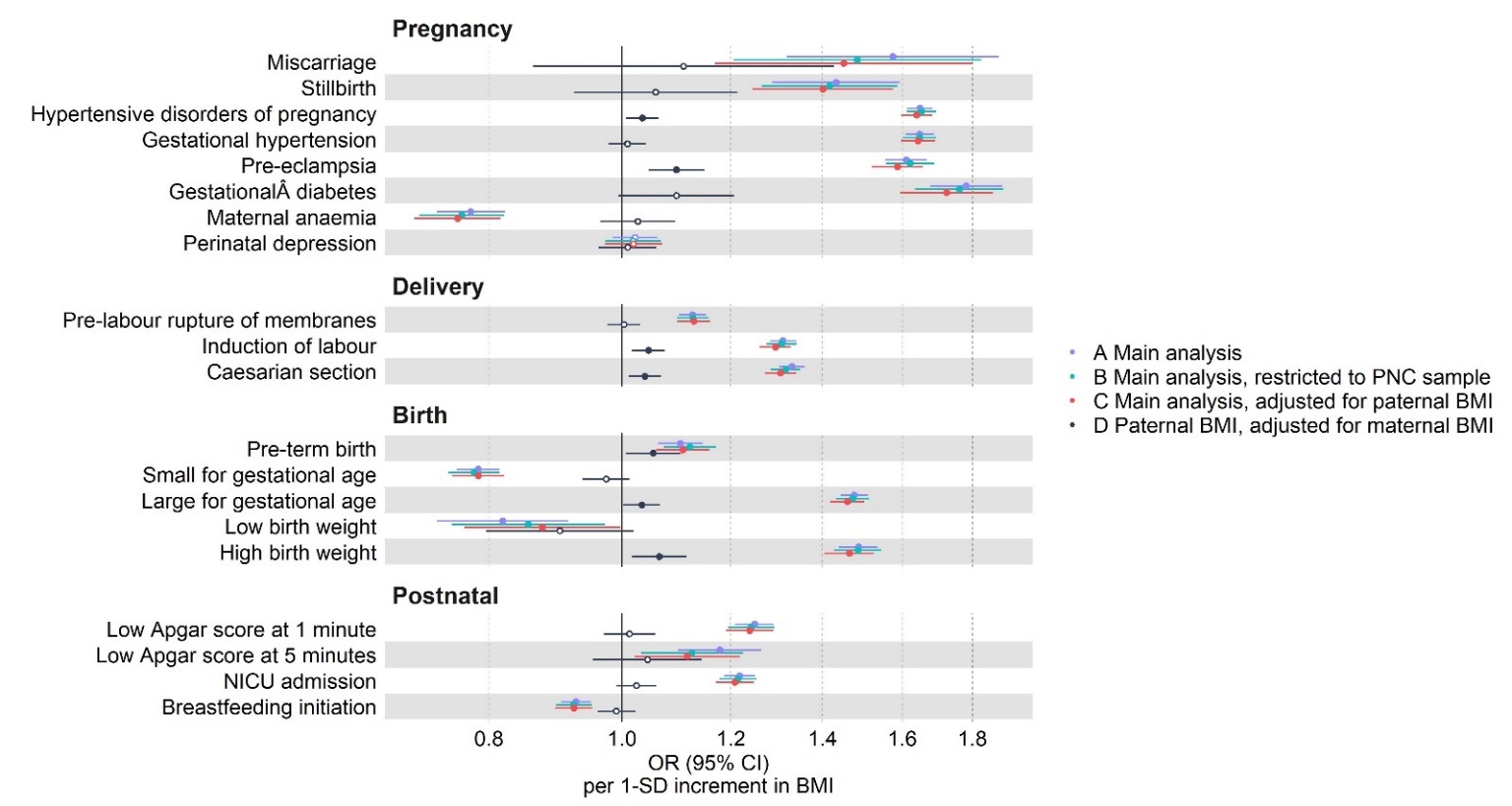

## B

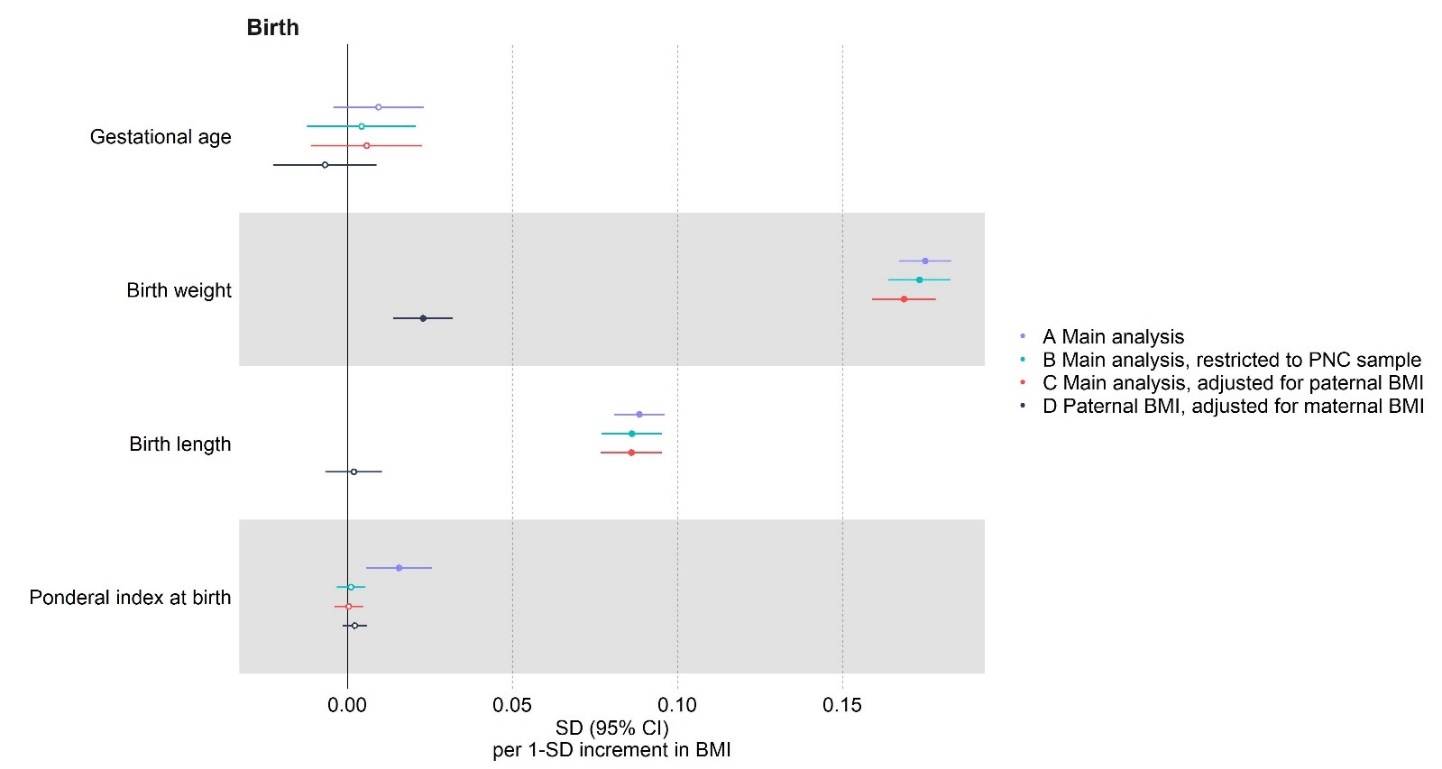

Results are expressed as odds ratio (OR) for binary outcomes (A) or mean difference in standard deviation (SD) unit for continuous outcomes (B) per SD unit of maternal BMI (for analyses A-C) and paternal BMI (for analysis D) respectively. The following study specific results for MoBa are presented: A Main analysis (per SD unit of maternal BMI); B Main analysis, restricted to PNC sample (per SD unit of maternal BMI); C Main analysis adjusted for paternal BMI (per SD unit of maternal BMI); D Paternal BMI adjusted for maternal BMI (per SD unit of paternal BMI). Our main comparison of interest here is between C and D, however we additionally check for sample selection or selection bias by including analyses A and B as sensitivity analyses. As analysis B restricts to only those with paternal BMI data, we would expect analyses A and B to be similar if there was little to no selection bias. as BMI: body mass index; NICU: neonatal intensive care unit; SD: standard deviation units. PNC: Paternal negative control.

### Supplementary figure 16 - Partner negative control – sensitivity analyses – for (A) binary outcomes and (B) continuous outcomes: GenR

## A

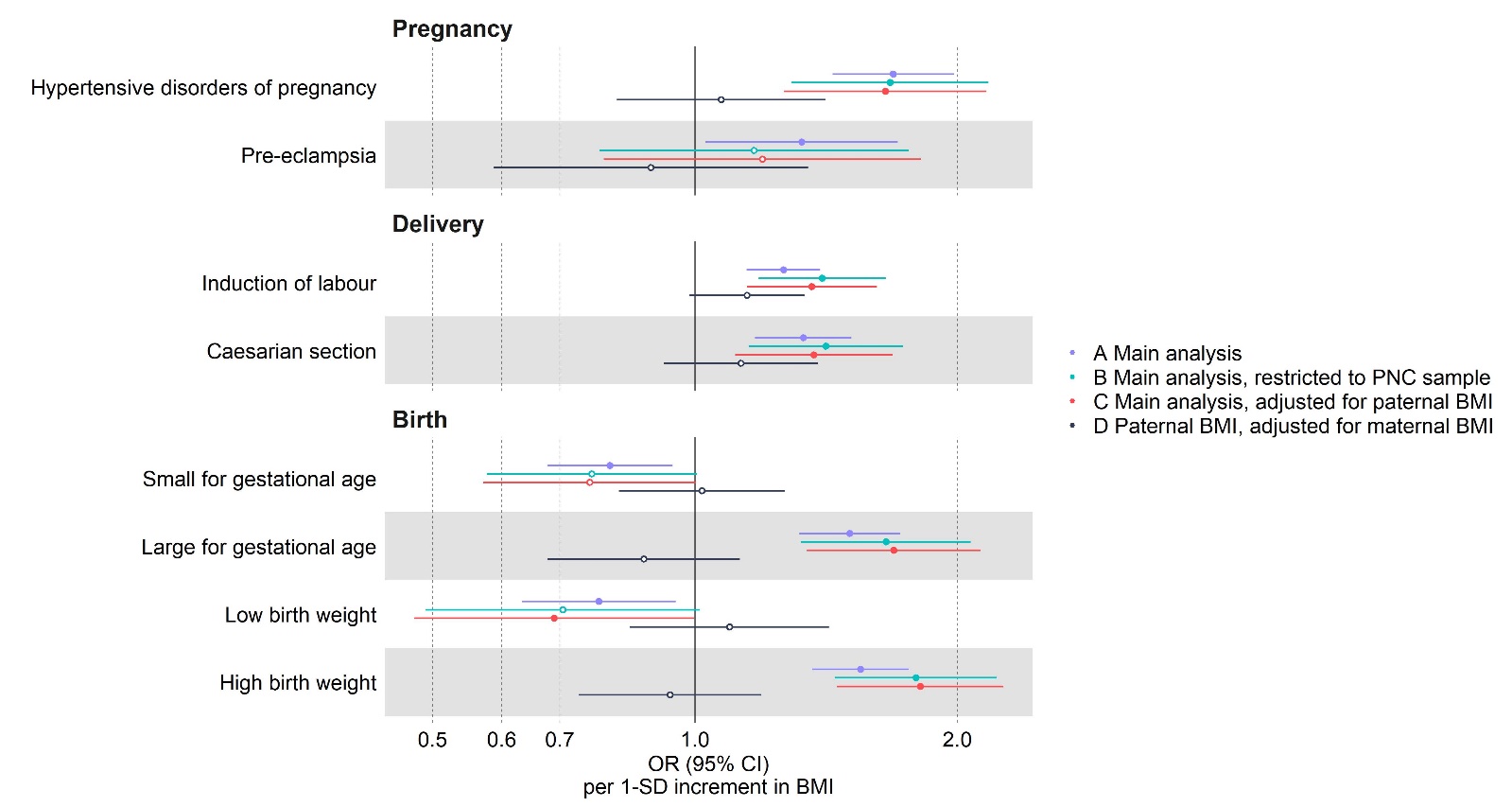

## B

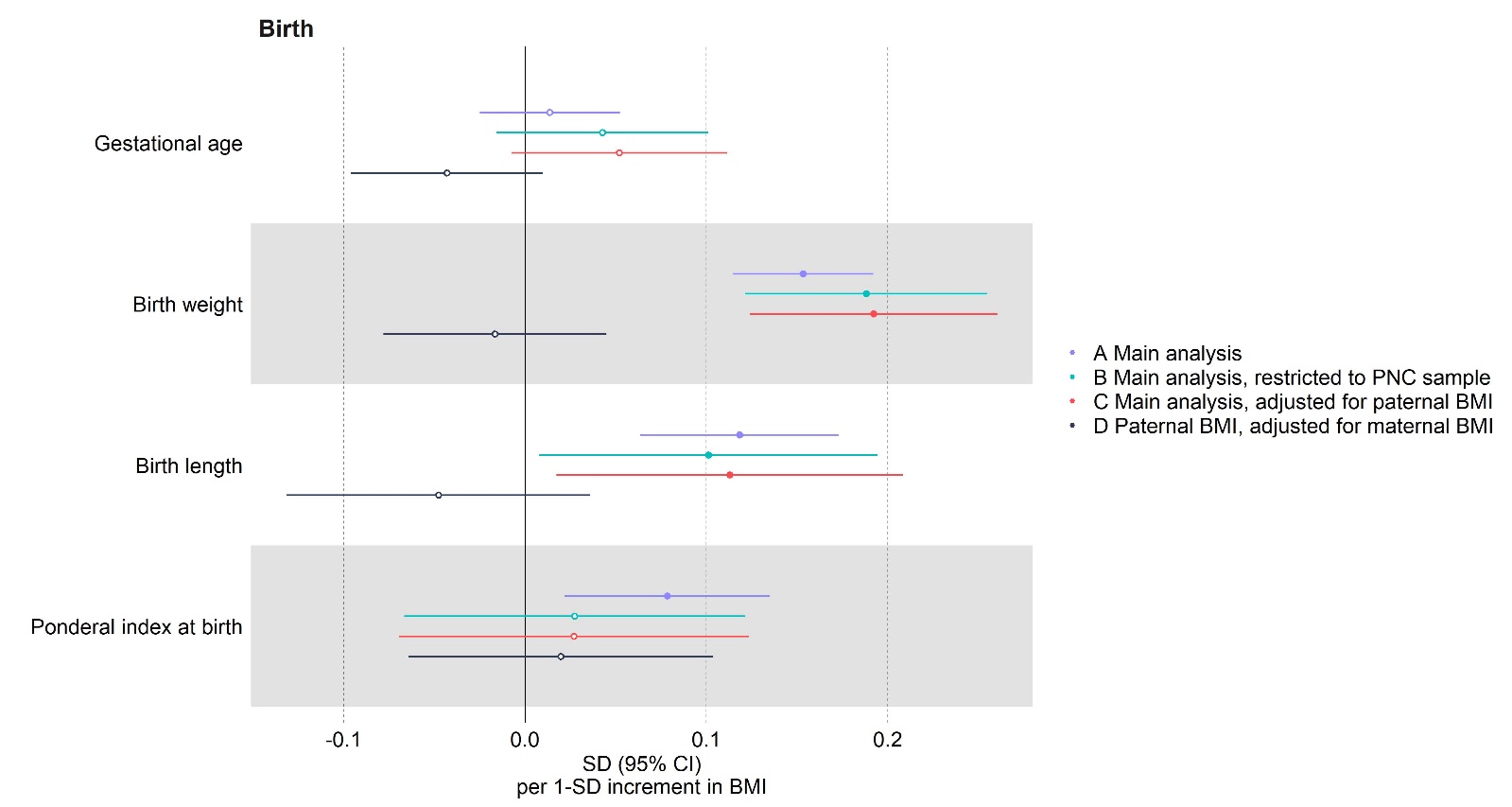

Results are expressed as odds ratio (OR) for binary outcomes (A) or mean difference in standard deviation (SD) unit for continuous outcomes (B) per SD unit of maternal BMI (for analyses A-C) and paternal BMI (for analysis D) respectively. The following study specific results for Gen-R are presented: A Main analysis (per SD unit of maternal BMI); B Main analysis, restricted to PNC sample (per SD unit of maternal BMI); C Main analysis adjusted for paternal BMI (per SD unit of maternal BMI); D Paternal BMI adjusted for maternal BMI (per SD unit of paternal BMI). Our main comparison of interest here is between C and D, however we additionally check for sample selection or selection bias by including analyses A and B as sensitivity analyses. As analysis B restricts to only those with paternal BMI data, we would expect analyses A and B to be similar if there was little to no selection bias. as BMI: body mass index; NICU: neonatal intensive care unit; SD: standard deviation units. PNC: Paternal negative control.

### Supplementary figure 17 - Comparison of multivariable regression pooled estimates across studies using fixed- or random-effect metanalyses split by studies reporting pre and during pregnancy weight for (A) binary outcomes and (B) continuous outcomes.

## A

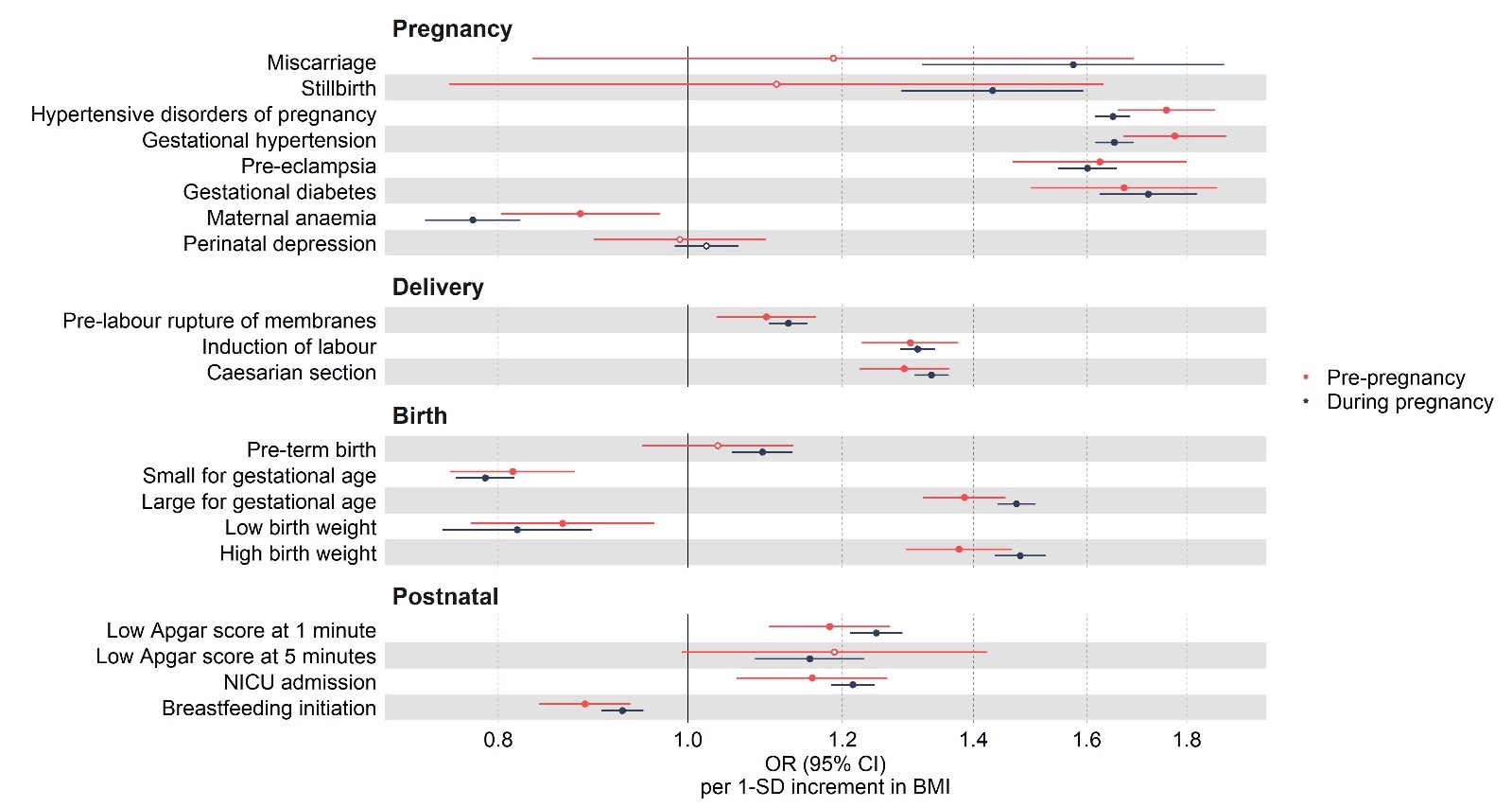

## B

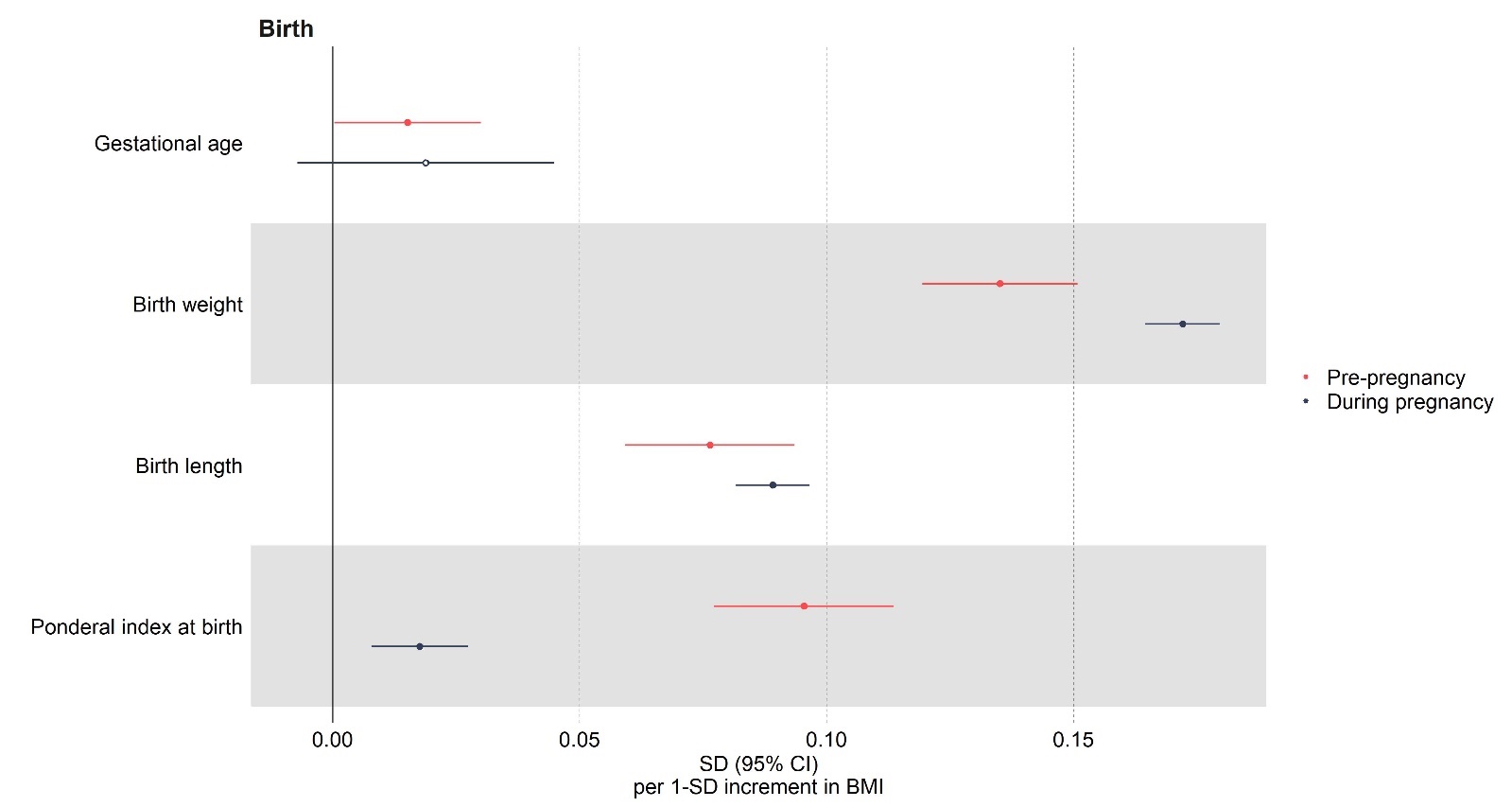
